# Honey and *Nigella sativa* against COVID-19 in Pakistan (HNS-COVID-PK): A multi-center placebo-controlled randomized clinical trial

**DOI:** 10.1101/2020.10.30.20217364

**Authors:** Sohaib Ashraf, Shoaib Ashraf, Moneeb Ashraf, Muhammad Ahmad Imran, Larab Kalsoom, Uzma Nasim Siddiqui, Iqra Farooq, Zaighum Habib, Sidra Ashraf, Muhammad Ghufran, Muhammad Kiwan Akram, Nighat Majeed, Zain-ul-Abdin, Rutaba Akmal, Sundas Rafique, Khawar Nawaz, Muhammad Ismail K Yousaf, Sohail Ahmad, Muhammad Sarmad Shahab, Muhammad Faisal Nadeem, Muhammad Azam, Hui Zheng, Amber Malik, Mahmood Ayyaz, Talha Mahmud, Qazi Abdul Saboor, Ali Ahmad, Muhammad Ashraf, Mateen Izhar, for the COALITION COVID-19 Shaikh Zayed, Abubakar Hilal, Arz Muhammad, Zeeshan Shaukat, Ayesha Khaqan, Kanwal Hayat, Shahroze Arshad, Muhammad Hassan, Abeer-bin-Awais, Ammara Ahmad, Tayyab Mughal, Abdur Rehman Virk, Muhammad Umer, Muhammad Suhail, Sibgha Zulfiqar, Saulat Sarfraz, Muhammad Imran Anwar, Ayesha Humayun, R A Khokhar, S Siddique

## Abstract

**BACKGROUND:** No definitive treatment exists for Coronavirus Disease 2019 (COVID-19). Honey and *Nigella sativa* (HNS) have established antiviral, antibacterial, anti-inflammatory and immunomodulatory properties. Hence, we investigated efficacy of HNS against COVID-19. wide

**METHODS:** We conducted a multicenter, placebo-controlled, randomized clinical trial at 4 centers in Pakistan. RT-PCR confirmed COVID-19 adults showing moderate or severe disease were enrolled in the study. Patients presenting with multi-organ failure, ventilator support, and chronic diseases (except diabetes mellitus and hypertension) were excluded. Patients were randomly assigned in 1:1 ratio to receive either honey (1 gm/Kg/day) and *Nigella sativa* seeds (80 mg/Kg/day) or placebo up-to 13 days along with standard care. The outcomes included symptom alleviation, viral clearance, and a 30-day mortality in intention-to-treat population. This trial was registered with ClinicalTrials.gov, **NCT04347382**.

**RESULTS:** Three hundred and thirteen patients - 210 moderate and 103 severe - underwent randomization from April 30 to July 29, 2020. Among these, 107 were assigned to HNS whereas 103 to placebo for moderate cases. For severe cases, 50 were given HNS and 53 were given placebos. HNS resulted in ∼50% reduction in time taken to alleviate symptoms as compared to placebo (Moderate (4 versus 7 days), Hazard Ratio [HR]: 6.11; 95% Confidence Interval [CI]: 4.23-8.84, P<0.0001 and severe (6 versus 13 days) HR: 4.04; 95% CI, 2.46-6.64, P<0.0001). HNS also cleared the virus 4 days earlier than placebo group in moderate (6 versus 10 days, HR: 5.53; 95% CI: 3.76-8.14, P<0.0001) and severe cases (8.5 versus 12 days, HR: 4.32; 95% CI: 2.62-7.13, P<0.0001). HNS further led to a better clinical score on day 6 with normal activity resumption in 63.6% versus 10.9% among moderate cases (OR: 0.07; 95% CI: 0.03-0.13, P<0.0001) and hospital discharge in 50% versus 2.8% in severe cases (OR: 0.03; 95% CI: 0.01-0.09, P<0.0001). In severe cases, mortality rate was four-fold lower in HNS group than placebo (4% versus 18.87%, OR: 0.18; 95% CI: 0.02-0.92, P=0.029). No HNS-related adverse effects were observed.

**CONCLUSION:** HNS significantly improved symptoms, viral clearance and mortality in COVID-19 patients. Thus, HNS represents an affordable over the counter therapy and can either be used alone or in combination with other treatments to achieve potentiating effects against COVID-19.

**FUNDING:** Funded by Smile Welfare Organization, Shaikh Zayed Medical Complex, and Services Institute of Medical Sciences.

## BACKGROUND

The Coronavirus Disease 2019 (COVID-19) pandemic has infected more than forty million people and has resulted in more than a million deaths worldwide. In the absence of an effective prophylactic vaccine, there is a dire need for finding effective treatments for COVID-19 patients. At a minimum, an ideal treatment should expedite symptomatic recovery, decrease viral transmission in the community with earlier viral clearance in the infected patients, and reduce mortality. In this context, treatments including hydroxychloroquine/azithromycin, lopinavir-ritonavir, remdesivir, dexamethasone, convalescent plasma and antibody therapies have shown some efficacy.^1-4^ However, there is still a long way to go before we have an effective treatment regimen for COVID-19. To this end, we have conducted a clinical trial in which we have investigated the potential efficacy of a combination of honey and *Nigella sativa* (HNS) in treating COVID-19 patients.

Both components of HNS have anti-viral, anti-microbial, anti-inflammatory and immune-modulatory effects with proven safety profiles. ^5-8^ The beneficial effects of honey against different viruses including rubella virus, Herpes Simplex virus, Hepatitis virus, and Varicella-Zoster virus have been reported earlier.^6^ Moreover, in-silico molecular docking studies have shown that six flavonoid compounds from honey might inhibit severe acute respiratory syndrome coronavirus 2 (SARS-CoV-2) replication by binding to the viral 3-chymotrypsin-like-cysteine protease.^9^ Honey also has strong antibacterial activity against clinically important gram-positive bacteria (methicillin-resistant *Staphylococcus aureus*) and gram-negative bacteria (*Pseudomonas aeruginosa, Enterobacter spp, and Klebsiella*). Additionally, honey has shown synergism with other antibiotics like oxacillin, tetracycline, imipenem and meropenem.^6^ The use of honey not only improves the proliferation of T and B lymphocytes, but also their phagocytic activity. It additionally inhibits the expression of vital pro-inflammatory cytokines such as interleukin (IL) 1 beta and IL-6. Lymphocyte-mediated antiviral activity has proven to be poorly effective against COVID-19, especially considering the exaggerated release of pro-inflammatory mediators despite lymphocytopenia. Thus, honey is postulated to play a pivotal role in fighting COVID-19.^10^ Its’ use has shown to be more beneficial in upper respiratory tract infections than usual care especially in the context of cough frequency and severity.^11^

*Nigella sativa* (NS), a widely used medicinal plant of the family Ranunculaceae and commonly known as Black Cumin/Kalonji, has shown to exert antiviral effects against various viruses such as human immunodeficiency virus and hepatitis C virus.^12^ It has also shown to decrease the replication of SARS-CoV in-vitro in cell cultures.^13^ Molecular docking studies have shown that some of its’ components such as nigelledine, α-hederin and thymoquinone have high affinity with several SARS-CoV-2 enzymes and proteins. In fact, they exhibit an energy complex score better than that of chloroquine, hydroxychloroquine and favipiravir – the drugs that have shown some anti-SARS-CoV-2 activity. NS has shown antibacterial properties against many bacteria including drug sensitive and resistant S. *aureus, P. aeruginosa, Helicobacter pylori*, and *Escherichia coli*. Moreover, NS has shown synergism with streptomycin and gentamycin. It also demodulates the secretion of a number of pro-inflammatory mediators and improves helper-T cell (T4) and suppressor-T-cell (T8) ratio with increased natural killer (NK) cell activity. It also manifests potential radical scavenging.^14, 15^

As honey and *Nigella sativa* exhibit overlapping pharmacological profiles, we reasoned that the combination could be more effective in mitigating severity of the disease, controlling viral replication and curing COVID-19 patients. The combination has been used successfully in various disease conditions.^16-18^ We report here that the HNS treatment results in earlier recovery and viral clearance in COVID-19 patients.

## METHODOLOGY

### STUDY DESIGN

The study was an investigator-initiated, multicenter placebo controlled randomized trial with superiority framework conducted in four medical care facilities in Pakistan (Shaikh Zayed Post-Graduate Medical Complex, Services Institute of Medical Sciences, Doctor’s Lounge and Ali Clinic; all located in Lahore). The trial was approved by the institutional review boards of Shaikh Zayed Post-Graduate Medical Complex and Services Institute of Medical Sciences and supervised by an independent trial steering committee. The trial was conducted in accordance with principles of Good Clinical Practice Guidelines of the International Conference on Harmonization.

### PATIENTS /PARTICIPANTS

Suspected COVID-19 patients presenting with positive SARS-CoV-2 by RT-PCR of their nasopharyngeal swabs in International Organization for Standardization (ISO) certified Pakistan designated laboratories using quantitative real-time RT-PCR were screened. The virus nucleic acid positive, adult males and non-pregnant females, who presented to seek medical care within 96 h of ailment underwent randomization. Exclusion criteria included having mild to lacking clinical symptoms, inability to give written consent, multi-organ dysfunction, ventilator support or PaO2/FIO_2_ of less than 100, septic shock, known hypersensitivity to HNS and chronic illness other than hypertension and diabetes mellitus. Patients with positive SARS-CoV-2 screening during elective lists for any procedure were also excluded. Written informed consent was obtained from each participant or their legal representative if too unwell to provide consent.

### RANDOMIZATION AND MASKING

Eligible patients were stratified in a 1:1 ratio based upon the severity of their clinical symptoms into two groups: mild to moderate (cough, fever, sore throat, nasal congestion, malaise and/or shortness of breath), and severe cases (fever and/or cough along with pneumonia, severe dyspnea, respiratory distress, tachypnea (>30 breaths/min) or hypoxia (SpO_2_ <90% on room air) The severity of disease was defined as outlined in the Clinical Management Guidelines for COVID-19 by the Ministry of National Health Services, Pakistan. Within each of these two groups, patients were randomized (by lottery method) into treatment and control groups. Stratification was done via SAS software version 9.4 for age groups, gender, baseline severity of symptoms and co-morbidities to ensure that groups remain balanced in size for either arm. We performed allocation concealment with an interactive voice/web-based response system until randomization was finished on the system through a computer or phone. Care providers and outcome assessors were blinded with site investigators’ help to provide placebo or therapeutic regimen to the participants and masked clinicians to assess the clinical, laboratory and radiological findings. Data analysts were blinded by using statistical analysts from other institutions that did not have any conflict of research interest. All data were recorded on paper case record forms and then double entered into an electronic database and validated by trial staff.

### PROCEDURE

The HNS group received honey (1 gm) plus encapsulated *Nigella sativa* seeds (80 mg) per kg body weight orally in 2-3 divided doses daily for up-to 13 days while the control group received placebo (empty capsules). Both the products were certified for purity by botany department of Government College University, Lahore, Pakistan. Additionally, each patient in the trial received standard care therapy (SCT) as recommended by the treating physician and the clinical management guidelines for COVID-19 established by the Ministry of National Health Services of Pakistan. SCT primarily comprised of antipyretics, antibiotics, anticoagulants, steroids, supplemental oxygen and mechanical ventilation. The study participants were assessed for clinical symptoms daily by an on-site investigator for 13 days. During the study, when a patient recovered and remained asymptomatic for 48 h, he/she underwent a second SARS-CoV-2 RT-PCR test within the next 48 h (Figure 1). If the patient tested negative, they were deemed to have cleared the infection and their treatment stopped. In case of a positive test, a third PCR test was performed on day 14 with no further follow-up. A clinical grading score (CGS) was recorded for each patient on day 0, 4, 6, 8, 10 and 12. It was based on a seven-point ordinal scale: grade 1 (not hospitalized, no evidence of infection and resumption of normal activities), grade 2 (not hospitalized, but unable to resume normal activities), grade 3 (hospitalized, not requiring supplemental oxygen), grade 4 (hospitalized, requiring supplemental oxygen), grade 5 (hospitalized, requiring nasal high-flow oxygen therapy and/or noninvasive mechanical ventilation), grade 6 (hospitalized, requiring ECMO and/or invasive mechanical ventilation) and grade 7 (death). This scale has previously been used as end point in clinical trials in COVID-19 patients.^2^ Severity of symptoms was categorized as mild disease including patients with mild symptoms of COVID-19 but no radiological evidence of pneumonia. Moderate disease included patients with hypoxia (oxygen saturation <94% but >90%) or chest X-ray with infiltrates involving <50% of the lung fields or fever, cough, sputum production, and other respiratory tract related symptoms. Severe disease was defined as the presence of SaO_2_/SpO_2_ below 90% on room air or a PaO_2_ to FiO_2_ ratio of 300 or lower or radiological evidence showing more than 50% lungs involvement. Body temperature was measured, and fever was graded as no fever (98-99 °F), mild (>99-<100 °F), moderate (100-101.9 °F) and severe (≤102°F). The cough was classified as mild (occasional but transient cough), moderate (frequent but slightly influencing daytime activities) and severe (frequent cough but significantly influencing daytime activities). The shortness of breath was categorized as Grade1 (not troubled by breathlessness except on strenuous exercise), grade 2 (Short of breath when hurrying on the level or walking up a slight hill), grade 3 (walks slower than most people on the level, stops after a mile or so, or stop after 15 minutes walking at own pace), grade 4 (tops for breath after walking about 100 yards or a few minutes on level ground) and grade 5 (too breathless to leave the house, or breathless when undressing). Myalgia and “how sick do you feel” were subjective feeling assessed on 10-point chart and classified as mild, moderate and severe. Serum C-reactive Protein (CRP) levels were measured by ELISA kit (Invitrogen, USA). Safety outcomes including adverse events were categorized according to the National Cancer Institute Common Terminology Criteria for Adverse Events, version 4.0. Laboratory investigations were assessed as a part of the protocol as per recommendations of the treating physician. The trial steering committee monitored trial safety. For patients who were discharged before day 13 or were home-quarantined, follow-up was done by telemedicine.

**Figure 1:**
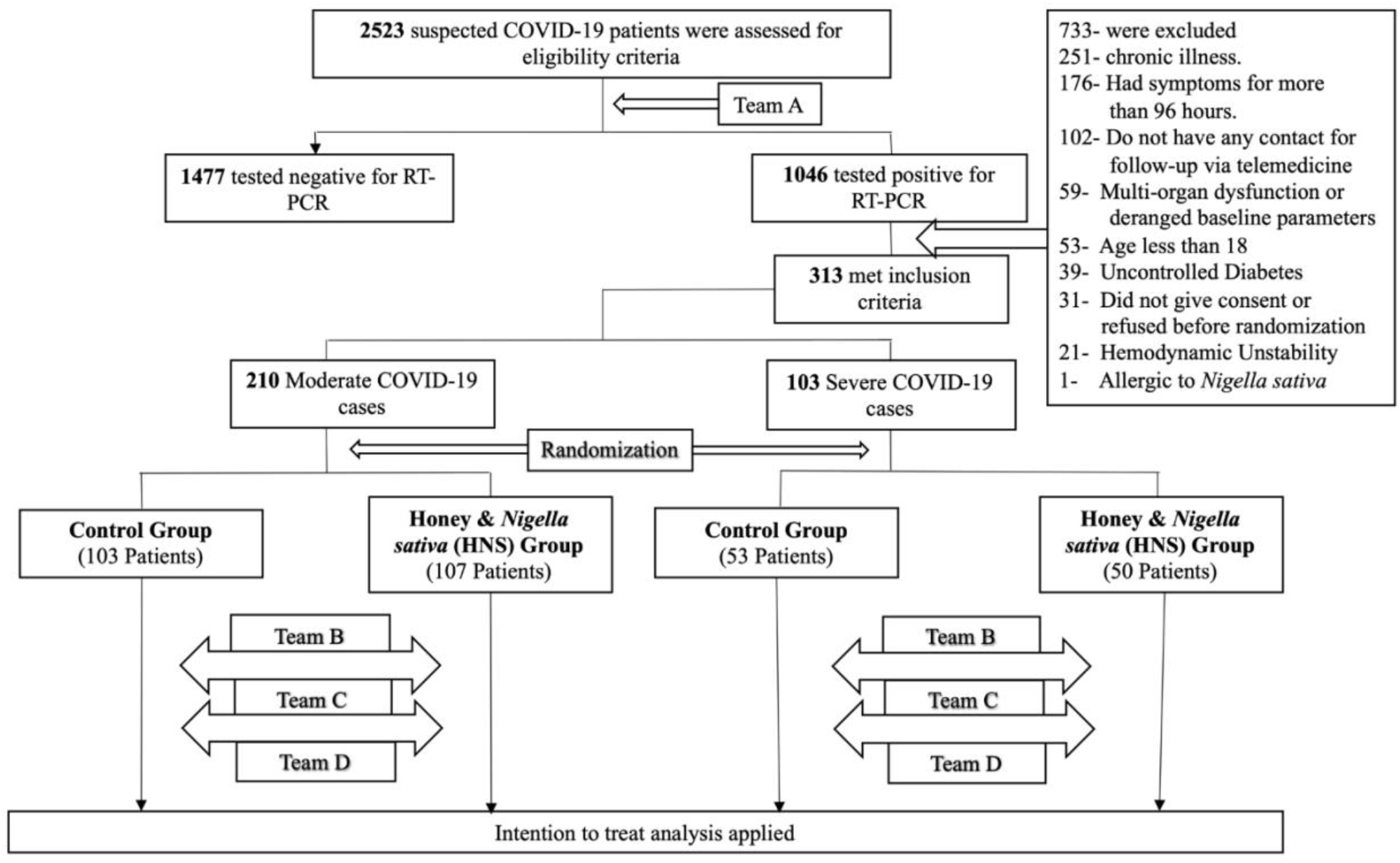
Study Flow Chart. Team A: Responsible for the recruitment and SARS-CoV-2 RT-PCR testing; Team B: Daily evaluated degree of fever, cough, myalgia, shortness of breath, oxygen therapy, “how sick do you feel” and rate emotional status; Team C: Reported clinical grading scale (CGS) on seven-point ordinal scale assessment as 0, 4, 6, 8, 10 and 12 days; Team D conducted follow-up PCR and CRP determinations.

### OUTCOMES

The primary outcomes were viral clearance (negative RT-PCR for the SARS-CoV-2 RNA), alleviation of clinical symptoms and CGS lowering on day 6. Secondary outcomes included reduction in fever degree (day 4), CRP levels (day 6), the severity of symptoms (day 8), CGS score (day 10) and mortality on day 30. Additional outcomes included median time to clinical improvement of severity of symptoms, degree of fever, cough, shortness of breath, myalgia and “how sick do you feel”. Median time to clinical improvement was assessed as one or two categories betterment or achievement of normal status on ordinal scale of clinical finding.

### STATISTICAL ANALYSIS

The sample size was not estimated when the trial was being planned since it was the first trial of its kind and the number of enrolled patients was dependent on the patient load in the clinical facilities, though 3 months’ time period was pre-specified. The efficacy was assessed on an intention to treat analysis of the randomly assigned groups in moderate and severe cases. The independent data monitoring committee reviewed unblinded analyses of the study data and any relevant information on monthly basis. In univariate analyses, we used a log-rank test to compare time taken for viral clearance, alleviation of symptoms, time to improvement in the severity of clinical symptoms, degree of fever, cough, shortness of breath, myalgia and “how sick do you feel”. Kaplan Meier method was applied to estimate survival curves for time for alleviation of symptoms and viral clearance. The Fisher’s Exact test was used to compare 30-day mortality. In the multivariate analyses, we used multivariate regression models to adjust for the effects of age (<40 or >=40), gender, baseline clinical status grade, history of diabetes/hypertension, and oxygen use. In the multivariate analyses of ordinal outcomes, we used ordinal logistic regression models assuming proportional odds. We also used a linear regression model to analyze the continuous outcome CRP and Cox proportional hazards models to analyze time to symptom alleviation and the time to viral clearance. SAS software version 9.4 (SAS Institute Inc., Cary, NC) was used for these analyses. This trial is registered with ClinicalTrials.gov, **NCT04347382**.

## RESULTS

Two thousand five hundred and twenty-three suspected COVID-19 patients were screened from April 30, to July 29, 2020, in which 1046 patients tested positive for the SARS-CoV-2 nucleic acid. Of these participants, 313 met the inclusion criteria (Figure 1). The spectrum of their clinical symptoms was stratified into two groups: moderate and severe. The two groups comprised of 210 and 103 patients, respectively. The patients within each of the two groups were randomly assigned to the treatment and control groups. The number of patients in moderate control, moderate HNS, severe control and severe HNS were 103, 107, 53, and 50, respectively. Their baseline demographics with clinical and laboratory parameters are shown in Table 1. Paracetamol and azithromycin were the top two prescribed drugs as part of the SCT. Two patients opted for home quarantine despite needing oxygen therapy.

**Table 1.** Baseline characteristics of study participants*.

| Table 1. Baseline characteristics of study participants* |  |  |  |  |
| --- | --- | --- | --- | --- |
| Parameter | Total<br>(n=313) | Control<br>(n=156) | Honey-Nigella<br>Sativa (n=157) | P-Value<sup>f</sup> |
| Age (Years) |  |  |  |  |
| ≤40 | 156 (49.84) | 80 (51.28) | 76 (48.4) | 0.48 |
| 40-59 | 93 (29.71) | 45 (28.85) | 48 (30.57) |  |
| 60-79 | 52 (16.61) | 26 (16.67) | 26 (16.56) |  |
| ≥80 | 12 (3.83) | 5 (3.2) | 7 (4.45) |  |
| Sex |  |  |  |  |
| Male | 178 (56.87) | 88 (56.41) | 90 (57.32) | 0.87 |
| Female | 135 (43.13) | 68 (43.59) | 67 (42.68) |  |
| Profession |  |  |  |  |
| Health care<sup>g</sup> | 71 (22.68) | 38 (24.36) | 33 (21.02) | 0.48 |
| Non-Health care | 242 (77.32) | 118 (75.64) | 124 (78.98) |  |
| Co-Morbidities |  |  |  |  |
| Hypertension | 99 (31.63) | 51 (32.69) | 48 (30.57) | 0.69 |
| Diabetes Mellitus | 115 (36.74) | 60 (38.46) | 55 (35.03) | 0.53 |
| Onset of symptoms before admission |  |  |  |  |
| 48 hours | 88 (38.1) | 49 (41.53) | 39 (34.51) | 0.22 |
| 72 hours | 143 (61.9) | 69 (58.47) | 74 (65.49) |  |
| 96 hours | 82 (36.44) | 38 (35.51) | 44 (37.29) |  |
| Severity of Symptoms |  |  |  |  |
| Moderate | 210 (67.09) | 103 (66.03) | 107 (68.15) | 0.69 |
| Severe | 103 (32.91) | 53 (33.97) | 50 (31.85) |  |
| ARDS | 57 (17.38) | 28 (17.95) | 29 (16.86) | 0.9 |
| Chest X-Ray |  |  |  |  |
| Normal | 217 (66.16) | 101 (64.74) | 116 (73.88) | 0.71 |
| Pneumonic Patch | 12 (3.66) | 8 (5.13) | 4 (2.54) |  |
| Unilateral Infiltrates | 40 (12.2) | 19 (12.18) | 21 (13.38) |  |
| Bilateral Infiltrates | 59 (17.99) | 28 (17.94) | 31 (19.74) |  |
| Clinical Grading Score at day 0 |  |  |  |  |
| Median Grade Score (IQR) | 3 (2-4) | 3 (2-4) | 3 (2-4) | 0.73 |
| 2- Not hospitalized with unable to resume normal activities | 139 (44.41) | 68 (43.59) | 71 (45.22) |  |
| 3- Hospitalized, not requiring supplemental oxygen | 71 (22.68) | 35 (22.44) | 36 (22.93) |  |
| 4- Hospitalized, requiring low flow supplemental oxygen | 44 (14.06) | 23 (14.74) | 21 (13.38) |  |
| 5- Hospitalized, requiring high flow supplemental oxygen | 59 (18.85) | 30 (19.23) | 29 (18.47) |  |
| Patients hospitalized in |  |  |  |  |
| Shaikh Zayed Hospital | 78 (25.66) | 39 (25.83) | 39 (25.49) | 0.56 |
| Services Institute of Medical Sciences | 91 (29.93) | 48 (31.79) | 43 (28.1) |  |
| Doctors Lounge | 52 (17.11) | 27 (17.88) | 25 (16.34) |  |
| Ali Clinic | 83 (27.3) | 37 (24.5) | 46 (30.07) |  |

| Patients showing symptoms |  |  |  |  |
| --- | --- | --- | --- | --- |
| Fever | 303 (96.81) | 152 (97.44) | 151 (96.17) | 0.53 |
| SOB | 106 (33.87) | 56 (35.9) | 50 (31.85) | 0.45 |
| Cough | 192 (61.34) | 90 (57.69) | 102 (64.97) | 0.19 |
| Myalgia | 169 (53.99) | 89 (57.05) | 80 (50.96) | 0.28 |
| Patients receiving<sup>‡</sup> |  |  |  |  |
| Panadol | 297 (94.89) | 147 (94.23) | 150 (97.54) | 0.6 |
| Azithromycin | 231 (73.8) | 120 (76.92) | 111 (70.7) | 0.21 |
| Montelukast | 106 (33.87) | 56 (35.9) | 50 (31.85) | 0.45 |
| Supplemental Oxygen | 105 (33.55) | 55 (35.25) | 50 (31.85) | 0.52 |
| Low Molecular Weight Heparin | 72 (23) | 38 (24.36) | 34 (21.66) | 0.57 |
| Hydrocortisone | 83 (26.52) | 45 (28.85) | 38 (24.2) | 0.35 |
| Multivitamins | 147 (46.96) | 73 (46.8) | 74 (47.13) | 0.95 |
| Tanzobactam + Piperacillin | 73 (23.32) | 42 (26.92) | 31 (19.74) | 0.13 |
| Ivermectin | 114 (36.42) | 60 (38.46) | 54 (34.39) | 0.45 |
| Meropenem | 62 (19.81) | 35 (22.43) | 27 (17.2) | 0.25 |

\* Data are presented as no. (%) unless indicated. The Intention-to-Treat analysis was performed on all the patients who had undergone randomization. ECMO: Extracorporeal membrane oxygenation; CRP: C-reactive protein; AST: Aspartate transaminase; ALT: Alanine transaminase; ECG: Electrocardiography; ARDS: Acute respiratory distress syndrome; SOB: Shortness of breath.
‡ P ≤ 0.05 was determined significant
¶ Medical doctors, nurses and pharmacists.
‡ These medications were part of standard care therapy as per decision of treating physician and clinical Management Guidelines for COVID-19 by Ministry of National Health Services, Pakistan.

### PRIMARY OUTCOMES

Primary outcomes are shown in Table 2. Alleviation of COVID-19 symptoms for patients in the HNS groups occurred earlier than control groups: 4 versus 7 days for the moderate patients (HR: 6.11; 95% CI: 4.23-8.84; P<0.0001) and 6 versus 13 days for the severe disease patients (HR: 4.04; 95% CI: 2.46-6.64; P<0.0001). Viral clearance (being negative for the SARS-CoV-2 RT-PCR test) occurred 4 days sooner in the HNS group for both moderate (HR: 5.53; 95% CI: 3.76-8.14; P<0.0001) and severe cases (HR: 4.32; 95% CI: 2.62-7.13; P<0.0001). The Kaplan-Meier curves for these variables are shown in Figure 2. In moderate patients, the HNS group resumed earlier while control group was unable to resume daily life activities as evident by the lower median CGS at day 6 (odds ratio: 0.07; 95% CI: 0.03-0.13; P<0.0001). Meanwhile, in severe groups, the HNS cases were discharged from the hospital, whereas the control cases were still hospitalized on supplemental oxygen as per median CGS at day 6 (Odds Ratio: 0.03; 95% CI: 0.01-0.09; P<0.0001).

**Table 2.**
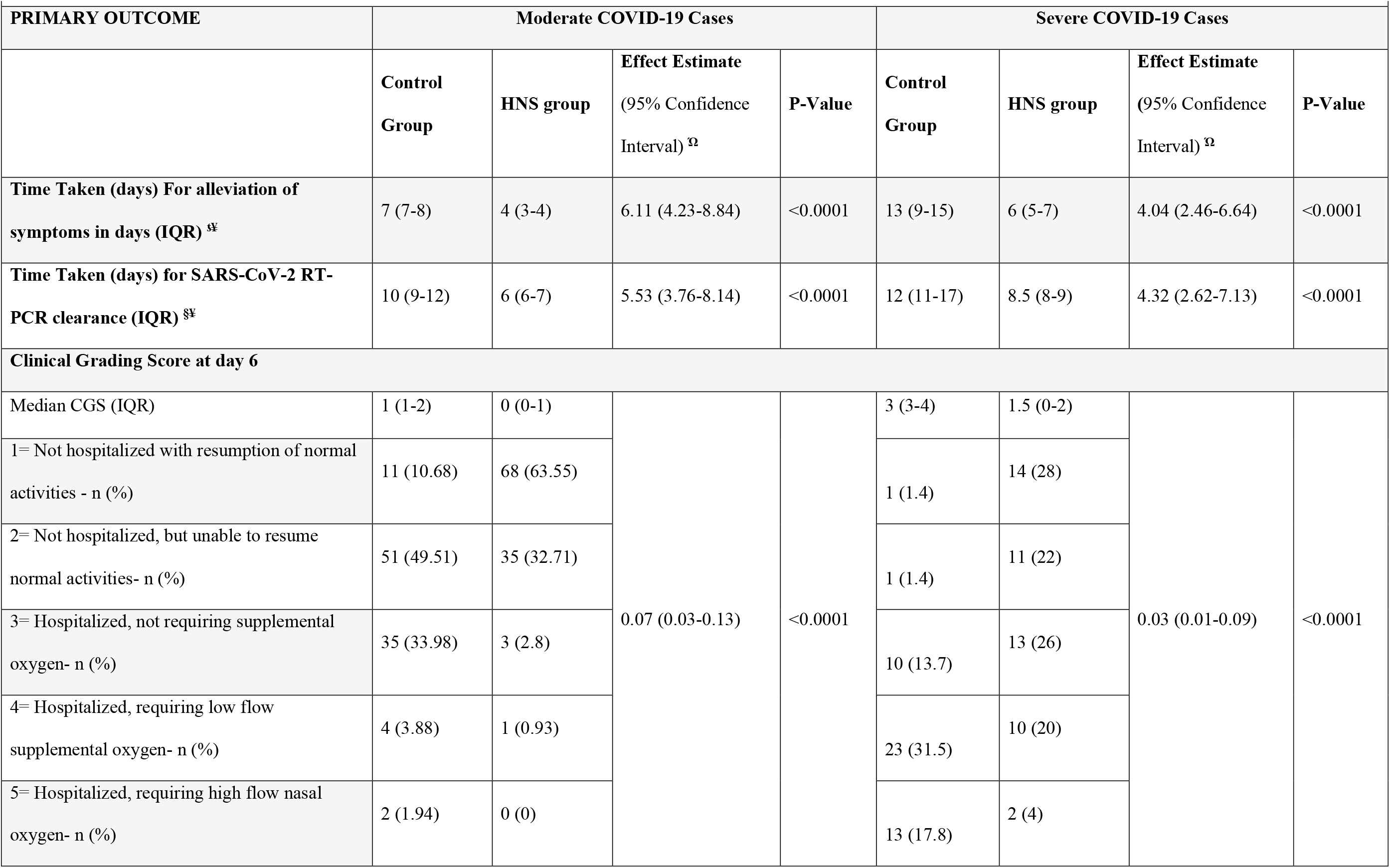
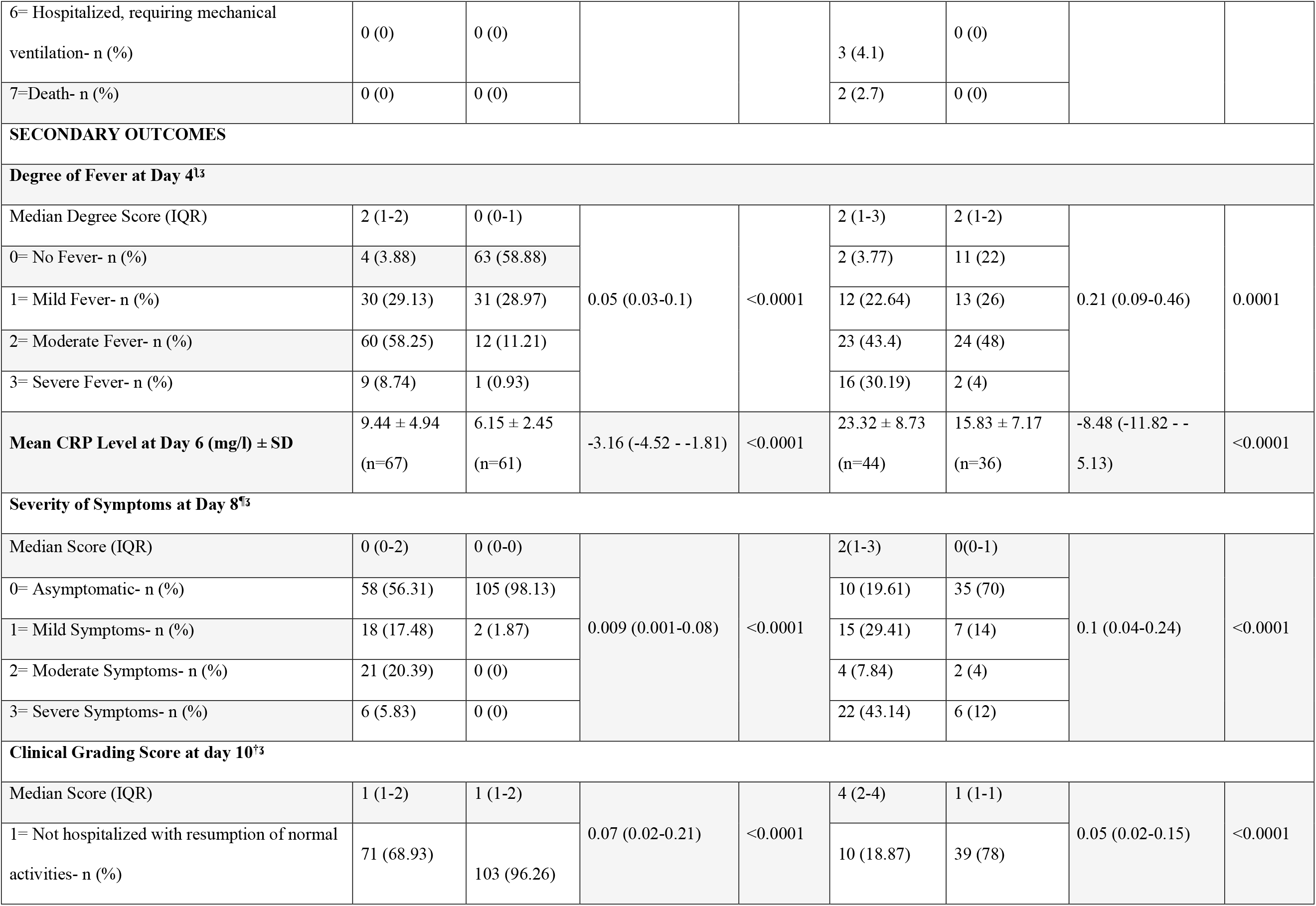
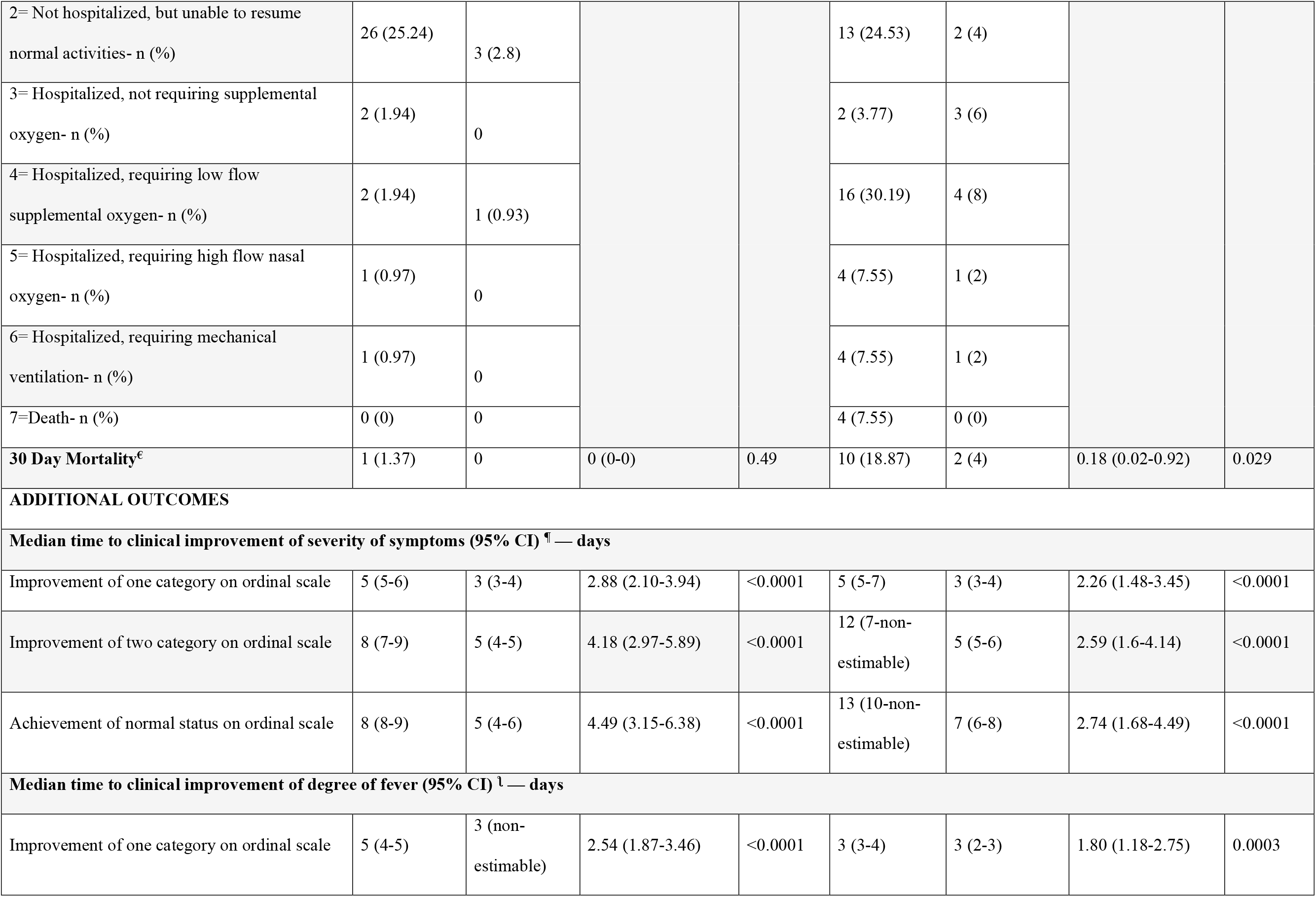
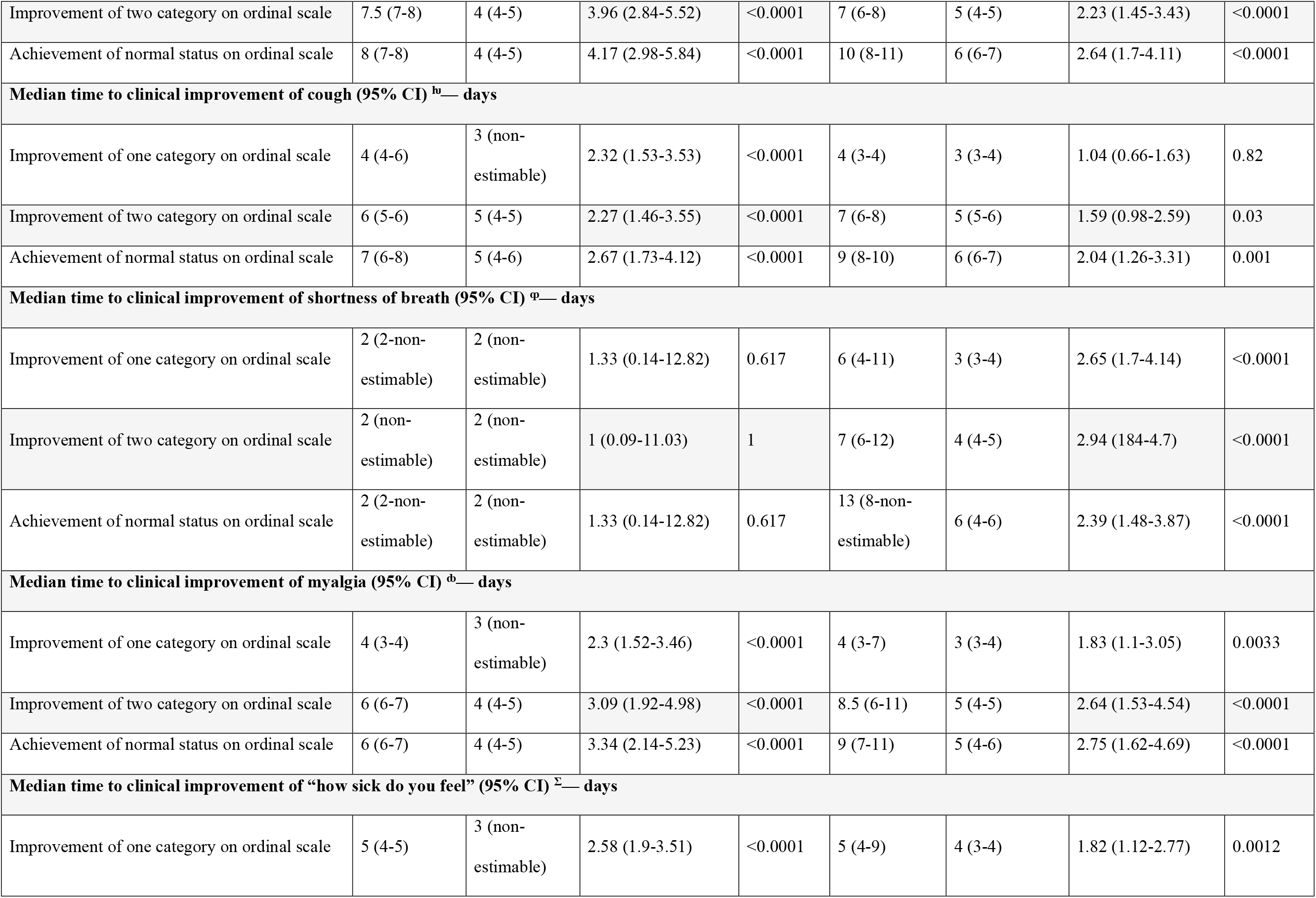
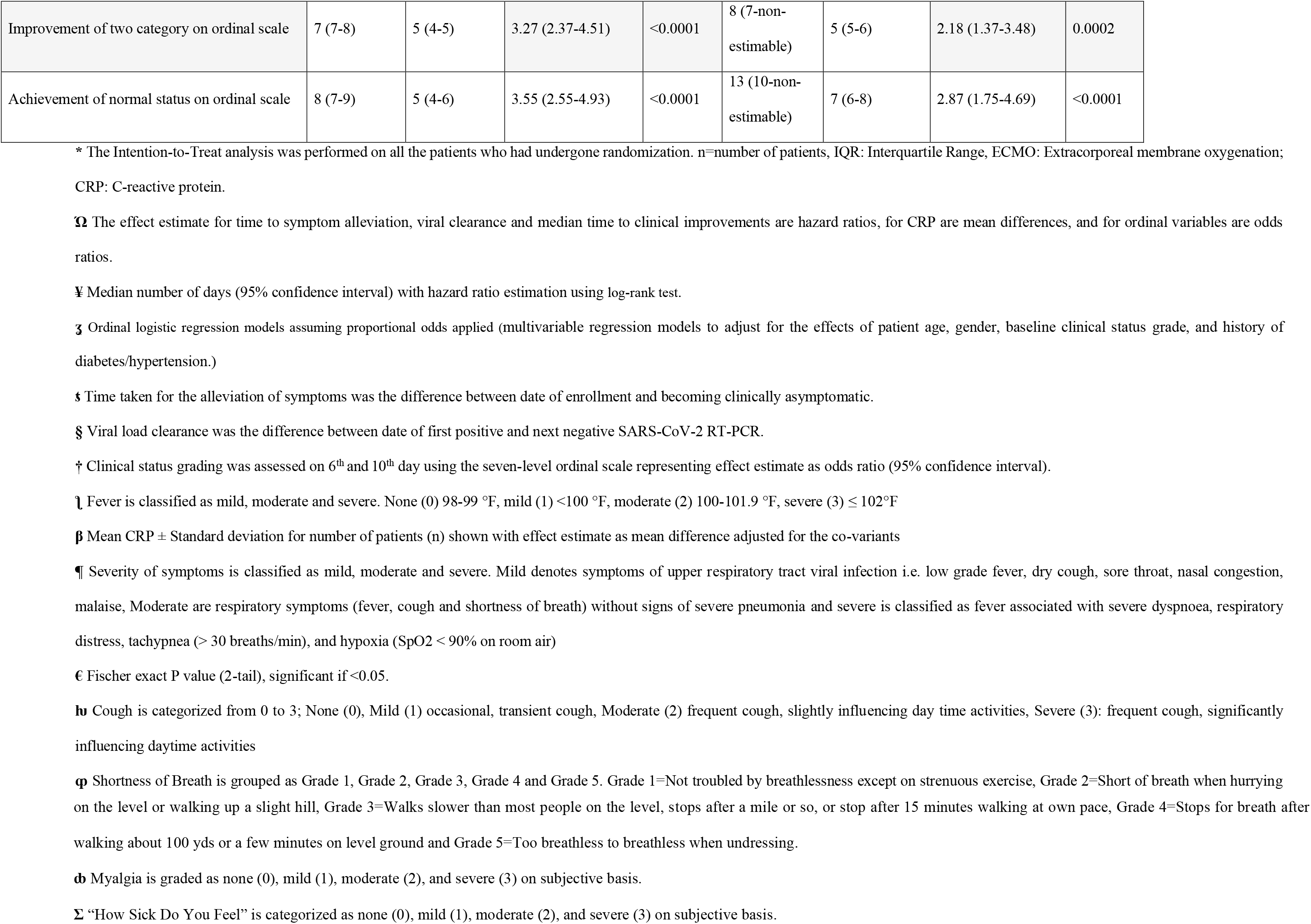
Primary and Secondary Outcome (Intention-to-Treat Population) *.

| Table 2. Primary and Secondary Outcome (Intention-to-Treat Population) * |  |  |  |  |  |  |  |  |
| --- | --- | --- | --- | --- | --- | --- | --- | --- |
| PRIMARY OUTCOME | Moderate COVID-19 Cases |  |  |  | Severe COVID-19 Cases |  |  |  |
|  | Control Group | HNS group | Effect Estimate<br>(95% Confidence Interval) <sup>Ω</sup> | P-Value | Control Group | HNS group | Effect Estimate<br>(95% Confidence Interval) <sup>Ω</sup> | P-Value |
| Time Taken (days) For alleviation of symptoms in days (IQR) <sup>§¥</sup> | 7 (7-8) | 4 (3-4) | 6.11 (4.23-8.84) | <0.0001 | 13 (9-15) | 6 (5-7) | 4.04 (2.46-6.64) | <0.0001 |
| Time Taken (days) for SARS-CoV-2 RT-PCR clearance (IQR) <sup>§¥</sup> | 10 (9-12) | 6 (6-7) | 5.53 (3.76-8.14) | <0.0001 | 12 (11-17) | 8.5 (8-9) | 4.32 (2.62-7.13) | <0.0001 |
| Clinical Grading Score at day 6 |  |  |  |  |  |  |  |  |
| Median CGS (IQR) | 1 (1-2) | 0 (0-1) | 0.07 (0.03-0.13) | <0.0001 | 3 (3-4) | 1.5 (0-2) | 0.03 (0.01-0.09) | <0.0001 |
| 1= Not hospitalized with resumption of normal activities - n (%) | 11 (10.68) | 68 (63.55) |  |  | 1 (1.4) | 14 (28) |  |  |
| 2= Not hospitalized, but unable to resume normal activities- n (%) | 51 (49.51) | 35 (32.71) |  |  | 1 (1.4) | 11 (22) |  |  |
| 3= Hospitalized, not requiring supplemental oxygen- n (%) | 35 (33.98) | 3 (2.8) |  |  | 10 (13.7) | 13 (26) |  |  |
| 4= Hospitalized, requiring low flow supplemental oxygen- n (%) | 4 (3.88) | 1 (0.93) |  |  | 23 (31.5) | 10 (20) |  |  |
| 5= Hospitalized, requiring high flow nasal oxygen- n (%) | 2 (1.94) | 0 (0) |  |  | 13 (17.8) | 2 (4) |  |  |
| 6= Hospitalized, requiring mechanical ventilation- n (%) | 0 (0) | 0 (0) |  |  | 3 (4.1) | 0 (0) |  |  |
| 7=Death- n (%) | 0 (0) | 0 (0) |  |  | 2 (2.7) | 0 (0) |  |  |
| SECONDARY OUTCOMES |  |  |  |  |  |  |  |  |
| Degree of Fever at Day 4<sup>13</sup> |  |  |  |  |  |  |  |  |
| Median Degree Score (IQR) | 2 (1-2) | 0 (0-1) | 0.05 (0.03-0.1) | <0.0001 | 2 (1-3) | 2 (1-2) | 0.21 (0.09-0.46) | 0.0001 |
| 0= No Fever- n (%) | 4 (3.88) | 63 (58.88) |  |  | 2 (3.77) | 11 (22) |  |  |
| 1= Mild Fever- n (%) | 30 (29.13) | 31 (28.97) |  |  | 12 (22.64) | 13 (26) |  |  |
| 2= Moderate Fever- n (%) | 60 (58.25) | 12 (11.21) |  |  | 23 (43.4) | 24 (48) |  |  |
| 3= Severe Fever- n (%) | 9 (8.74) | 1 (0.93) |  |  | 16 (30.19) | 2 (4) |  |  |
| Mean CRP Level at Day 6 (mg/l) ± SD | 9.44 ± 4.94<br>(n=67) | 6.15 ± 2.45<br>(n=61) | -3.16 (-4.52 - -1.81) | <0.0001 | 23.32 ± 8.73<br>(n=44) | 15.83 ± 7.17<br>(n=36) | -8.48 (-11.82 - -5.13) | <0.0001 |
| Severity of Symptoms at Day 8<sup>13</sup> |  |  |  |  |  |  |  |  |
| Median Score (IQR) | 0 (0-2) | 0 (0-0) | 0.009 (0.001-0.08) | <0.0001 | 2(1-3) | 0(0-1) | 0.1 (0.04-0.24) | <0.0001 |
| 0= Asymptomatic- n (%) | 58 (56.31) | 105 (98.13) |  |  | 10 (19.61) | 35 (70) |  |  |
| 1= Mild Symptoms- n (%) | 18 (17.48) | 2 (1.87) |  |  | 15 (29.41) | 7 (14) |  |  |
| 2= Moderate Symptoms- n (%) | 21 (20.39) | 0 (0) |  |  | 4 (7.84) | 2 (4) |  |  |
| 3= Severe Symptoms- n (%) | 6 (5.83) | 0 (0) |  |  | 22 (43.14) | 6 (12) |  |  |
| Clinical Grading Score at day 10<sup>†3</sup> |  |  |  |  |  |  |  |  |
| Median Score (IQR) | 1 (1-2) | 1 (1-2) | 0.07 (0.02-0.21) | <0.0001 | 4 (2-4) | 1 (1-1) | 0.05 (0.02-0.15) | <0.0001 |
| 1= Not hospitalized with resumption of normal activities- n (%) | 71 (68.93) | 103 (96.26) |  |  | 10 (18.87) | 39 (78) |  |  |
| 2= Not hospitalized, but unable to resume normal activities- n (%) | 26 (25.24) | 3 (2.8) |  |  | 13 (24.53) | 2 (4) |  |  |
| 3= Hospitalized, not requiring supplemental oxygen- n (%) | 2 (1.94) | 0 |  |  | 2 (3.77) | 3 (6) |  |  |
| 4= Hospitalized, requiring low flow supplemental oxygen- n (%) | 2 (1.94) | 1 (0.93) |  |  | 16 (30.19) | 4 (8) |  |  |
| 5= Hospitalized, requiring high flow nasal oxygen- n (%) | 1 (0.97) | 0 |  |  | 4 (7.55) | 1 (2) |  |  |
| 6= Hospitalized, requiring mechanical ventilation- n (%) | 1 (0.97) | 0 |  |  | 4 (7.55) | 1 (2) |  |  |
| 7=Death- n (%) | 0 (0) | 0 |  |  | 4 (7.55) | 0 (0) |  |  |
| **30 Day Mortality<sup>€</sup>** | 1 (1.37) | 0 | 0 (0-0) | 0.49 | 10 (18.87) | 2 (4) | 0.18 (0.02-0.92) | 0.029 |
| **ADDITIONAL OUTCOMES** |  |  |  |  |  |  |  |  |
| **Median time to clinical improvement of severity of symptoms (95% CI) <sup>¶</sup> — days** |  |  |  |  |  |  |  |  |
| Improvement of one category on ordinal scale | 5 (5-6) | 3 (3-4) | 2.88 (2.10-3.94) | <0.0001 | 5 (5-7) | 3 (3-4) | 2.26 (1.48-3.45) | <0.0001 |
| Improvement of two category on ordinal scale | 8 (7-9) | 5 (4-5) | 4.18 (2.97-5.89) | <0.0001 | 12 (7-non-estimable) | 5 (5-6) | 2.59 (1.6-4.14) | <0.0001 |
| Achievement of normal status on ordinal scale | 8 (8-9) | 5 (4-6) | 4.49 (3.15-6.38) | <0.0001 | 13 (10-non-estimable) | 7 (6-8) | 2.74 (1.68-4.49) | <0.0001 |
| **Median time to clinical improvement of degree of fever (95% CI) <sup>‡</sup> — days** |  |  |  |  |  |  |  |  |
| Improvement of one category on ordinal scale | 5 (4-5) | 3 (non-estimable) | 2.54 (1.87-3.46) | <0.0001 | 3 (3-4) | 3 (2-3) | 1.80 (1.18-2.75) | 0.0003 |
| Improvement of two category on ordinal scale | 7.5 (7-8) | 4 (4-5) | 3.96 (2.84-5.52) | <0.0001 | 7 (6-8) | 5 (4-5) | 2.23 (1.45-3.43) | <0.0001 |
| Achievement of normal status on ordinal scale | 8 (7-8) | 4 (4-5) | 4.17 (2.98-5.84) | <0.0001 | 10 (8-11) | 6 (6-7) | 2.64 (1.7-4.11) | <0.0001 |
| **Median time to clinical improvement of cough (95% CI) <sup>h</sup>— days** |  |  |  |  |  |  |  |  |
| Improvement of one category on ordinal scale | 4 (4-6) | 3 (non-estimable) | 2.32 (1.53-3.53) | <0.0001 | 4 (3-4) | 3 (3-4) | 1.04 (0.66-1.63) | 0.82 |
| Improvement of two category on ordinal scale | 6 (5-6) | 5 (4-5) | 2.27 (1.46-3.55) | <0.0001 | 7 (6-8) | 5 (5-6) | 1.59 (0.98-2.59) | 0.03 |
| Achievement of normal status on ordinal scale | 7 (6-8) | 5 (4-6) | 2.67 (1.73-4.12) | <0.0001 | 9 (8-10) | 6 (6-7) | 2.04 (1.26-3.31) | 0.001 |
| **Median time to clinical improvement of shortness of breath (95% CI) <sup>o</sup>— days** |  |  |  |  |  |  |  |  |
| Improvement of one category on ordinal scale | 2 (2-non-estimable) | 2 (non-estimable) | 1.33 (0.14-12.82) | 0.617 | 6 (4-11) | 3 (3-4) | 2.65 (1.7-4.14) | <0.0001 |
| Improvement of two category on ordinal scale | 2 (non-estimable) | 2 (non-estimable) | 1 (0.09-11.03) | 1 | 7 (6-12) | 4 (4-5) | 2.94 (1.84-4.7) | <0.0001 |
| Achievement of normal status on ordinal scale | 2 (2-non-estimable) | 2 (non-estimable) | 1.33 (0.14-12.82) | 0.617 | 13 (8-non-estimable) | 6 (4-6) | 2.39 (1.48-3.87) | <0.0001 |
| **Median time to clinical improvement of myalgia (95% CI) <sup>o</sup>— days** |  |  |  |  |  |  |  |  |
| Improvement of one category on ordinal scale | 4 (3-4) | 3 (non-estimable) | 2.3 (1.52-3.46) | <0.0001 | 4 (3-7) | 3 (3-4) | 1.83 (1.1-3.05) | 0.0033 |
| Improvement of two category on ordinal scale | 6 (6-7) | 4 (4-5) | 3.09 (1.92-4.98) | <0.0001 | 8.5 (6-11) | 5 (4-5) | 2.64 (1.53-4.54) | <0.0001 |
| Achievement of normal status on ordinal scale | 6 (6-7) | 4 (4-5) | 3.34 (2.14-5.23) | <0.0001 | 9 (7-11) | 5 (4-6) | 2.75 (1.62-4.69) | <0.0001 |
| **Median time to clinical improvement of “how sick do you feel” (95% CI) <sup>z</sup>— days** |  |  |  |  |  |  |  |  |
| Improvement of one category on ordinal scale | 5 (4-5) | 3 (non-estimable) | 2.58 (1.9-3.51) | <0.0001 | 5 (4-9) | 4 (3-4) | 1.82 (1.12-2.77) | 0.0012 |
| Improvement of two category on ordinal scale | 7 (7-8) | 5 (4-5) | 3.27 (2.37-4.51) | <0.0001 | 8 (7-non-estimable) | 5 (5-6) | 2.18 (1.37-3.48) | 0.0002 |
| Achievement of normal status on ordinal scale | 8 (7-9) | 5 (4-6) | 3.55 (2.55-4.93) | <0.0001 | 13 (10-non-estimable) | 7 (6-8) | 2.87 (1.75-4.69) | <0.0001 |

\* The Intention-to-Treat analysis was performed on all the patients who had undergone randomization. n=number of patients, IQR: Interquartile Range, ECMO: Extracorporeal membrane oxygenation; CRP: C-reactive protein.

**Figure 2.**
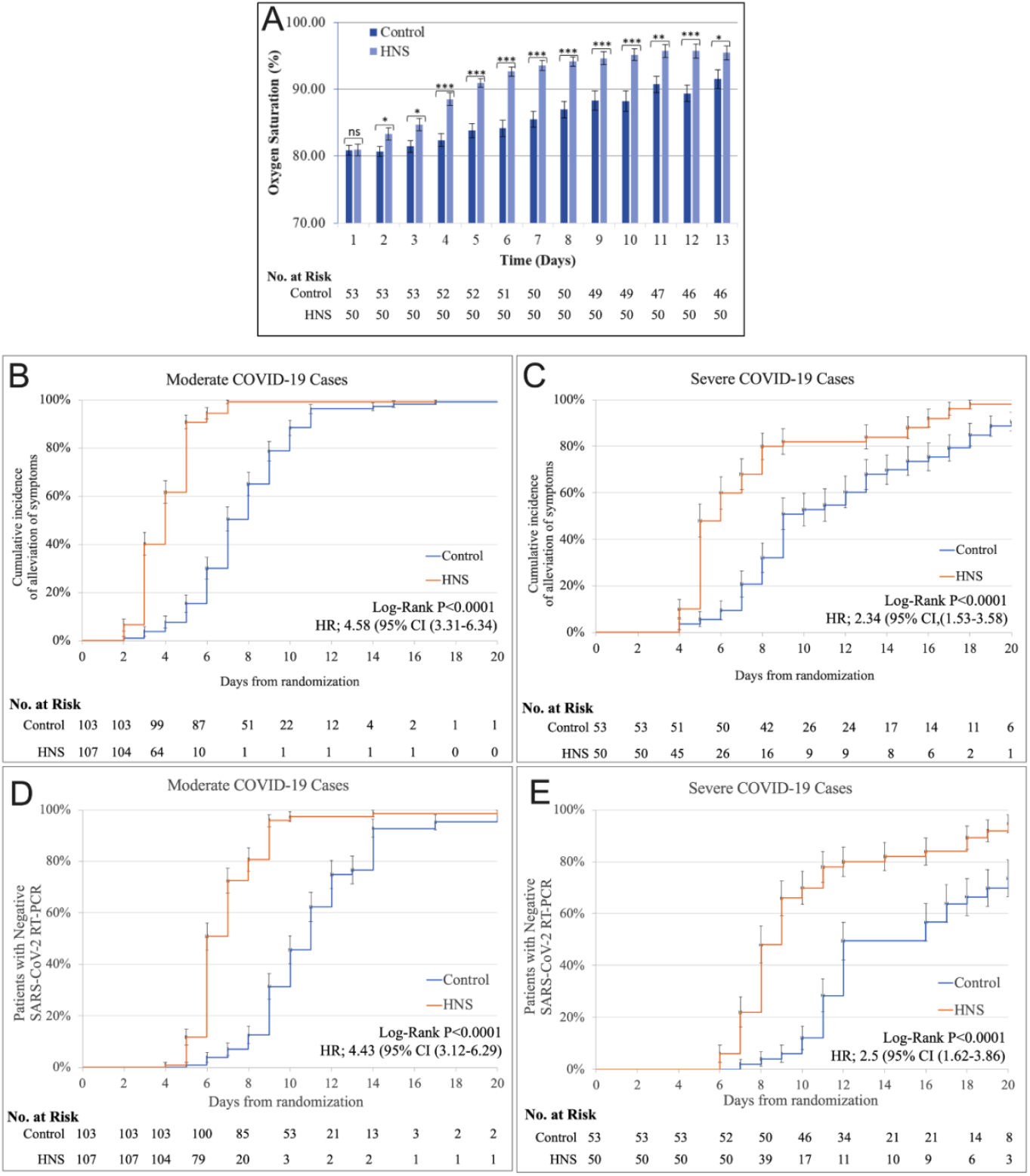
Kinetic changes in outcomes. A. Mean oxygen saturation spO2 over time in severe cases; Kaplan-Meier probability curves for time taken (in days) for alleviation of symptoms in moderate (B) and severe cases (C); Kaplan-Meier probability curves for time taken (in days) for vial clearance in moderate (D) and severe cases (E).ns = non-significant, *= P<0.05, **=P<0.001, ***=P<0.0001.

### SECONDARY OUTCOMES

There were significant differences in all secondary outcomes between the treatment and control groups (see Table 2 for secondary outcomes). In moderate COVID-19 patients, the degree of fever (median) was 100-101.9°F (moderate) in the control group while HNS arm participants were afebrile on day 4 (OR: 0.05; 95% CI: 0.03-0.1; P <0.0001). A significant reduction in the degree of fever was observed in severe cases on day 4 (OR: 0.21; 95% CI: 0.09-0.46; P=0.0001). CRP levels decreased significantly on day 6 in both the HNS groups compared with their respective control groups (moderate 6.15 ± 2.45 versus 9.44 ± 4.94 mg/L, P <0.0001 and severe cases 15.83 ± 7.17 versus 23.32 ± 8.73 mg/L, P <0.0001). On day 8, as per the median degree of symptoms severity, 98.13% patients were asymptomatic in HNS treated moderate cases compared to 56.31% in the control group (OR: 0.009; 95% CI: 0.001-0.08; P<0.0001). Similarly, in severe cases, more patients became asymptomatic in the HNS group in severe cases while a higher number had moderate symptoms (median) in the control arm (OR: 0.1; 95% CI: 0.04-0.24). By day 10, 96.26% of the moderate cases patients fully resumed normal activities with HNS compared to 68.93% in the control group (OR: 0.07; 95% CI: 0.02-0.21). For the severe group, the median CGS at day 10 revealed that mostly HNS group cases resumed normal activities while control patients were still hospitalized requiring oxygen therapy (OR:0.05; 95% CI: 0.02-0.15). The distribution of patients in the ordinal-scale categories over time is shown in Figure 3. Thirty-day mortality was 18.87% in the control group and 4% with HNS therapy (OR: 0.18 95% CI: 0.02-0.92).

**Figure 3.**
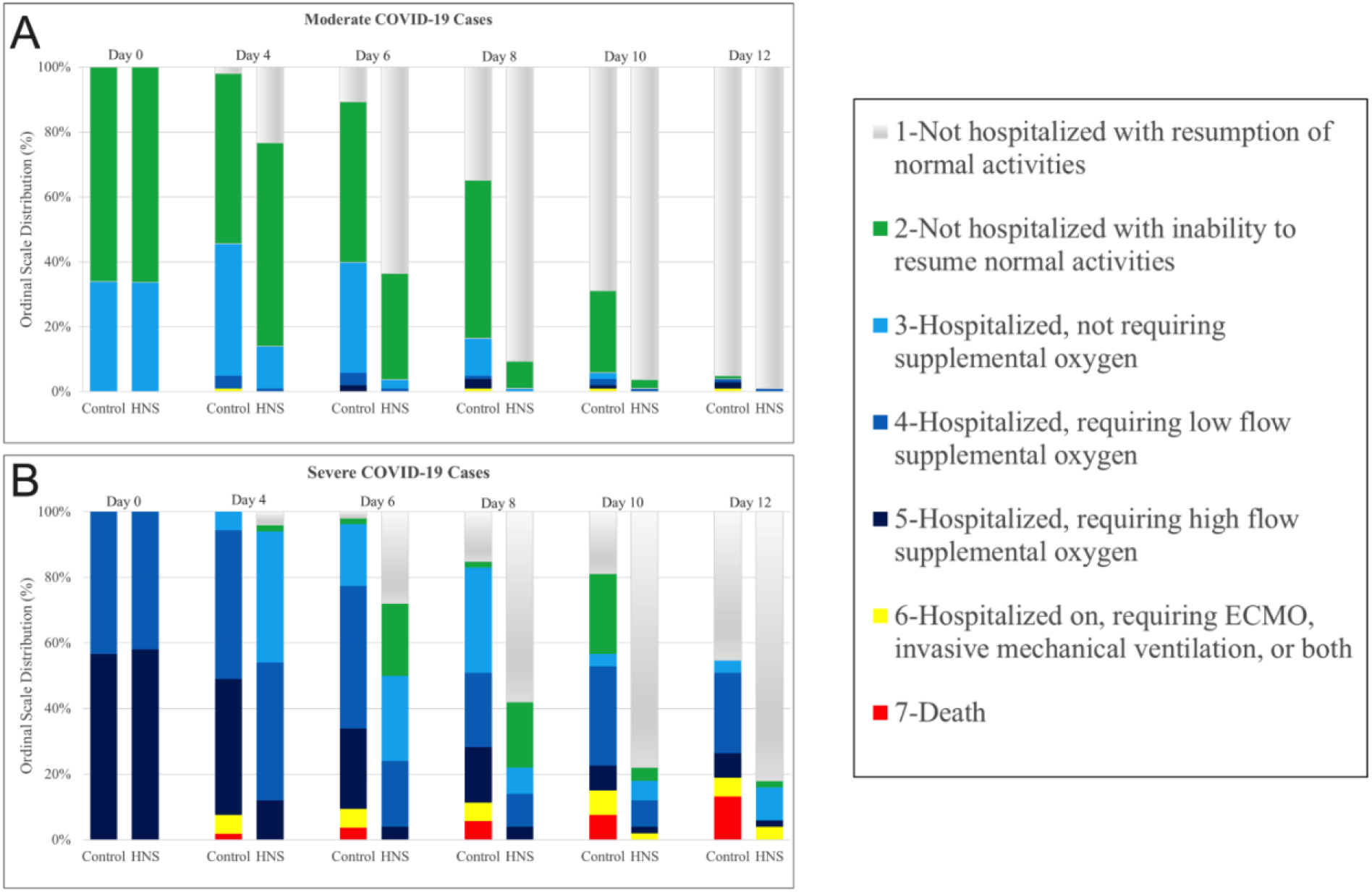
Kinetics of clinical status grading in Ordinal-Scale in COVID-19 patients. The Figure shows kinetic changes in clinical grade score (in 7-point ordinal-scale) in COVID-19 patient receiving the treatment (HNS) or placebo (Control). Note increases numbers of patients within scale 1 in the HNS group both for the moderate and severe cases.

### ADDITIONAL OUTCOMES

In HNS group, median day achievement of normal status on ordinal scale was earlier in severity of symptoms (moderate, 5 versus 8, HR; 4.49 (3.15-6.38), P<0.0001 and severe, 7 versus 13 HR; 2.74 (1.68-4.49), P<0.0001), degree of fever [4 versus 8, HR; 4.17 (2.98-5.84), P<0.0001 and severe 6 versus 10, HR; 2.64 (1.74-4.11), P<0.0001), degree of cough [moderate 5 versus 7, HR; 2.67 (1.73-4.12), P<0.0001 and severe 6 versus 9, HR; 2.04 (1.26-3.31), P=0.0001], degree of shortness of breath (severe 6 versus 13 HR; 2.39 (1.48-3.87), P<0.0001), degree of myalgia (moderate 4 versus 6 HR; 3.34 (2.14-5.25), P<0.0001) severe 5 versus 9, HR; 2.75 (1.62-4.69), P<0.0001) and “how sick do you feel” (moderate 5 versus 8 HR (3.55 (2.55-4.93), P<0.0001 and severe 7 versus 13 HR; 2.87 (1.75-4.69), P<0.0001) (Table 2). Similar results of earlier clinical improvement were seen with HNS group for severity of symptoms, degree of fever, cough, shortness of breath, myalgia and “how sick do you feel”. Furthermore, distribution on degree of fever, cough, myalgia, feeling of sickness, emotional status, shortness of breath, oxygen saturation, oxygen requirement and severity of symptoms over 13 days is given in supplementary Tables S2-S10. No evident adverse effects were noted with HNS.

## DISCUSSION

The study was a multicenter, randomized, placebo-controlled clinical trial investigating the therapeutic efficacy of HNS against COVID-19. To the best of our knowledge, this trial is the first of its’ kind in which a combination of two natural substances were investigated. Current study showed superior efficacy of HNS for COVID-19 in most of the studied outcomes. In control groups, about half of the patients required a 2-fold higher duration to become asymptomatic compared to those in the HNS groups. Similarly, in >50% of the participants, viral clearance occurred 4-5 days earlier in HNS groups, as tested by PCR. Mean oxygen saturation above 90%, in severe cases, was achieved 6 days earlier with HNS treatment (Figure 2). In the group of moderate cases, by day 4, more than half of participants (HNS group) became afebrile while control group patients persisted with a moderate fever. Furthermore, among severe cases 22% (versus 3.77%) of patients touched the baseline temperature. By day 6, among moderate cases, majority of the patients resumed normal daily activities in the HNS group compared to limited activities in the control group. Meanwhile, among severe cases, the majority of patients were discharged in HNS groups while control group participants were still hospitalized requiring oxygen therapy. On day 8, among moderate and severe cases, 98.13% (versus 56.31%) and 70% (versus 19.61%) became asymptomatic with HNS treatment, respectively. The mortality rate was ∽4 times higher in the control group. All data combined, is suggestive of a robust therapeutic profile of HNS and results can be highly encouraging amidst current second wave of SARS-CoV-2 infection. This superior efficacy of HNS is attributed to the combination of its several constituents.

Unlike other antivirals that target a specific structure or pathway of SARS-CoV-2 infection, HNS might kill/inhibit the virus through a multiprong strategy by targeting numerous viral sites or host-virus interactions. For example, in-silico and in-vitro studies predicted that multiple flavonoid and phenolic compounds present in HNS such as kaempferol, hesperidin’s, chrysin, quercetin and linoleic acid docked/bound to the SARS-CoV-2 spike protein–human Angiotensin Converting Enzyme-2 receptor (a receptor for spike protein binding of novel coronavirus) complex.^19,20^ Among the flavonoids in HNS, of particular interest is quercetin which is being investigated in many clinical trials against COVID-19. Other than quercetin’s predicted binding to the viral-human protein junction, it is also known to possess antibacterial properties by disrupting membranes, transport, and motility. HNS also contains Zinc, which has established antiviral properties against several viruses via inhibiting viral replication and acting as an immuno-modulatory agent.^21^ Till-date thirty-eight clinical trials are underway to test the efficacy of Zinc as an effective anti-COVID-19 agent. Another constituent of HNS is ascorbic acid (vitamin C), a common antioxidant and free radical scavenger with anti-inflammatory properties that reduces mediators such as IL-6 and endothelin-1. It also has proven antimicrobial and immunomodulatory properties and block several key components of cytokine storms.^22,23^ To this end, forty-five clinical trials have been registered using ascorbic acid to test its therapeutic benefit as prophylaxis and adjunctive medical therapy against COVID-19, thus far. Thus, HNS contains a cocktail of phytochemicals that complement one another to tackle SARS-CoV-2 related disease pathologies. For instance, quercetin is a zinc ionophore and their synergism with vitamin C against SARS-CoV-2 is suggested.^24^ Quercetin has also shown to inhibit pro-inflammatory cytokine responses (by reducing MHC class-II antigen presentation and TLR-signaling from activated dendritic cells) while stimulating T-helper 1 and cytotoxic-CD8 pathway for adequate viral clearance. Moreover, these processes are further enhanced by the presence of Zinc, thus potentiating each other in a coordinated fashion.^19^ In an in-vitro study, linoleic acid (flavonoid) has also shown synergy with the COVID-19 drug remdesivir, suppressing SARS-CoV-2 replication.^20^

Honey is mainly comprised of sugars with small amounts of amino acids, proteins, enzymes, organic acids, vitamins, minerals, volatile substances, and polyphenols. The antibacterial features of honey have been attributed to high sugar concentration, hydrogen peroxide (H_2_O_2_) and low pH along with methylglyoxal and the antimicrobial peptide bee defensin-1. The presence of H_2_O_2_ within honey irreversibly damages microbial DNA through the generation of hydroxyl radicals.^5^ Moreover, honey promotes lymphocyte proliferation, stimulates phagocytosis, and regulates the pro-inflammatory cytokine production. Some constituents (for instance, type II arabinogalactans, methylglyoxal, and the major royal jelly protein-1) cause immunostimulatory or pro-inflammatory action via stimulating the production of immunological mediators like tumor necrosis factor α (TNF-α), IL-1β, and IL-6. On the other hand, some components (like glucose oxidase, gluconic acid, MGO, and polyphenols) show anti-inflammatory action via suppression of the production of certain molecules, like matrix metalloproteinases, and reactive oxygen species.^6^

The active principles in NS include thymoquinone (TQ), thymohydroquinone, dithymoquinone, thymol, carvacrol, nigellicine, nigellidine, and -hedrin. However, one of the integral components of *Nigella sativa* seeds is TQ with hydrophobicity and relatively smaller size that can easily pass-through infected cells’ plasma membranes. As it transits to the infected cells, TQ can bind to the lipophilic envelope of the SARS-CoV-2 virus due to its hydrophobic nature and, thus, destroy it before entering the cells. It exhibits its’ antibacterial potential especially by inhibiting biofilm synthesis in some bacteria. It has shown its’ antioxidant property via induction of the expression and/or activity of glutathione-S-transferase, glutathione peroxidase, superoxide dismutase and glutathione reductase.^7^ It modulates or inhibits inflammatory responses e.g., IL-1, IL-6, IL-10, IL-18, TNF-α, and nuclear factor-κB, hence, can significantly lower the chances of cytokine storm related COVID-19 mortality.^25^

The anti-diabetic, anti-hypertensive, cardio-protective and broncho-dilatory properties of HNS might make it even more beneficial in diabetic, hypertensive, cardiac and asthmatic patients which have a higher COVID-19 associated mortality.^12,26^ Furthermore, anti-platelet and anti-coagulant effects of HNS may also shield COVID-19 patients from thromboembolic complications, which are among the leading complications and causes of mortality.^27^ The hepato- and reno-protective nature of HNS may offer added precedence over other drugs in limiting COVID-19 related hepatic and renal injuries.^12,26^ Anti-pyretic, analgesic and antitussive properties of HNS can also provide symptomatic relief.^11^ Furthermore, HNS’s antimicrobial properties and synergism with other antibiotics against superimposed infections might prevent sepsis-related deaths.^6,8,14^ NICE and Public Heath England guidelines also recommend honey as the first line of treatment for an acute cough caused by upper respiratory tract infections, known as one of the defining symptoms of COVID-19. These findings strengthen the use of HNS as a potential candidate for combating SARS-CoV-2 worldwide.

In comparison to this, the recovery time reported for remdesivir was 10 days versus 15 days for the control (P<0.001),^3^ whereas lopinavir-ritonavir resulted in no decrease in the recovery time (16 days versus 16 days; P=0.09).^2^ In our study, in ∼50% of cases, SARS-CoV-2 RT-PCR became negative 4 days sooner in HNS groups than control groups. In previous studies, mortality among severe cases in comparison to control group was 27.0% (versus 25.0%) for hydroxychloroquine,^1^ 19.2% (versus 25.0%) for lopinavir-ritonavir,^2^ 15.7% (versus 24.0%) for convalescent plasma,^28^ 11.4% (versus 15.2%) for remdesivir,^3^ 22.9% (versus 25.7%) for dexamethasone,^4^ and only 4% (versus 18.87%) for HNS. Thus, HNS provided clinical superiority in reducing mortality in COVID-19 patients. Of note, combined mortality data provided by Solidarity and ACTT-1 for remdesivir and by Solidarity and Recovery trial for lopinavir-ritonavir failed to provide statistical improvement in mortality.^29^ In contrast to these drugs, HNS represents a safer and more affordable option that can be used as an in-house remedy.

A major limitation of this study was that the honey and NS were not administered as treatments to the patients separately. Patients on ventilator support were not enrolled in this study. A multinational study with a larger sample size is required to investigate potential variations in responses to the treatment in COVID-19 patients from different racial and ethnic origins.

## CONCLUSION

In conclusion, HNS has proven itself a safe and effective remedy for COVID-19 patients. It promotes viral clearance, quicker recovery and reduces mortality. Its affordability (< $5 for the whole treatment course), over the counter availability and ease of administration (as an easy home-based remedy) make this treatment even more feasible. Furthermore, as an inexpensive nutraceutical, HNS can be used alone or in combination with other drugs for additive effects. The treatment is very likely to reduce burden on health care systems in a significant manner.

## Research in context

### Evidence before this study

We searched PubMed from inception up to April 29, 2020, for clinical trials published in English probing proven efficacy of any drug against COVID-19 using the search terms (“COVID-19” [All Fields] OR “SARS-CoV-2” [All Fields]) (filters: Clinical Trial, Randomized Controlled Trial). We identified no drug proven effective against COVID-19 among randomized clinical trials. Meanwhile, we also searched PubMed for drugs that can be globally accessible with efficacy against viruses and other microbes along with anti-inflammatory, immuno-modulatory and anti-coagulant properties with wide safety profile. All original articles, review publications, meta-analysis and clinical trials published in English with these properties using the search terms (“antiviral” [All Fields] AND/OR “antibacterial” [All Fields] AND/OR (“anti-inflammatory” [All Fields] AND/OR (“immunomodulatory” [All Fields] AND/OR (“SARS-CoV-1” [All Fields]) (filters: none). Two products honey and *Nigella sativa* were found to be the most suitable for the search. No published clinical trial was identified on the effect of honey and *Nigella sativa* in patients with COVID-19.

### Added value of this study

To the best of our knowledge, this study is the first randomized, placebo-controlled clinical trial assessing the efficacy of oral honey and *Nigella sativa* seeds among adults with moderate or severe COVID-19. In the intention-to-treat analysis, we provided comprehensive methodical descriptions of clinical parameters, and clinical outcomes. In COVID-19 patients, honey and *Nigella sativa* with standard care therapy resulted in earlier viral clearance, symptomatic relief, clinical improvement and mortality reduction. Moreover, similarly to previous research no adverse effects were reported regarding HNS.

### Implications of all the available evidence

Considering the economic crisis related to the COVID-19 pandemic, the use of honey and *Nigella sativa* will particularly be beneficial for impoverished populations in resource limited settings. The inexpensive over the counter treatment regimen would be a valuable source to lower the burden on healthcare system while significantly dampening impact of the disease. Addition of these two nutraceuticals will add great value to lower the morbidity/mortality against COVID-19. The study will affect clinical practice and direct future research in the field of emerging infectious diseases. Nevertheless, these findings should be tested and replicated in further multi-national, larger clinical trials.

## Supporting information

Protocol

## Data Availability

Clinical data is available as additional file and raw data can be provided on request to corresponding author

## ACKNOWLEDGEMENTS

The authors would like to pay gratitude to all the trial steering committee members and patients who participated in this research. Special thanks to the Government of Pakistan and Smile Welfare Organization for providing free COVID-19 testing facilities and honey and *Nigella sativa* seeds. All the clinicians, paramedical and laboratory staff who assisted the conduction of this study are worth appreciating. The team would like to acknowledge Prof. Zaheer Ahmad, Ph.D., (Professor of Botany, Government University Lahore, Pakistan) for testing the purity of honey and *Nigella sativa* used in the trial.

## AUTHORS’ CONTRIBUTIONS

SA and ShA contributed equally to this paper and share joint first authorship. TM, QAS, AA, MA, and MI are senior authors. MA, MAI, LK, UNS, and IF were co-chief authors of this draft. SA, MAI, AA and MA contributed to conception, designing, acquisition of data, manuscript drafting and intellectual input. SA and MoA proposed the hypothesis and study design and obtained the funding. RA, MSS, KH, HR and ABA added the research delivery to the study centers. MA, MoA, SiA and MFN contributed biochemical, pharmacological and pharmaceutical inputs along with dosimetry. MKA, SoA, MAz and HZ led the development of data cleaning and analysis and took responsibility for the results in this draft and future analysis. SA, MoA, RA and AH drafted the first version of the manuscript. NM, IF, SR, AbH, ZA, AK, ZH, ShaA, HR, ABA, KH and AAr represented the conduction and validation of the data compilation and analysis in the manuscript. KN, MSu, SZ, IA, AH, AM, TM, SS, MeA, AA, MA, QAS and MI has overlooked the conduction and validity of the trial along with contributed to intellectual inputs in study protocol and methodology along with final manuscript write up. MeA and MI made sure validity of the data collection, data analysis and ethical considerations in their institutes. All authors are responsible for their contributions, providing critical edits and final authorization of the article. The corresponding author attest the authenticity of that all listed authors meet authorship criteria.

