## Supplementary material for "Honey and *Nigella sativa* against COVID-19 in Pakistan (HNS-COVID-PK): A multi-center placebo-controlled randomized clinical trial": Protocol

**Trial registration**

Registered at clinicaltrials.gov with registration ID **NCT04347382** and IRB Shaikh Zayed Medical Complex, Lahore approval ID **SZH/IRB/0026/2020** and IRB Services Institute of Medical Sciences approval ID **IRB/2020/658/SIMS**

This supplement contains the following items:

a. Original Protocol
b. Final Protocol
c. Summary of Changes
d. Original statistical analysis plan
e. Final statistical analysis plan
f. Summary of changes in statistical plan
g. Final Patient Consent Form
h. Questionnaire

**ABBREVIATIONS:**

| | |
|---|---|
| ARDS | Acute Respiratory Distress Syndrome |
| CBC | Complete Blood Count |
| CMV | Cytomegalovirus |
| COVID-19 | Corona Virus Disease 19 |
| CRP | C-Reactive Protein |
| DM | Diabetes Mellitus |
| ECG | Electrocardiogram |
| FAIR | Findable, Accessible, Interoperable, Re-usable |
| HIV | *Human Immunodeficiency Virus* |
| HSV | Herpes Simplex Virus |
| HTN | Hypertension |
| IRB | Institutional Review Board |
| LFTs | Liver Function Tests |
| MRSA | Methicillin-resistant Staphylococcus aureus |
| NICE | National Institute For Health And Care Excellence |
| RCT | Randomized Control Trial |
| RFTs | Renal Function Tests |
| RT-PCR | Real Time Polymerase Chain Reaction |
| SARS-CoV-2 | Severe acute respiratory syndrome coronavirus 2 |
| SIMS | Services Institute of Medical Sciences |
| SpO2 | Percentage of Oxygen Saturation |
| SSC-2 Score | Shaikh Zayed SARS CoV-2 Score |
| SZH | Shaikh Zayed Medical Complex |
| TQ | Thymoquinone |
| WHO | World Health Organization |
| $ZnCl_2$ | Zinc Chloride |

Original Clinical Study Protocol

Version date April 09, 2020

#### Title

Role of honey and *Nigella sativa* in management of COVID-19 (HNS-COVID-PK): A randomized controlled trial (Version 1.0)

#### Introduction

##### Background and rationale

An outburst of COVID-19 started in December 2019 due to SARS-CoV-2 in China, Wuhan which then rapidly spread to the entire world(1, 2). The epidemic of COVID-19 virus (later named as SARS-CoV-2), was declared as a pandemic on March 12, 2020 by WHO(3). A recent Chinese study illustrates that patients presenting with mild disease are 80% of the population and the overall mortality is around 2.3% but in patients aged 70 to 79 years this ratio is considerably high (8.0%) while in patients aged >80 years it's 14.8%(4). A total of 80,955 cases have been reported in China and other countries reported 41,953, as of March 11, 2020(5) and the number is in rising trend. Thus, the need of the hour is an effective treatment for symptomatic patients along with decreasing the interval of virus harboring and shedding to control its transmission in the society. A number of recent studies show promising anti-viral effects of hydroxychloroquine against SARS-CoV-2(6, 7) and others suggest the role of iron chelators and ZnCl2 as well in combating COVID-19(8, 9).

Herbal medicines are folk medicines used to treat diseases. The wide therapeutic effects of *Nigella sativa* seeds against many ailments such as dermatological problems, gastrointestinal diseases, ophthalmological issues, autoimmune diseases, respiratory tract infection, gynecological problems, diabetes and hypertension(10). Till now, various human clinical trials have been published and available to prove its efficacy and drug safety on PubMed. Its most active constituent, Thymoquinone (TQ), has delivered promising results against Gram-positive, Gram-negative bacteria, parasites, viruses, fungi and Schistosoma (10) TQ also possess anticonvulsant activity(11, 12), antioxidant(13), anti-inflammatory(14) and anti-cancer(15) properties. *Nigella sativa* has shown properties to inhibit viral replication and antiviral effects against CMV infection(16), inhibition of hepatitis C virus(16, 17) and a number of other viruses like HIV, H9N2, mosaic virus and new castle disease virus(10). *Nigella sativa's* ingredient, Thymoquinone has shown to decrease the viral load by inhibiting viral replication in coronavirus(18). Moreover, the presence of iron chelators and zinc in Nigella sativa(19, 20) makes it a perfect remedy as an anti-viral.

Almost around 6000 years ago, civilizations started using honey as a medicine. Previous records (1900–1250 BC) evidently reveal the effectiveness of honey as it was being used by different philosophers and scientists as a remedy for various diseases (21). But in 1960s with the emergence of highly efficient antibiotics, honey was categorized as a "worthless but harmless substance"(22). However, in this era of antibiotic resistance where we lack up to the mark and updated anti-viral, revived interest in the use of honey, both as an effective agent and as a therapeutic drug is essential to develop new methods of treatment. As results of honey's anti-bacterial, anti-viral, anti-oxidative and anti-inflammatory properties modern research has also proven the therapeutic importance of honey. It also exhibits anti-tussive, anti-cancerous (bladder cancer) and wound healing characteristics. A study has also shown the reproductive benefits of honey, including concentration of serum testosterone, sperm count and fertility(23). Moreover, it also acts as cardio protective (24) and exhibits anti-diabetic(25, 26) and laxative properties. A strong anti-bacterial activity against gram-positive bacteria (B.subtilis,S.aureus, and Paenibacillus larvae) (27) and gram negative bacteria (Pseudomonas aeruginosa, Enterobacter spp., Klebsiella) is also shown as mentioned in the studies(28). Furthermore, in case of Gram positive methicillin-resistant Staphylococcusaureus a 100% inhibition was observed(29) honey also shows antimicrobial effects via elevated Ph(21). Further studies show synergistic effect of honey with antibiotics agents like oxacillin, tetracycline, imipenem and meropenum and MRSA(30). It has its antiviral role against rubella virus, herpes simplex virus (HSV), hepatitis viruses and anti-Varicella Zoster virus(21). The recent therapies used as antiviral for covid-19 are remdesivir, lopinavir and ritonavir which are organic acids. Moreover, lopinavir also gives phenolic dimers(31) and both of these (organic and phenolic) properties are shown by honey(32).

Honey also shows immunity-booster effects via polyphenolic components. The proliferation of lymphocytes (B and T), along with the production of antibodies during primary and secondary immune response is stimulated via honey's phenolic ingredients. Honey activates the immune response via stimulation of human monocytic cells to release inflammatory cytokines (e.g. TNF-a, IL-1and IL-6)(33).

More than 250 human clinical trials have been conducted to prove its efficacy and drug safety and as per NICE and Public Health England guidelines recommend honey as the first line treatment remedy for acute cough caused by upper respiratory tract infection which is the cornerstone symptom in COVID-19.

Therefore, it is planned to conduct a clinical trial aiming at the combined effect of Nigella sativa and honey on SARS-CoV-2-infected patients. Aim of the study is to determine the use of Nigella sativa and honey in the treatment of Corona virus infection. This effect has not been seen in earlier trials.

**Objectives**

The objective of the study is to measure the efficacy of Honey and *Nigella sativa* in reducing viral load clearance and improving the clinical grading status along with reduction in time taken to resume normal activities as compared to control group and improvement in HRCT; preventing mortality and onset of pneumonia as compared to standard treatment for mild to moderate COVID-19 patients.

**Trial design**

This is a placebo-controlled, add-on, randomized trial using parallel group design. It is open-label, and sample size re-estimation adaptive, multi-centered, interventional design with 1:1 allocation ratio with a superiority framework.

**Methods: Participants, interventions and outcomes**

**Study setting**

Clinical sampling will be multi-centered, and RT-PCR (real time polymerase chain reaction) testing will be done in laboratories designated by the Pakistan government.

**Eligibility criteria**

All diagnosed and declared patients of COVID-19 admitted in corona centers of Pakistan during the study period.

Inclusion Criteria:

1. All suspected patients with a SSC-2 score (Figure 2) fulfilling the enrolment criteria (Figure 1)
2. Positive RT-PCR (Real Time Polymerase Chain Reaction)
3. HRCT (High Resolution Computerized Tomography) Chest score greater than 5 (using internationally standard nomenclature defined by the Fleischner Society glossary and peer-reviewed literature on viral pneumonia)
4. Both genders and age 18 years and above.
5. Patients who will present to seek medical care within 96 hours of ailment/randomization with positive RT-PCR and normal baseline laboratory parameters
6. Varying degree of COVID-19 severity

Exclusion Criteria:

1. Presence of co-morbidities like liver disease, immuno-compromised patient or any other chronic (other than Diabetes and Hypertension)
2. Females who are pregnant and breastfeeding
3. Uncontrolled Diabetes Mellitus and Hypertension
4. If a patient is allergic to either honey or *Nigella sativa*.
5. Refusal or withdrawal from consent.
6. patient on ventilator support or PaO2/FIO2 of less than 100
7. septic shock
8. Patients with positive SARS-CoV-2 screening during elective lists for any procedure will also excluded.

**Who will take informed consent?**

Site investigator will take informed written consent or assent from potential trial participants on specifically constructed informed consent forms.

**Additional consent provisions for collection and use of participant data and biological specimens**

Informed consent form will contain section on permission to draw and conduct specified tests on blood samples and conduct radiological investigations as per protocol of the study.

**Interventions**

**Explanation for the choice of comparators**

Both arms will be receiving standard care as per advised by treating consultant and “Clinical Management Guidelines for COVID-19 by Ministry of National Health Services, Pakistan”, standard care administered will comprise primarily on anti-pyretics, analgesics, antibiotics, multi-vitamins, intravenous fluids, supplemental oxygen, invasive/noninvasive ventilation, immune-modulatory agents, and inotropic support. Placebo comparator group will be placebo in addition to standard care while intervention arm will be receiving honey and *Nigella sativa*, as add-on.

**Intervention description**

High quality honey and *Nigella sativa* seeds will be provided by smile welfare organization, Lahore, Pakistan. Both products will be certified by botanical department of Government College University, Lahore Pakistan. *Nigella sativa* seeds (80 mg/kg/day) will be grinded and packed in capsules which will be given to the interventional group along with 1 gm/kg/day of honey stirred in 250ml of warm water in 2 to 3 divided doses, for a maximum of 13 days or until the patient recovers. This intervention will be given in addition to standard care as per advised by treating consultant and “Clinical Management Guidelines for COVID-19 by Ministry of National Health Services, Pakistan”, standard care administered will comprise primarily on anti-pyretics, analgesics, anti-virals, antibiotics, multi-vitamins, intravenous fluids, supplemental oxygen, invasive/noninvasive ventilation, immune-modulatory agents, and inotropic support.

**Strategies to improve adherence to interventions**

Assessment of participant’s adherence during clinical trial will be done by direct observation of dose administration by site investigators.

**Relevant concomitant care permitted or prohibited during the trial**

As per protocol of the hospital (study setting) the care and interventions permitted will be used and no specific prohibited care is in this trial

**Provisions for post-trial care**

No specific post trial care is needed in COVID-19 patients after recovery. No sound scientific evidence is there about harm with honey and *Nigella sativa* with the dose to be used in the trial.

**Recruitment criteria:**

All suspected patients will be assessed on SSC-2 score. If score is less than 5, the patient will be excluded from the study. If score is between 5 to 10, RT-PCR will be performed, if negative, patient will be excluded but if PCR is positive, patient will be included. For the patients with SSC-2 score greater than 10, PCR will be performed if positive, patient is enrolled but if negative HRCT will be done to rule out false negative patient. If HRCT positive for such patients, they will be included in study(36) if negative will be excluded.

**OUTCOMES**

**Primary Outcomes:**

1. Time taken for viral Load Clearance by $6^{th}$ Day
2. Clinical Grading Improvement at Day 6 on seven-point ordinal scale
3. Time Taken for Alleviation of Symptoms
4. HRCT Chest

**Secondary Outcomes:**

- Degree of Fever at day 4
- CRP at day 6
- Severity of symptoms at day 8
- 30 Day Mortality
- Clinical Grading Improvement at Day 6 on seven-point ordinal scale (as per World Health Organization R&D Blueprint Ordinal Clinical Scale).

**Additional Outcomes:**

- Median time to clinical improvement of COVID-19 symptoms

**RT-PCR** will be done on admission day (i.e. day 0) which would repeat after every 4th day for a maximum of 13 days or till the symptoms resolved and RT-PCR gets negative. RT-PCR is shown as positive or negative (because it is a limitation of our study of not getting the viral load). This study will include minimum of 15 patients (as per enrolment criteria) in each arm and these patients will be reviewed for 13 days(37) or patient is fully recovered or whichever comes first.

A maximum of 4 **HRCT chests** will be performed starting from day zero followed by every fourth day till patients PCR become negative(37).

HRCT SCORING will be done as follow:

Lungs divided into five lobes and each lobe is given 1 number. Total score is 25. Each lobe is scored from 0 to 5 as:

- 0 = no involvement
- 1 = <5% involvement
- 2 = 25% involvement
- 3 = 26%-49% involvement
- 4 = 50%-75% involvement
- 5 = >75% involvement

The individual lobar scores i.e. from 0 (no involvement) to 25 (maximum involvement) (39) make up the total HRCT chest score.

The HRCT findings are described via international standard terms, which are classified by the Fleischner Society glossary with peer-reviewed literature on viral pneumonia. The terms being used are ground glass opacity (GGO), crazy-paving pattern, and consolidation(38, 39).

**OPERATIONAL DEFINITION**

**Severity of symptoms** is classified as mild, moderate, severe and ARDS(40)

Mild: "symptoms of an upper respiratory tract viral infection i.e. low grade fever, dry cough, sore throat, nasal congestion, malaise but no radiological evidence of pneumonia"

Moderate: ". Moderate disease includes patients with Hypoxia (Oxygen saturation <94% but >90%) or chest X-ray with infiltrates involving <50% of the lung fields or fever, cough, sputum production, and other respiratory tract or non-specific symptoms along with manifestation of pneumonia on imaging but no signs of severe pneumonia"

Severe: "Fever is associated with severe dyspnoea, respiratory distress, tachypnea (> 30 breaths/min), and hypoxia ($SpO_2$ < 94% on room air)" or a $PaO_2$ to $FiO_2$ ratio of 300 or lower.

**Cough**

Mild: occasional, transient cough

Moderate: frequent cough, slightly influencing day time activities

Severe: frequent cough, significantly influencing daytime activities

**Fever**

Mild <100 °F

Moderate100-101.9 °F

Severe ≥ 102°F

**Myalgia**

It is classified as mild, moderate and severe on subjective basis. Patients have been provided with a scale of 1-10, with 1 being the lowest and 10 being the highest value.

Mild 1-3

Moderate 4-6

Severe >6-10

**Shortness of Breath**

It is classified as Grade1, Grade2, Grade3, Grade4 and Grade5.

Grade1=Not troubled by breathlessness except on strenuous exercise

Grade 2=Short of breath when hurrying on the level or walking up a slight hill

Grade 3=Walks slower than most people on the level, stops after a mile or so, or stop after 15 minutes walking at own pace

Grade 4=Stops for breath after walking about 100yds or a few minutes on level ground

Grade 5=Too breathless to leave the house, or breathless when undressing

**How Sick Do You Feel**

It is classified as mild, moderate and severe on subjective basis. Patients have been provided with a scale of 1-10, with 1 being the highest and 10 being the highest lowest

Mild >6-10

Moderate 4-6

Severe 1-3

**Duration of hospital stay:**

The number of days the patient stayed in the hospital setting during course of his/her treatment. The date of admission and date of discharge would give us the outcome in terms of good or poor prognosis.

**Mortality:** This criterion will tell us the number of patients died after 30 days of treatment with *Nigella sativa* and honey.

**Clinical Status Grading:**

It will be assessed every 4th day on a seven-pointer ordinal scale.

Grade1. Not hospitalized with resumption of normal daily life activities
Grade2. Not hospitalized but unable to resume normal daily life activities
Grade3. Hospitalized, but do not require supplemental oxygen therapy
Grade4. Hospitalized but require low flow supplemental oxygen therapy
Grade5. Hospitalized, requiring high flow supplemental oxygen

Grade6. Hospitalized, requiring mechanical ventilation

Grade7. Death

**Sample size**

15 patients in each arm with a total of 30 patients sample size in this pilot study to access the feasibility, efficacy, harmful effects, if any, followed by extension of study duration to three months.

**Recruitment**

Recruitment will be multi-centred and eligible patients will be identified using our eligibility criteria (mentioned above). Participants will be recruited to complete the desired sample size in each arm after taking complete written informed consent by the site investigators.

#### Assignment of interventions: allocation

##### Sequence generation

Stratification for age groups, gender, base line severity of symptoms and co-morbidities will be used to ensure that groups remain balanced in size for either arm after written informed consent to participate in our study. Randomization will be done using lottery method. As patients might be admitted at different times so they will be recruited after taking written informed consent (following all standard protocol for infection control and disinfection).

##### Concealment mechanism

Allocation sequence will be implemented immediately using sequential numbering once the patient is admitted in the corona center.

##### Implementation

Participants will be enrolled after written consent and allocation sequence will be generated using lottery method. Site investigator will assign participants to intervention.

**Assignment of interventions: Blinding**

**Who will be blinded?**

Care providers, outcome assessors and data analysts will be blinded, respectively, by using site investigators to provide placebo or drugs to participants, by using blinded clinicians to assess the clinical outcome and laboratory or radiological findings while by using statistical analysts from other institutions that will not be having any conflict of interest in research.

**Procedure for unblinding if needed**

Unblinding is permissible if the patients develop severe respiratory symptoms and need mechanical ventilation by revealing a participant's allocated intervention during the trial. If unblinding is required, the trial managers and data coordinators will have access to group allocations and any unblinding will be reported.

**Data collection and management**

Microsoft Excel will be used to ensure the data safety. Two site investigators will enter the data, recheck twice for possible errors separately, and make certain its integrity.  Principal investigators will visit twice weekly the study site while ethical committee will overview the study on weekly basis. Unexpected and unplanned visits will be made by trial steering committee members as well. There is no conflict of financial and non-financial interest with sponsors and researchers.

**Plans to promote participant retention and complete follow-up**

For hospitalized patients, all the data will be collected while the patient is already admitted in the hospital except 30-day mortality. For home quarantined patients a structured call will be made, and telemedicine will be used. Follow up will be done using phone numbers of the patients to access the mortality benefit of intervention.

**Data management**

Participants IDs will be used for confidentiality purposes and these IDs will be linked to demographic information securely and separately. The final data set of RCT will have coded data and can only be assessed by principal investigators. All outcomes will be doubled checked by the researchers prior to data collection and data storage. To ensure data's integrity and safety various meetings by the research team will be conducted

**Confidentiality**

In addition, confidentiality of participants' data will be ensured by using participants' IDs rather than identifiable information in the dataset (i.e., coding) and personal data will be optional for the patients considering their regional and religious believes though storing the document linking the IDs to the identifiable information separately and securely.

**Plans for collection, laboratory evaluation and storage of biological specimens for genetic or molecular analysis in this trial/future use**

Trained staff will collect nasopharyngeal swabs samples as per World Health Organization guidelines. Samples will be maintained at -80 degrees. Patients follow up will be done by investigators on 4th and 8th day. Those positive on 8th day will be followed up further for monitoring of primary end point (measuring time to return for COVID-19 PCR test to return negative).

As per biosafety and personal safety guidelines, nasopharyngeal swabs samples will be taken by site investigator from the participants at 4th and 8th day (those positive on 8th day will be followed up further) post treatment. The investigators will also monitor primary and secondary outcomes. Patients will be evaluated clinically on daily basis and relevant investigations will be repeated based on the symptoms or suspected complication.

#### Oversight and monitoring

##### Composition of the coordinating center and trial steering committee

Coordinating centre and trial executive committee will be comprised of medical lab technologists, biochemists, clinical pharmacists, clinical pharmacologists and toxicologists, virologists, immunologists, biostatisticians, public health experts, epidemiologists, ethical experts and consultants of medicine, pulmonology, cardiology and infectious disease departments. This committee will be responsible for the safety and results of endpoints. This will have all the authority to stop the clinical trial all together. Site investigators will be responsible for data collection and quality check of data at collection points/ study setting.

##### Composition of the data monitoring committee, its role and reporting structure

Data management team will be comprised of national and international level investigators. This team will be responsible to ensure the execution of the clinical trial in best possible way as defined by the study protocol. Data monitoring committee has a biostatistician, an epidemiologist, and public health experts, study chair and study director in it. There is no financial or non-financial conflict of interest. For the initial pilot phase, no funding/sponsorship are obtained.

##### Executive committee

The executive committee vouches for the completeness and accuracy of the data and for the fidelity of the trial to the protocol.

##### Adverse event reporting and harms

The researchers will record any adverse, unpredictable or undesirable sign and symptom and it will be discussed with the care providers. A comprehensive evaluation will be conducted to evaluate the co-relation between experimental drug and the developing sign and symptoms. The investigator will respond appropriately to ensure the wellbeing of the patient in case of above-mentioned unforeseen event and all the details will be written carefully. Moreover, regular follow-up will be made certain until the patient regains his/her health. Furthermore, during this study the participants will be insured for the financial compensation, if they are harmed via interventional drug.

##### Frequency and plans for auditing trial conduct

Auditing of the clinical trial will be done by trail steering committee in zoom meeting where the entire audit will be provided by the site investigators and research coordinators.

##### Plans for communicating important protocol amendments to relevant parties (e.g. trial participants, ethical committees)

In order to modify protocols including eligibility criteria and outcomes, permission will be needed to get it approved by the trial steering committee and the notification will be done to relevant parties including IRB, trial registries and journals.

##### Dissemination plans

The Publications subcommittee will review all publications following their guidelines and report its recommendations to the Steering Committee.

##### Trial status

Protocol version date and number: April 09, 2020 Original version.

Patient recruitment will begin in April 2020 and it will end by July 2020.

##### Availability of data and materials

The datasets used or analysed during the current study will be available from the corresponding author upon reasonable request.

##### Competing interests

No conflict of financial or other competing interests for the overall trial.

**Funding:**

Funding will be provided by smile welfare organization, Lahore, Pakistan and Shaikh Zayed Medical Complex, Lahore, Pakistan and Services Institute of Medical Sciences

### Enrolment Criteria:

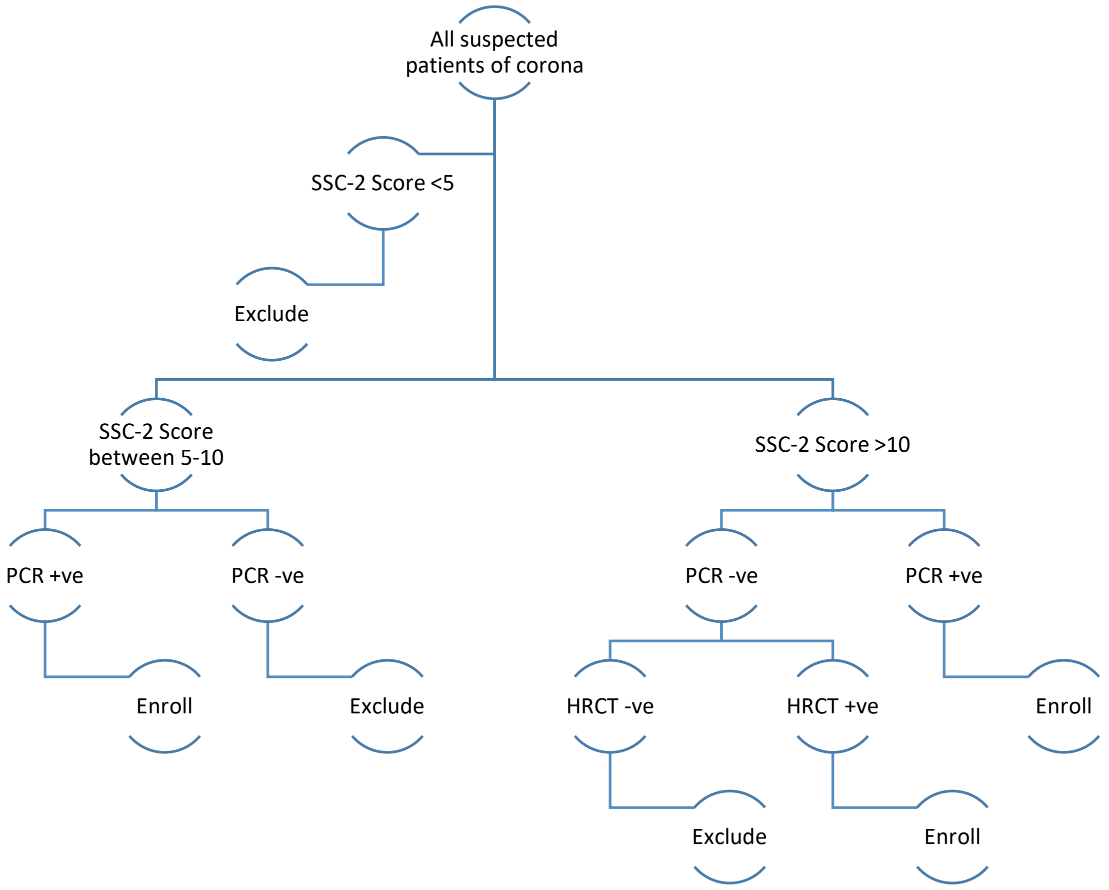


**Figure 1:** All suspected patients will be assessed on SSC-2 score. If score is less than 5, the patient will be excluded from the study. If score is between 5 to 10, RT-PCR will be performed (41) , if negative, patient will be excluded but if PCR is positive, patient will be included. For the patients with SSC-2 score greater than 10, PCR will be performed if positive, patient is enrolled but if negative HRCT will be done to rule out false negative patient. If HRCT positive for such patients, they will be included in study if negative will be excluded.

### CONSORT 2010 Flow Diagram

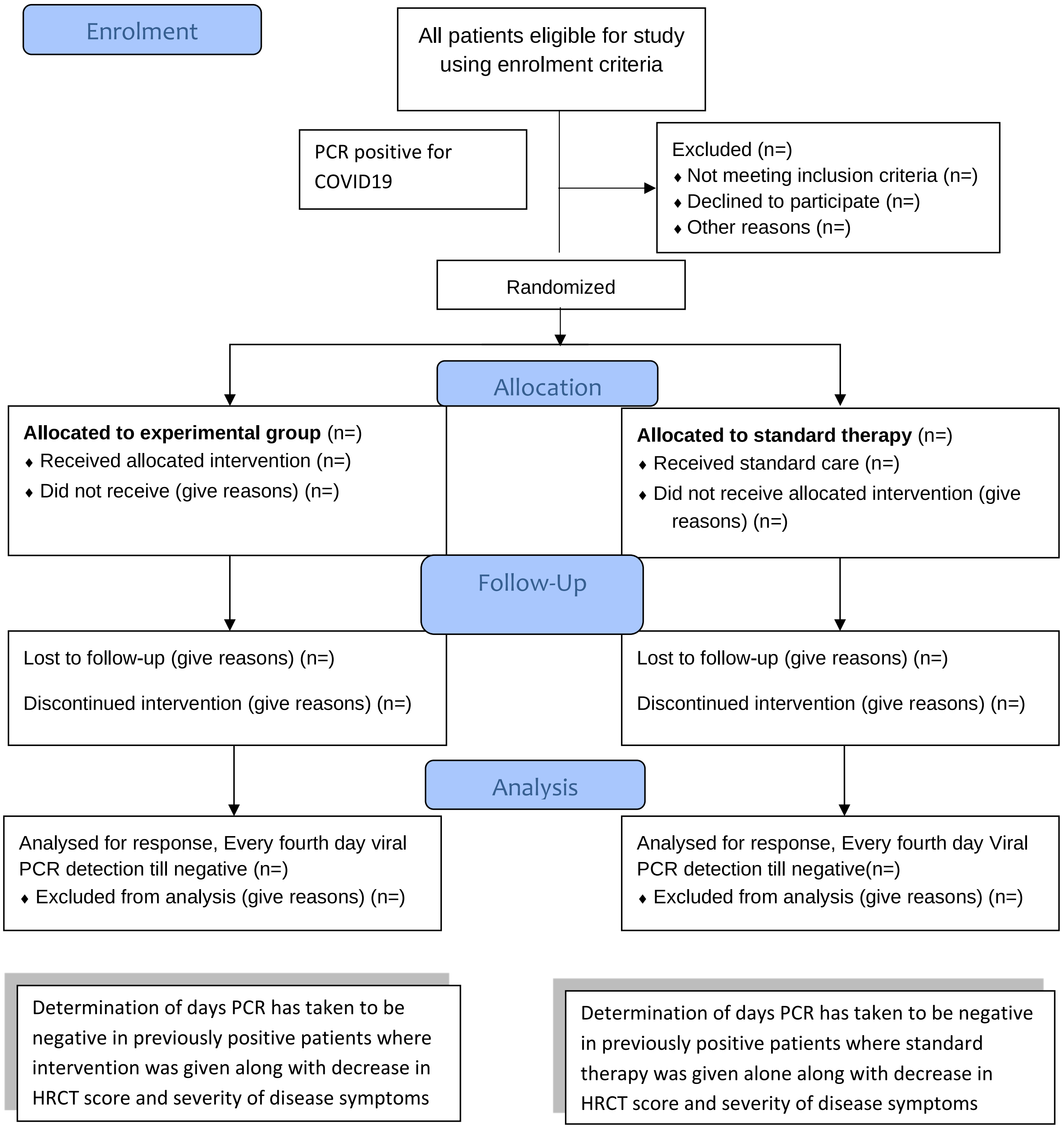

**SSC-2 (Shaikh Zayed SARS-CoV-2) SCORE**

| | |
|---|---|
| High grade Fever (without any other evident source of infection) | 5 |
| Low grade Fever (without any other evident source of infection) | 3 |
| Fever (with evident source of infection) | 2 |
| Fatigue/Myalgia | 2 |
| Dry Cough | 2 |
| Dyspnea | 3 |
| Any Atypical Symptom* | 1 |
| ARDS ` | 5 |
| Any other diagnosis unlikely | 2 |

**Score of less than 5:** Low probability

**Score of between 5 &10:** Moderate probability (need PCR to label as Corona infected Patient)

**Score of more than 10:** High probability

*(Headache, Anorexia, Sputum production, Pharyngitis, GI disturbances)

Figure 2: A score based on prevalence of symptoms reported to diagnose/ suspect patients with corona as per reported in multiple studies in COVID-19 patients

**(Consent Form for Medical Treatment)**

MR NO: .................................

Date: .................................

Patient Name: .................................

Age: .................................

Father/husband Name: .................................

I, hereby give full consent to my doctor for History taking, medical examination, and prescription.

I, hereby declare that I will not do any legal proceedings in future in any

Circumstances.

Patient signature/thumb impression:

CNIC Number:

Relative/family member signature:

CNIC Number:

Final Clinical Study Protocol:

Version date April 30, 2020

**Title**

Role of honey and *Nigella sativa* in the management of COVID-19 (HNS-COVID-PK): A randomized controlled trial

**Background and rationale**

Sudden appearances of cluster pneumonia cases started in December 2019in Wuhan, China which was diagnosed to be the cause of SARS-CoV-2. Over time, the disease named as COVID-19 rapidly travelled to the entire world(1, 2). The COVID-19 epidemic was declared as a global pandemic on March 12, 2020 by WHO(3). Researchers throughout the world took the notice of proceedings and started exploring different facets of the disease. In context of its severity, a recent study illustrates that 80% of the positive tested patients present with mild disease symptoms and the overall mortality lies around 2.3%. However, in patients aged 70 to 79 years, this ratio is considerably higher (8.0%) while it reaches to 14.8% in patients aged >80 years(4). It is estimated that majority of cases are disregarded due to either asymptomatic nature, lack of testing facilities or social dilemmas. What requires our immediate attention is the pursuit of an effective treatment for symptomatic patients. On the same lines, an intervention should also be sought to decrease viral latency duration of its shedding in order to overcome transmission in the community. In response to this emergency, a number of recent studies have been conducted to explore a potential miracle drug. Such efforts included novel antivirals like remdesivir and favipiravir on one side while testing many existing antiviral drugs like lopinavir/ritonavir, darunavir/umifenovir and ivermectin on the other side(6-12). Among the main highlights of such findings is hydroxychloroquine, which has shown promising anti-viral effects against SARS-CoV-2(13-15). With lesser media reporting, other studies have also suggested a positive impact of iron chelators and $ZnCl_2$ as well, in combating COVID-19. In parallel to these lines, many traditional therapies are also being interrogated for their possible antiviral properties against COVID-19(16, 17).

Herbal extracts, evidence based medicine, traditional therapeutics and therapies from folklores have continuously been put to rigorous scientific investigations in the past. One of the most studied amid traditional herbs is *Nigella sativa*. Lately, the widespread therapeutic effects of *Nigella sativa* seeds against many ailments have been historically documented and have been tested by researchers. The broad spectrum of ailments for *Nigella sativa* treatment include dermatological problems, gastrointestinal diseases, ophthalmological issues, autoimmune diseases, respiratory tract infections, gynecological problems, diabetes and hypertension(18). Till now, PubMed indexing reports more than 50 human clinical trials that provide a good rationale about its efficacy and safety. After fractionation, the most active constituent, Thymoquinone (TQ) has been attributed mainly for its activities. TQ has also been demonstrated to possess anticonvulsant(19, 20), antioxidant(21), anti-inflammatory(22) and anti-cancer(23)properties. What has been interesting for us is its significant antimicrobial value which has shown promising results against Gram-positive and Gram-negative bacteria, parasites, viruses, fungi and Schistosoma(18)

In milieu of our trial, it is quite encouraging that *Nigella Sativa* has been shown previously to inhibit viral replication, demonstrated antiviral effects against CMV (24), hepatitis C virus(24, 25) and a number of other viruses such as HIV(26), H9N2(18, 27), mosaic virus and new castle disease virus(18, 28). We have quite encouraging findings which display that *Nigella sativa's* main ingredient, TQ has been shown to decrease viral replication in coronavirus-infected cells (SARS-CoV-1)(29). Moreover, the reports of naturally occurring iron chelators and zinc in Nigella Sativa(30, 31) which are already under exploration in COVID-19, additionally may contribute to its antiviral effects against SARS-CoV-2.

The second ingredient of our Anti-Covid-19 mix is natural honey. From the last six eras, human civilization records demonstrate the use of honey as medicine. Previous scrolls (1900–1250 BC) evidently reveal the effectiveness of honey as it was being used by different philosophers and physicians as a remedy for various illnesses(32). Clinical data indicates that honey possesses strong anti-tussive, anti-cancerous (bladder cancer) and wound healing characteristics (Manuka Honey). Although traditionally believed from a long time, a study has confirmed the reproductive system benefits of honey including increases in concentration of serum testosterone, enhancements of sperm count and increased chances of conception(32). Moreover, it also acts as cardio-protective(33) , exhibits anti-diabetic(34, 35) and laxative properties. Our interest in honey revolves around its strong antimicrobial efficacy. Despite being a food in nature, honey is immune to microbial attacks. In the 1960's with the emergence of highly efficient antibiotics, honey was comparatively perceived as a "worthless but harmless substance"(36). However, a recent burst in emergence of antibiotic resistance where we lack remedies against highly resistant microbes, attentiveness towards honey is being resuscitated. Many studies have optimized the use of honey as an effective antimicrobial agent and as a therapeutic drug for various infections. As honey has been reported by scientific annals to possess anti-bacterial, anti-viral, anti-oxidative and anti-inflammatory properties, modern research has verified the therapeutic inevitability of honey. Honey is bactericidal. A strong anti-bacterial activity against Gram-positive bacteria (B.subtilis,S.aureus, and Paenibacillus larvae)(37) and Gram-negative bacteria (Pseudomonas aeruginosa, Enterobacter spp., Klebsiella) has been demonstrated in different studies(38). Furthermore, in cases of highly resistant Gram-positive methicillin-resistant *Staphylococcus aureus*, a 100% inhibition was detected(39). Such antimicrobial effects of Honey could be attributed towards its sugar concentration, its unique constituents and elevated pH values(32). Further studies show synergistic effects of honey with antibiotic agents comparable to oxacillin, tetracycline, imipenem and meropenem against MRSA(40). In perspective of its antiviral activity, as hypothesized in our trials, previously it has demonstrated antiviral effects against rubella virus, herpes simplex virus (HSV), hepatitis and anti-Varicella Zoster virus(32). A stimulating aspect of currently tested anti-covid-19 therapies could be the common trait of being acidic in nature. The recent therapies tested as antiviral drugs for COVID-19 are remdesivir, lopinavir and ritonavir which are organic acids, like honey. Another aspect of a prominent similarity is that lopinavir gives phenolic dimers(41) and both of these (organic and phenolic) properties are also shown by honey(42).

In addition to its direct antiviral effects, honey also exhibits immunity-booster effects mainly via polyphenolic components. The proliferation of lymphocytes (B and T), along with the production of antibodies during primary and secondary immune response is stimulated via honey's phenolic ingredients(43). Honey is known to activate the immune response via stimulation of human monocytic cells to release inflammatory cytokines (e.g. TNF-a, IL-1and IL-6)(43). Taken together, honey could be a vastly effective therapeutic agent against COVID-19 either through its antiviral properties, its immune boosting characteristics or the blend of both.

While considering its safety profile in COVID-19 patients, we have found that more than 250 human clinical trials have been conducted to determine efficacy and safety profiles of honey in different ailments. As an outcome, NICE and Public Heath England guidelines recommend honey as the first line of treatment for an acute cough caused by upper respiratory tract infections, known as one of the defining symptoms of COVID-19.

We believe that we have surplus data regarding proposed activities, pharmacokinetics and safety profiles in favor of our proposed therapeutic regimen. Therefore, we have planned to conduct a clinical trial aiming at the observation of combined effect of *Nigella sativa* and honey on SARS-CoV-2-infected patients. The aim of the study is to explore the possible therapeutic use of *Nigella sativa* and honey in the treatment of Corona virus infection. This effect has not been investigated in earlier human trials.

**Objectives**

The objective of the study is to measure the efficacy of Honey and *Nigella sativa* in reducing viral load clearance and improving the clinical grading status along with reduction in time taken to resume normal activities as compared to control group.

**Trial design**

This is a placebo-controlled, add-on, randomized trial using parallel group design. It is an open-labeled and sample size re-estimation adaptive, multi-center and interventional study with a 1:1 allocation ratio design with superiority framework.

**Methods: Participants, interventions and outcomes**

**Study setting**

The study centers chosen for this clinical trial will include:

- Services Institute of Medical Sciences, Lahore, Pakistan
- Shaikh Zayed Medical Complex, Lahore, Pakistan
- Doctors lounge for Telemedicine (Rustam Park Lahore, Pakistan)
- Ali clinic for OPD cases (Sandha, Lahore, Pakistan)

RT-PCR testing will be done in centers established by Government of Pakistan.

**Eligibility criteria**

All COVID-19 patients presenting in study center for medical help will be enrolled during the study period.

<u>Inclusion Criteria:</u>

- All suspected patients with an SSC-2 score (Figure 2) fulfilling the enrolment criteria (Figure 1)
- Positive RT-PCR (Real Time Polymerase Chain Reaction)
- Patients who will present to seek medical care within 96 hours of ailment/randomization with positive RT-PCR and normal baseline laboratory parameters
- Varying degree of COVID-19 severity
- Both genders and age 18 years and above

<u>Exclusion Criteria:</u>

- Presence of co-morbidities like liver disease, immuno-compromised patient or any other chronic ailment (other than Diabetes and Hypertension)
- Females who are pregnant and breast feeding
- Uncontrolled Diabetes Mellitus and Hypertension
- If a patient is allergic to either honey or *Nigella sativa*.
- Refusal or withdrawal from informed consent.
- patient on ventilator support or $PaO_2/FIO_2$ of less than 100
- Hemodynamic instability
- Patients with positive SARS-CoV-2 screening during elective lists for any procedure will also be excluded.

**Who will take informed consent?**

Site investigator will take written informed consent from all trial participants by giving them the specifically constructed informed consent form (provided at the end of proposal)

**Additional consent provisions for collection and use of participant data and biological specimens**

Site investigators will also be responsible for taking consent regarding every other matter if required. Informed consent form will contain a section on permission to draw and conduct specified tests on blood samples and conduct radiological investigations as per protocol of the study.

##### Interventions

##### Explanation for the choice of comparators

Both arms will be receiving standard care as per advised by treating consultant and "Clinical Management Guidelines for COVID-19 by Ministry of National Health Services, Pakistan", standard care administered will comprise primarily of anti-pyretic, analgesics, anti-virals, antibiotics, multi-vitamins, intravenous fluids, supplemental oxygen, invasive/noninvasive ventilation, immune-modulatory agents, and inotropic support. The placebo comparator group will receive a placebo (empty capsule) in addition to standard care while the intervention arm will be receiving honey and *Nigella sativa* an add-on.

##### Intervention Description

High quality honey and *Nigella sativa* seeds will be provided by smile welfare organization, Lahore, Pakistan. Both products will be certified by botanical department of Government College University, Lahore Pakistan. *Nigella sativa* seeds (80 mg/kg/day) will be grinded and packed in capsules which will be given to the interventional group along with 1 gm/kg/day of honey stirred in 250ml of warm water in 2 to 3 divided doses, for a maximum of 13 days or until the patient recovers. This intervention will be given in addition to standard care as per advised by treating consultant and Clinical Management Guidelines for COVID-19 by Ministry of National Health Services, Pakistan, standard care administered will comprise primarily of anti-pyretic, analgesics, antibiotics, multi-vitamins, intravenous fluids, supplemental oxygen, invasive/noninvasive ventilation, immune-modulatory agents, and inotropic support.

##### Criteria for discontinuing or modifying allocated interventions

Fixed dose will be given throughout the study and interventional drug administration will be stopped immediately in following conditions:

1. Any adverse drug reaction
2. Organ failure secondary to honey and *Nigella sativa*
3. Patient denies/back off from the further participation
4. Patients on ventilator support

Regardless of any of the condition, participant's data will be retained and analyzed in the trial in order to follow-up and prevent any data missing.

##### Strategies to improve adherence to interventions

To improve adherence to the intervention, participants will be counselled about the advantages of this study. All the participants will be monitored regarding compliance to their assigned treatment strategy and health professionals will give drugs. As this trial will include admitted patients and patients enrolled via telemedicine and OPD direct observational method will be followed for admitted patients and structured telephone calls for 13 days should be sufficient to make sure compliance and adherence to the interventions for telemedicine and OPD enrolled patients (in case, whenever a OPD patient misses a visit in his/her follow up 13$^{th}$ day period). In case of unsuccessful phone calls, site investigators will be asked for additional information including phone numbers of close relatives, e-mail addresses, and attempted again to contact the patient or a relative. If these attempts would go unsuccessful, as a last resort, for patients living in Lahore, a trained person will visit the patient's house and obtain follow-up data. Distribution of honey, *Nigella sativa* and pulse oximeters to the patients of telemedicine and OPD will be the responsibility of Smile Welfare Organization.

##### Relevant concomitant care permitted or prohibited during the trial

As per hospital protocol (study setting), the care and interventions permitted will be used and no specific prohibited care is in this trial.

##### Provisions for post-trial care

No specific post trial care will be needed in COVID-19 patients after recovery. No sound documented scientific evidence is there about adverse effects with Honey or *Nigella sativa* with this dose for this trial.

**Recruitment criteria**:

All suspected patients will be assessed on SSC-2 score. If a score is less than 5, the patient will be excluded from the study. If the score is ≥5, RT-PCR will be performed, if first RT-PCR negative, patient will be excluded but if PCR is positive, patient will be included. (This decision is made on first RT-PCR)

**OUTCOMES**

**Primary Outcomes:**

- Time taken for viral load clearance
- Clinical status grading at Day 6 (defined by seven-point ordinal scale as per World Health Organization R&D Blueprint Ordinal Clinical Scale).
- Time taken for alleviation of symptoms

**Secondary Outcomes:**

- Degree of Fever at day 4
- CRP at day 6
- Severity of symptoms at day 8
- Clinical Status grading at day 10
- 30 Day Mortality

**Additional Outcomes:**

- Median time to clinical improvement of COVID-19 symptoms

**RT-PCR** will be done on admission day (i.e. day 0) which will be repeated once patient is asymptomatic for more than 48 to 72 hours. If the PCR is still positive, it will be repeated on day $14^{th}$. RT-PCR is shown as positive or negative (because it is a limitation of our study of not getting the viral load).

**OPERATIONAL DEFINITION**

**Severity of respiratory symptoms**:

- Mild: “symptoms of an upper respiratory tract viral infection i.e. low-grade fever, dry cough, sore throat, nasal congestion, malaise but no radiological evidence of pneumonia”
- Moderate: “. Moderate disease includes patients with Hypoxia (Oxygen saturation <94% but >90%) or chest X-ray with infiltrates involving <50% of the lung fields or fever, cough, sputum production, and other respiratory tract or non-specific symptoms along with manifestation of pneumonia on imaging but no signs of severe pneumonia”
- Severe: “Fever is associated with severe dyspnoea, respiratory distress, tachypnea (> 30 breaths/min), and hypoxia ($SpO_2$ < 94% on room air)” or a $PaO_2$ to $FiO_2$ ratio of 300 or lower.

**Grading of Clinical Status:**

Grading of clinical status will be assessed at 0, 4, 6, 8, 10, 12th day considering patient condition on that moment.

- Grade 1. Not hospitalized with resumption of normal daily life activities
- Grade 2. Not hospitalized but unable to resume normal daily life activities
- Grade 3. Hospitalized, but do not require supplemental oxygen therapy
- Grade 4. Hospitalized but require low flow supplemental oxygen therapy
- Grade 5. Hospitalized, requiring high flow supplemental oxygen
- Grade 6. Hospitalized, requiring mechanical ventilation
- Grade 7. Death

**Cough**

- Mild: occasional, transient cough
- Moderate: frequent cough, slightly influencing day time activities
- Severe: frequent cough, significantly influencing daytime activities

**Fever**

- Mild <100 °F
- Moderate100-101.9 °F
- Severe ≥ 102°F

**Myalgia**

It is classified as mild, moderate and severe on subjective basis. Patients have been provided with a scale of 1-10, with 1 being the lowest and 10 being the highest value.

- Mild 1-3
- Moderate 4-6
- Severe >6-10

**Shortness of Breath**

- It is classified as Grade1, Grade2, Grade3, Grade4 and Grade5.
- Grade1=Not troubled by breathlessness except on strenuous exercise
- Grade 2=Short of breath when hurrying on the level or walking up a slight hill
- Grade 3=Walks slower than most people on the level, stops after a mile or so, or stop after 15 minutes walking at own pace
- Grade 4=Stops for breath after walking about 100yds or a few minutes on level ground
- Grade 5=Too breathless to leave the house, or breathless when undressing

**How Sick Do You Feel**

It is classified as mild, moderate and severe on subjective basis. Patients have been provided with a scale of 1-10, with 1 being the highest and 10 being the lowest

- Mild >6-10
- Moderate 4-6
- Severe 1-3

**Time Taken for alleviation of symptoms:**

Number of days taken by the patients in experimental and control groups for resolution of symptoms.

**Sample size and Study Duration**

As a pilot project, 15 patients in each arm will be taken initially for this multi-centered study in Pakistan. Later on, this study will be expanded considering the feasibility of the trial, for the 3 months from the start of study.

**Recruitment**

Recruitment will be done from Services Institute of Medical Sciences, Shaikh Zayed Medical Complex, Ali Clinic and Doctors Lounge from COVID-19 wards, OPD and telemedicine, respectively. Eligible patients will be identified using our eligibility criteria (mentioned above). Participants will be recruited in each arm after taking complete written informed consent by the site investigators.

##### Assignment of interventions: allocation

###### Sequence generation

Stratification will be done via SAS software version 9.4 for age groups, gender, additional drugs administered, base line severity of symptoms and co-morbidities will be used to ensure that groups remain balanced in size for either arm after written informed consent to participate in our study. Randomization will be done using a lottery method. As patients might be admitted at different times, they will be recruited after taking written informed consent (following all standard protocol for infection control and disinfection). To minimize allocation bias, we will perform allocation concealment with an interactive voice/web-based response system until randomization will be finished on the system through a computer or phone.

###### Concealment mechanism

The allocation sequence will be computer generated that will be concealed from all site investigators, allocated participants and treatment providers until the final interventional allocation is done.

**Assignment of interventions: Blinding**

**Who will be blinded?**

Care providers, outcome assessors and data analysts are blinded, respectively, by using site investigators to provide placebo or drugs to participants, by using blinded clinicians to assess the clinical outcome and laboratory or radiological findings while by using statistical analysts from other institutions that are not having any conflict of interest in research.

**Procedure for unblinding if needed**

Unblinding is permissible if the patients develop severe respiratory symptoms and need mechanical ventilation by revealing a participant's allocated intervention during the trial. If unblinding is required, the trial managers and data coordinators will have access to group allocations and any unblinding will be reported.

**Data collection and management**

Microsoft Excel will be used to ensure the data safety. Two site investigators will enter the data, recheck twice for possible errors separately, and make certain its integrity. Principal investigators will visit twice weekly the study site while ethical committee will overview the study on weekly basis. Unexpected and unplanned visits will be made by trial steering committee members as well. There is no conflict of financial and non-financial interest with sponsors and researchers.

**Plans to promote participant retention and complete follow-up**

All the data will be collected while the patient is already admitted in the hospital except 30-day mortality. For home quarantined patients a structured call will be made, and telemedicine will be used. Follow up will be done using phone numbers of the patients to access the mortality benefit of intervention.

**Data management**

Participants IDs will be used for confidentiality purposes and these IDs will be linked to demographic information securely and separately. The final data set of RCT will have coded data and can only be assessed by principal investigators. All outcomes will be doubled checked by the researchers prior to data collection and data storage. To ensure data's integrity and safety various meetings by the research team will be conducted

**Confidentiality**

In addition, confidentiality of participants' data will be ensured by using participants' IDs rather than identifiable information in the dataset (i.e. coding) and personal data will be optional for the patients considering their regional and religious believes though storing the document linking the IDs to the identifiable information separately and securely.

**Plans for collection, laboratory evaluation and storage of biological specimens for genetic or molecular analysis in this trial/future use**

Baseline laboratory investigations and nasopharyngeal swab samples will be obtained among suspected patients of COVID-19 (before randomization and allocation will be administered). Those who would be enrolled, second SARS-CoV-2 PCR testing will be performed between 48-96 hours once the patient would be asymptomatic or touched baseline health. In case, if second PCR would still positive third PCR will be executed at 14$^{th}$ day with no further follow up. Trained staff will collect nasopharyngeal swabs samples as per biosafety and personal safety guidelines of World Health Organization. Samples will be maintained at -80 degrees. Patients' follow up will be done by 4 different teams of site investigators (Team A, B, C and D). Each team will consist of two site investigators. Team A will be responsible for recruitment of patients. Team B will monitor symptoms daily. Team C will access clinical status grading on day 0, 4, 6 ,8,10 and 12 and team D will be responsible for PCR sampling.

#### Oversight and monitoring

##### Composition of the coordinating center and trial steering committee

Coordinating centre and trial executive committee will be comprised of medical lab technologists, biochemists, clinical pharmacists, clinical pharmacologists and toxicologists, virologists, immunologists, biostatisticians, public health experts, epidemiologists, ethical experts and consultants of medicine, pulmonology, cardiology and infectious disease departments. This committee will be responsible for the safety and results of endpoints. This will have all the authority to stop the clinical trial all together. Site investigators will be responsible for data collection and quality check of data at collection points/ study setting.

##### Executive committee

The executive committee vouches for the completeness and accuracy of the data and for the fidelity of the trial to the protocol.

##### Data Collection:

Data will be collected daily, from randomization until day 13, in the case-report form. For the patients who would be discharged before day 13, a structured telephone call to the patient or patient's family will be conducted daily by an interviewer who would be unaware of the assigned trial group in order to assess vital status and return to routine activities. A final follow-up call will be made on day 30. For the patients who will be enrolled via telemedicine same protocol of daily structured telephone calls for 13 days and a 30 day follow up call will be applied.

##### Limitation:

Patients with multi-organ failure and requiring invasive positive ventilation will be excluded from the study.

##### Adverse event reporting and harms

The researchers will record any adverse, unpredictable or undesirable sign and symptom and it will be discussed with the care providers. A comprehensive evaluation will be conducted to evaluate the co relation between experimental drug and the developing signs and symptoms. The investigator will respond appropriately to ensure the wellbeing of the patient in case of any unforeseen event and all the details will be written carefully. Moreover, regular follow-ups will be made certain until the patient regains his/her health. If the adverse event happens during the study intervention will be reported to Institutional Review Board (IRB).

##### Frequency and plans for auditing trial conduct

Auditing of the clinical trial will be done by trial steering committee in meetings where all the audits will be provided by the site investigators and research coordinators.

##### Plans for communicating important protocol amendments to relevant parties (e.g. trial participants, ethical committees)

In order to modify protocols including eligibility criteria and outcomes, permission will be needed to get approved by the trial steering committee and the notification will be done to relevant parties including IRB trial registries and journals.

##### Dissemination plans

The publications subcommittee will review the publication, all the endpoint data, primary outcome analysis and the study results and recommend the changes to the author. After the changes being done it will finally submit its recommendations to the steering committee for approval. Study results will be disclosed to all study participants, member physicians, patients and other medical personnel.

##### Trial status

Protocol version date and number: April 30, 2020; finalized version. Patient recruitment will be completed by July 2020.

**Availability of data and materials**

The datasets used or analysed during the current study will be available from the corresponding author upon reasonable requests.

**Competing interests**

The authors affirm that they have no competing interests and according to the standards of scientific integrity the publication of both positive and negative study results will be ensured. All the study data (if reasonable) will be made accessible guided by the FAIR principles along with the perspective of relevant laws and privacy regulations. Authorship eligibility follows conventional academic standards. No professional writers have been involved in this.

**Funding:**

Funding has been provided by smile welfare organization and Shaikh Zayed Medical Complex, Lahore, Pakistan and Services Institute of Medical Sciences for drugs and testing.

### Enrolment Criteria:

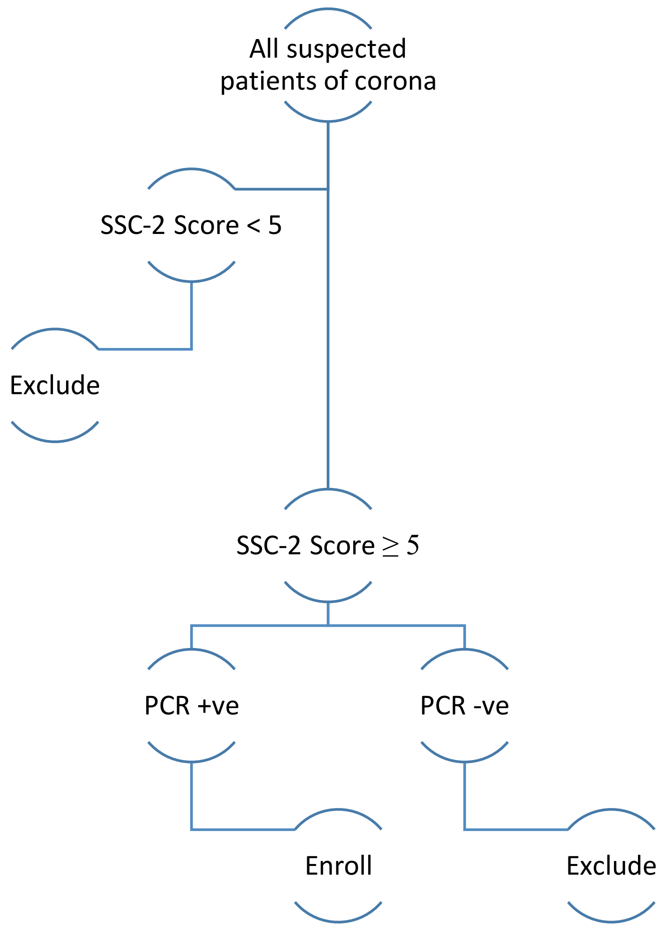


**Figure 1:** All suspected patients will be assessed on SSC-2 score. If a score is less than 5, the patient will be excluded from the study. If score is greater than 5, RT-PCR will be performed (41) , if negative, patient will be excluded but if PCR is positive, patient will be included.

#### CONSORT 2010 Flow Diagram

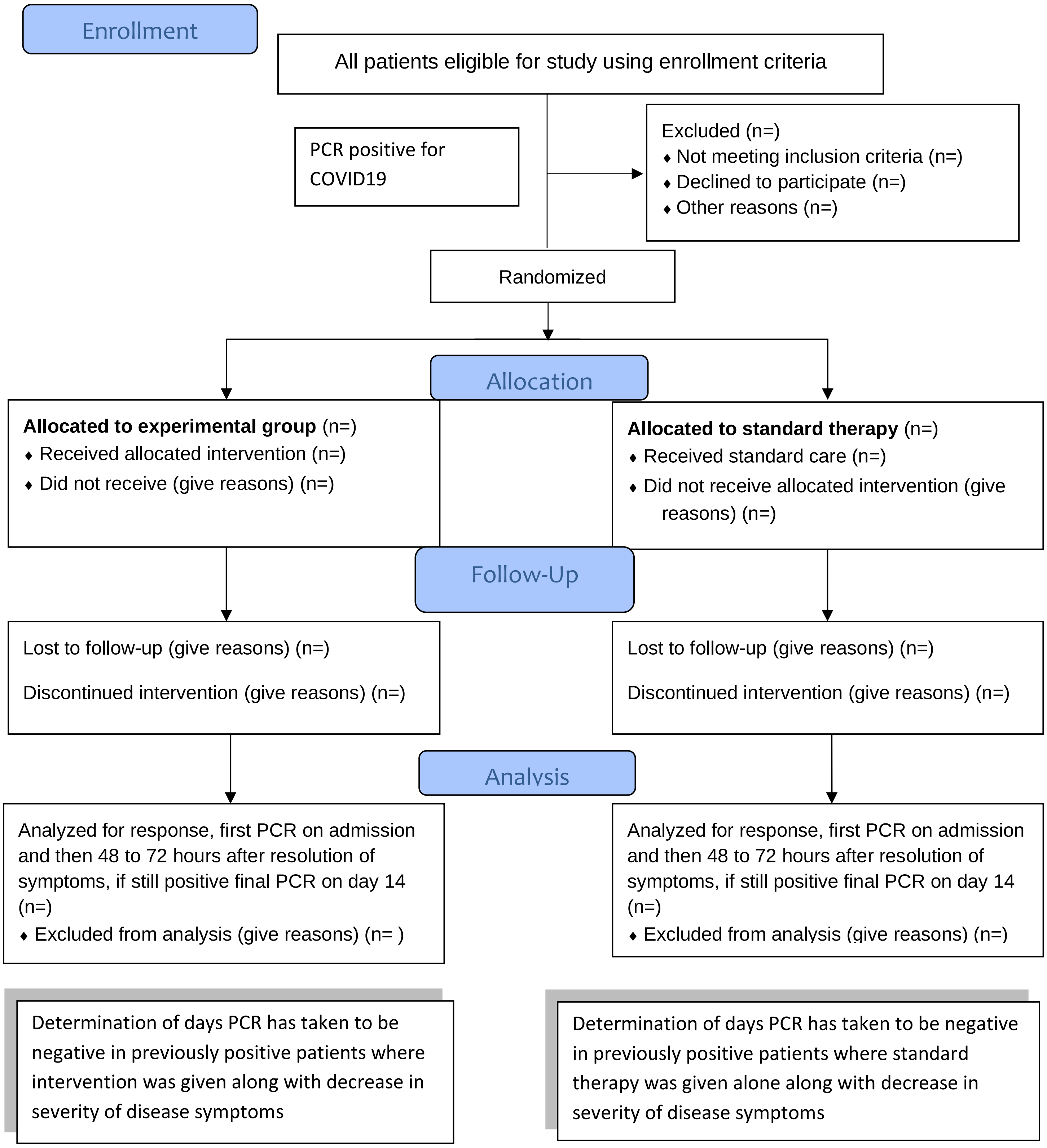

**SSC-2 (Shaikh Zayed SARS-CoV-2) SCORE**

| | |
|---|---|
| High grade Fever (without any other evident source of infection) | 5 |
| Low grade Fever (without any other evident source of infection) | 3 |
| Fever (with evident source of infection) | 2 |
| Fatigue/Myalgia | 2 |
| Dry Cough | 2 |
| Dyspnea | 3 |
| Any Atypical Symptom* | 1 |
| ARDS ` | 5 |
| Any other diagnosis unlikely | 2 |

**Score of less than 5:** Low probability

**Score of between 5 &10:** Moderate probability (need PCR to label as Corona infected Patient)

**Score of more than 10:** High probability

*(Headache, Anorexia, Sputum production, Pharyngitis, GI disturbances)

Figure 2: A score based on prevalence of symptoms reported to diagnose/ suspect patients with corona as per reported in multiple studies in COVID-19 patients

#### SUMMARY OF PROTOCOL CHANGES

Role of honey and *Nigella sativa* in the management of COVID-19 (HNS-COVID-PK): A randomized controlled trial

**SUMMARY OF CHANGES**

Protocol Version 1.0-3.0

(All changes were suggested by trial executive and steering committee which then were approved by ethical review board of Shaikh Zayed hospital Lahore, Pakistan)

**Key Changes:**

1. The introduction was modified and updated based upon recent knowledge on COVID-19, honey and *Nigella sativa*.
2. Change in methodology regarding every 4$^{th}$ day RT-PCR was made and it was switched to first PCR on admission date and second 48 to 72 hours after the patient becomes asymptomatic. It was due to following reasons:
   - Limited number of kits available for tests.
   - More exposure risk of health professionals.
   - Updated clinical literature indicated that it was not an absolute indicator of infectivity.
   - Poor socio-economic status of majority of the patients.
3. HRCT was removed in the final version of the protocol as it was observed during pilot study that odds of infectivity along with radiation exposure were increased. Furthermore, patients were quite sick and their transport to the HRCT room was an ethical concern. In the original version, HRCT was also the part of SSC-2 score, eligibility and recruitment criteria which was removed due to above mentioned reasons.
4. A well-constructed written informed consent form was created according to WHO and NIH guidelines and site investigators were made responsible for this informed consent.
5. In final version the protocol more details and strategies were added to ensure compliance and adherence.
6. The questionnaire was updated and modified considering the important and missing parameters.

**Original Statistical Analysis Plan:**

Primary and secondary outcomes will be measured as per protocol. Biostatisticians using SAS software 9.4 will analyze all trial data. Mean ± S.D will be used for quantitative data and f (%) will be used for categorical data. Frequency and percentages will be measured for categorical data. Data normality will be checked using Shapiro Wilks test, if data is normal independent sample t-test will be used to compare quantitative outcome otherwise Mann Whitney U test will be used to compare median of these quantitative data. Long rank test will be used for univariate analysis of categorical time to event data. Measurement of association using chi-square will be estimated for SARS-CoV-2 RT-PCR done every $4^{th}$ day. Repeated measured design using analysis of variance and chi square will be used for HRCT being done at $4^{th}$ day. For follow up analysis Wilcoxon test will be applied. If data will support the necessary assumptions of time to event data, survival analysis/ Kaplan Meier test will be applied. Multivariate regression models will be adjusted for the effects of age (<40 or >=40), gender, baseline clinical status grade, history of diabetes/hypertension and oxygen use. Mixed Ordinal Logistic Regression Model will be used to access ordinal outcomes and cox proportional hazards models will be used to compare time taken for alleviation of symptoms. Fisher's Exact test will be applied to compare mortality. P-value ≤ 0.05 will be considered as significant.

| **Outcome** | **Hypothesis** | **Outcome measure** | **Method of analysis** |
|---|---|---|---|
| **PRIMARY OUTCOMES** | | | |
| 1. RT-PCR (every $4^{th}$ day) | Earlier RT-PCR negative | Positive/ Negative | Measurement of association using chi square |
| 2. HRCT Chest<br>(every $4^{th}$ day) | Improvement in HRCT chest | Lungs divided into five lobes and each lobe is given 1 number. Total score is 25. Each lobe is scored from 0 to 5 as:<br>0 = no involvement<br>1 = <5% involvement<br>2 = 25% involvement<br>3 = 26%-49% involvement<br>4 = 50%-75% involvement<br>5 = >75% involvement | Repeated measured design using analysis of variance<br><br>Chi square test |

| | | | |
|---|---|---|---|
| 3. Clinical Grading Score (CGS) at Day 6 | Lower CGS score on seven pointer ordinal scale at day 6 | Grade from 1-7<br>Grade 1. Not hospitalized with resumption of normal daily life activities<br>Grade 2. Not hospitalized but unable to resume normal daily life activities<br>Grade 3. Hospitalized, but do not require supplemental oxygen therapy<br>Grade 4. Hospitalized but require low flow supplemental oxygen therapy<br>Grade 5. Hospitalized, requiring high flow supplemental oxygen<br>Grade 6. Hospitalized, requiring mechanical ventilation<br>Grade 7. Death | Mixed Ordinal Logistic Regression Model |
| 4. Time Taken for Alleviation of Symptoms | Reduction in symptomatic phase | Number of days patient remain admitted | Cox proportional hazards models |
| **SECONDARY OUTCOMES** | | | |
| 1. Degree of fever at day 4 | Lower degree of fever at day 4 | Yes/No | Mixed Ordinal Logistic Regression Model |
| 2. CRP at Day 6 | Lower CRP levels at day 6 | Yes/No | T-Test |
| 3. Severity of symptoms at day 8 | Lower degree of severity at day 8 | Severity of symptoms is classified as mild, moderate or severe. | Mixed Ordinal Logistic Regression Model |
| 4. Clinical Grading Score (CGS) at Day 10 | Lower CGS score on seven pointer ordinal scale at day 10 | Grade from 1-7<br>Grade 1. Not hospitalized with resumption of normal daily life activities<br>Grade 2. Not hospitalized but unable to resume normal daily life activities | Mixed Ordinal Logistic Regression Mode |

| | | | |
|---|---|---|---|
| | | Grade 3. Hospitalized, but do not require supplemental oxygen therapy<br>Grade 4. Hospitalized but require low flow supplemental oxygen therapy<br>Grade 5. Hospitalized, requiring high flow supplemental oxygen<br>Grade 6. Hospitalized, requiring mechanical ventilation<br>Grade 7. Death | |
| 5. 30 Day Mortality | Lower mortality | Yes/No | Fisher's Exact test |
| **ADDITIONAL OUTCOMES** | | | |
| **Median time to clinical improvement of severity of symptoms** | | | |
| Improvement of one category on ordinal scale | Earlier improvement in severity of symptoms | 1. None<br>2. Mild<br>3. Moderate<br>4. Severe | Univariate Log-rank test |
| Improvement of two category on ordinal scale | | | |
| Achievement of normal status on ordinal scale | | | |
| **Median time to clinical improvement of degree of fever** | | | |
| Improvement of one category on ordinal scale | Earlier improvement in degree of fever | 1. None: <99 °F<br>2. Mild: 99-100 °F<br>3. Moderate: 100-101.9 °F<br>4. Severe: ≥ 102°F | Univariate Log-rank test |
| Improvement of two category on ordinal scale | | | |
| Achievement of normal status on ordinal | | | |

| | | | |
|---|---|---|---|
| scale | | | |
| **Median time to clinical improvement of cough** | | | |
| Improvement of one category on ordinal scale | Earlier improvement in degree of cough | 1. None: No cough.<br>2. Mild: Occasional, transient cough<br>3. Moderate: Frequent cough, slightly influencing day time activities<br>5. Severe: Frequent cough, significantly influencing daytime activities | Univariate Log-rank test |
| Improvement of two category on ordinal scale | | | |
| Achievement of normal status on ordinal scale | | | |
| **Median time to clinical improvement of shortness of breath** | | | |
| Improvement of one category on ordinal scale | Earlier improvement in degree of shortness of breath | Grade1= Not troubled by breathlessness except on strenuous exercise<br>Grade 2= Short of breath when hurrying on the level or walking up a slight hill<br>Grade 3= Walks slower than most people on the level, stops after a mile or so, or stop after 15 minutes walking at own pace<br>Grade 4= Stops for breath after walking about 100yds or a few minutes on level ground.<br>Grade 5= Too breathless to leave the house, or breathless when undressing | Univariate Log-rank test |
| Improvement of two category on ordinal scale | | | |
| Achievement of normal status on ordinal scale | | | |
| **Median time to clinical improvement of myalgia** | | | |
| Improvement of one category on ordinal scale | Earlier improvement in degree of myalgia | 1. None | Univariate Log-rank test |

| | | | |
|---|---|---|---|
| Improvement of two category on ordinal scale | | 2. Mild<br>3. Moderate<br>4. Severe | |
| Achievement of normal status on ordinal scale | | | |
| **Median time to clinical improvement of "how sick do you feel"** | | | |
| Improvement of one category on ordinal scale | Earlier improvement in status "How sick do you feel" | 1. None<br>2. Mild<br>3. Moderate<br>4. Severe | Univariate Log-rank test |
| Improvement of two category on ordinal scale | | | |
| Achievement of normal status on ordinal scale | | | |

**Final Statistical Analysis Plan:**

Primary and secondary outcomes will be measured as per protocol. Biostatisticians using SAS software 9.4 will analyze all trial data. Mean ± S.D will be used for quantitative data and f (%) will be used for categorical data. Frequency and percentages will be measured for categorical data. Data normality will be checked using Shapiro Wilks test, if data is normal independent sample t-test will be used to compare quantitative outcome otherwise Mann Whitney U test will be used to compare median of these quantitative data. Long rank test will be used for univariate analysis of categorical time to event data. For follow up analysis Wilcoxon test will be applied. If data will support the necessary assumptions of time to event data, survival analysis/ Kaplan Meier test will be applied. Multivariate regression models will be adjusted for the effects of age (<40 or >=40), gender, baseline clinical status grade, history of diabetes/hypertension and oxygen use. Mixed Ordinal Logistic Regression Model will be used to access ordinal outcomes and cox proportional hazards models will be used to compare time taken for alleviation of symptoms and viral clearance. Fisher's Exact test will be applied to compare mortality. P-value ≤ 0.05 will be considered as significant.

| **Outcome** | **Hypothesis** | **Outcome measure** | **Method of analysis** |
|---|---|---|---|
| **PRIMARY OUTCOMES** | | | |
| 1. Time Taken for Alleviation of Symptoms | Earlier resolution of symptoms | Yes/No | Multivariant analysis, cox proportional hazards models |
| 2. Viral Clearance | Earlier RT-PCR negative | Positive/ negative | |
| 3. Clinical Grading Score (CGS) at Day 6 | Lower CGS score on seven pointer ordinal scale at day 6 | Grade from 1-7<br>Grade 1. Not hospitalized with resumption of normal daily life activities<br>Grade 2. Not hospitalized but unable to resume normal daily life activities<br>Grade 3. Hospitalized, but do not require supplemental oxygen therapy<br>Grade 4. Hospitalized but require low flow supplemental oxygen therapy<br>Grade 5. Hospitalized, requiring high flow supplemental oxygen<br>Grade 6. Hospitalized, requiring mechanical ventilation<br>Grade 7. Death | Mixed Ordinal Logistic Regression Model |
| **SECONDARY OUTCOMES** | | | |

| | | | |
|---|---|---|---|
| 1. Degree of fever at day 4 | Lower degree of fever at day 4 | Yes/No | Mixed Ordinal Logistic Regression Model |
| 2. CRP at Day 6 | Lower CRP levels at day 6 | Yes/No | Linear regression model |
| 3. Severity of symptoms at day 8 | Lower degree of severity at day 8 | Severity of symptoms is classified as mild, moderate or severe. | Mixed Ordinal Logistic Regression Model |
| 4. Clinical Grading Score (CGS) at Day 10 | Lower CGS score on seven pointer ordinal scale at day 10 | Grade from 1-7<br>Grade 1. Not hospitalized with resumption of normal daily life activities<br>Grade 2. Not hospitalized but unable to resume normal daily life activities<br>Grade 3. Hospitalized, but do not require supplemental oxygen therapy<br>Grade 4. Hospitalized but require low flow supplemental oxygen therapy<br>Grade 5. Hospitalized, requiring high flow supplemental oxygen<br>Grade 6. Hospitalized, requiring mechanical ventilation<br>Grade 7. Death | Mixed Ordinal Logistic Regression Mode |
| 5. 30 Day Mortality | Lower mortality | Yes/No | Fisher’s Exact test |
| **ADDITIONAL OUTCOMES** | | | |
| **Median time to clinical improvement of severity of symptoms** | | | |
| Improvement of one category on ordinal scale | Earlier improvement in severity of symptoms | 1. None<br>2. Mild<br>3. Moderate<br>4. Severe | Univariate Log-rank test |
| Improvement of two category on ordinal scale | | | |
| Achievement of normal status on ordinal scale | | | |

| | | | |
|---|---|---|---|
| **Median time to clinical improvement of degree of fever** | | | |
| Improvement of one category on ordinal scale | Earlier improvement in degree of fever | 1. None: <99 °F<br>2. Mild: 99-100 °F<br>3. Moderate: 100-101.9 °F<br>4. Severe: ≥ 102°F | Univariate Log-rank test |
| Improvement of two category on ordinal scale | | | |
| Achievement of normal status on ordinal scale | | | |
| **Median time to clinical improvement of cough** | | | |
| Improvement of one category on ordinal scale | Earlier improvement in degree of cough | 1. None: No cough.<br>2. Mild: Occasional, transient cough<br>3. Moderate: Frequent cough, slightly influencing day time activities<br>5. Severe: Frequent cough, significantly influencing daytime activities | Univariate Log-rank test |
| Improvement of two category on ordinal scale | | | |
| Achievement of normal status on ordinal scale | | | |
| **Median time to clinical improvement of shortness of breath** | | | |
| Improvement of one category on ordinal scale | Earlier improvement in degree of shortness of breath | Grade1= Not troubled by breathlessness except on strenuous exercise<br>Grade 2= Short of breath when hurrying on the level or walking up a slight hill<br>Grade 3= Walks slower than most people on the level, stops after a mile or so, or stop after 15 minutes walking at own pace<br>Grade 4= Stops for breath after walking about 100yds or a few minutes on level ground. | Univariate Log-rank test |
| Improvement of two category on ordinal scale | | | |
| Achievement of normal status on ordinal scale | | | |

| | | | |
|---|---|---|---|
| | | Grade 5= Too breathless to leave the house, or breathless when undressing | |
| **Median time to clinical improvement of myalgia** | | | |
| Improvement of one category on ordinal scale | Earlier improvement in degree of myalgia | 1. None<br>2. Mild<br>3. Moderate<br>4. Severe | Univariate Log-rank test |
| Improvement of two category on ordinal scale | | | |
| Achievement of normal status on ordinal scale | | | |
| **Median time to clinical improvement of "how sick do you feel"** | | | |
| Improvement of one category on ordinal scale | Earlier improvement in status "How sick do you feel" | 1. None<br>2. Mild<br>3. Moderate<br>4. Severe | Univariate Log-rank test |
| Improvement of two category on ordinal scale | | | |
| Achievement of normal status on ordinal scale | | | |

**Summary of changes in statistical analysis plan:**

Following changes were made in statistical analysis plan

- The statistical analysis plan was changed from repeated measurements of association using chi-square to cox proportional hazards models. It was done due to change in outcome methodology from every $4^{th}$ day RT-PCR to $1^{st}$ PCR followed by $2^{nd}$ PCR (48 to 72 hours after the patient becomes asymptomatic).

- The statistical analysis plan was removed for HRCT as it was removed as an endpoint due to ethical and practical concerns.

### CONSENT FORM

This consent form is designed to give information regarding the trial and to obtain a well-informed consent of the enrolled patients.

**Title**: Role of honey and *Nigella sativa* in the management of COVID-19 (HNS-COVID-PK): A randomized controlled trial

**Principal Investigator**: Dr. Sohaib Ashraf

**Name of Organization**: Shaikh Zayed Medical Complex Lahore and Services Institute of Medical Sciences

The Consent Form is divided into two parts:

• Part 1, gives you a brief introduction about the trial

• Part 2, is the Consent Form

**PART I:**

**Introduction:**

I am _________________________ our team is conducting a research on COVID-19, which is a global health concern. I am highlighting all the relevant information and inviting you to be part of this trail. Prior to any decision concerning your participation, take your time and talk to someone you feel at ease with about this research. If you have any difficulty understanding anything, you may stop me, and I will try my best to explain as I walk you through the information. If you still have any queries, you can clear them with the relevant doctor or the staff. A copy of this form will also be given to you.

**Explanation**:

The proposed study is a placebo-controlled, add-on, randomized trial using parallel group designs. This is an open-label and adaptive, multi-centered design with 1:1 allocation ratio and superiority framework. This study will have two arms interventional arm will be Honey and *Nigella sativa* while other arm will be standard care with placebo. Data will be collected on self-constructed, close-ended questionnaires after obtaining written consent. Data will be analyzed using SAS version 9.4.

COVID-19 positive patients will be designated via qRT-PCR. Primary and secondary outcomes will be analyzed in both groups. The trial will aid in devising a better strategy to cope COVID-19 in a relatively inexpensive and accessible. The implications are global, and this could prove itself to be the most manageable intervention against COVID-19 especially for patients from limited-resource countries with deprived socioeconomic statuses.

**Voluntary Participation**:

It's a voluntary participation in this trial. Whether you choose to be a part of it or not, all the services you receive at this hospital will continue and nothing will change. If you choose to participate you will have the authority to change your mind later and stop participating even if you agreed earlier.

**Procedures and Protocol**

Participants in one group will be given the experimental treatment along with the standard treatment while participants in the other group will only be given the standard treatment as per hospital protocol. The healthcare workers will be monitoring you and the other volunteers' vigilantly during the study. If there is anything you are anxious about or that is troubling you about the research, please talk to me or one of the other colleagues. For this study we will be withdrawing 10ml of your blood (when needed), which will help us access your clinical laboratory data. Blood will be withdrawn from your arm, via trained staff, using a syringe through arterial/venous site. Furthermore, a baseline X-ray chest would also be performed.

You will not be monetarily benefited to take part in this research and confidentiality will be maintained. Identity of those taking part in this trial will not be revealed. The personal information that we collect from this research project will be kept confidential. Your personal information will be given numbers instead of your names. Only the researchers will be aware of the assigned number and we will lock that information up with a lock and key. This information can only be accessed by study director and study chair.

**Sharing the Results:**

Prior to making the knowledge, we get from this research, publically available to the outside world; it will be shared with the participants through community/zoom meetings. Confidential information will not be shared. After these meetings, we will publish the results so that the knowledge gained can be shared with rest of the world.

**Right to Refuse or Withdraw:**

You do not have to take part in this research if you do not wish to do so. You may also stop participating in the research at any time you choose. It is your choice and all of your rights will still be respected. Alternatives to Participating If you do not wish to take part in the research, you will be provided with the established standard treatment available at the center/institute/hospital.

**Who to Contact:**

If you have any questions you may ask them now or later, even after the study has started. If you wish to ask questions later, you may contact any of the following:

Dr Sohaib Ashraf +923334474523,

Dr Muhammad Ahmad Imran +923338110708

Dr Shahroze Arshad +923244516332

This proposal has been reviewed and approved by ethical review board of Shaikh Zayed Medical Complex, Lahore, (**SZH/IRB/0026/2020**) and Services Institute of Medical Sciences, Lahore(**IRB/2020/658/SIMS**) which is a committee whose task it is to make sure that research participants are protected from harm. You can ask me any more questions about any part of the research study if you wish to.

Do you have any questions?

**PART II:**

I have read the foregoing information, or it has been read to me. I have had the opportunity to ask questions about it and any questions that I have asked have been answered to my satisfaction. I consent voluntarily to participate as a participant in this research.

Print Name of Participant__________________

Signature of Participant ___________________

Date ___________________________ Day/month/year

If Illiterate (consent form being read to the witness) I have witnessed the accurate reading of the consent form to the potential participant, and the individual has had the opportunity to ask questions. I confirm that the individual has given consent freely.

Print name of witness_____________________

Thumb print of participant/ Signature of witness ______________________

Date ________________________ Day/month/year

I have accurately read out the information sheet to the potential participant, and to the best of my ability made sure that the participant understands. I confirm that the participant was given an opportunity to ask questions about the study, and all the questions asked by the participant have been answered correctly and to the best of my ability.

I confirm that the individual has not been coerced into giving consent, and the consent has been given freely and voluntarily.

A copy of this ICF has been provided to the participant.

Name of the person taking the consent ________________________

Signature of Researcher /person taking the consent__________________________

Date ___________________________ Day/month/year

**QUESTIONAIRE**

DEMOGRAPHIC INFORMATION:

Medical Record #: ____________________________ ID: ___________________________

Name (optional): ___________________________________________________________

Age: ___________ Gender: Male / Female

Contact Number: __________________

Oxygen Saturation: __________________

Profession: _______________________

Study Center: ______________________

Blood Group: _____________________

Date of admission: __________________

Date of 1st PCR: ____________________

Date of symptom resolution: __________

Date of 2nd PCR: ___________________

Fever: __________

Anorexia: ______

GI disturbances: _______

Fatigue: ______

Sputum production: ____

Severity of Symptoms: __________

Dry Cough: __________

Sore throat: ______

Loss of smell: _______

SOB: ___________

Myalgia: _______

Change in taste: _______

CBC: ____________________________________________________________________

RFTs: ___________________________________________________________________

LFTs: ___________________________________________________________________

ECG: ____________________________________________________________________

CRP day 0: _______

CRP day 6: _______

Chest X-ray: ____________________

ARDS: _____

**Clinical Course of Treatment:**

Vital Signs:

Heart rate: ________ Temperature: ________ Respiratory Rate: ________ Systolic BP: ________ Diastolic BP: ________

| Day | Fever | Cough | Shortness of Breath | Symptoms | How sick do you feel | Rate Your Emotional Status | Myalgia | Oxygen Saturation at Room Air | Oxygen Therapy | Additional Medicine |
|---|---|---|---|---|---|---|---|---|---|---|
| **0** | | | | | | | | | | |
| **1** | | | | | | | | | | |
| **2** | | | | | | | | | | |
| **3** | | | | | | | | | | |
| **4** | | | | | | | | | | |
| **5** | | | | | | | | | | |
| **6** | | | | | | | | | | |
| **7** | | | | | | | | | | |
| **8** | | | | | | | | | | |
| **9** | | | | | | | | | | |
| **10** | | | | | | | | | | |
| **11** | | | | | | | | | | |
| **12** | | | | | | | | | | |
| **13** | | | | | | | | | | |

**Clinical Grading Scale:**

Grade the clinical status of patient 0, 4, 6, 8, 10 and 12. Choose the appropriate.

| Grade | Day 0 | Day 4 | Day 6 | Day 8 | Day 10 | Day 12 |
|---|---|---|---|---|---|---|
| Grade 1. Not hospitalized with resumption of normal daily life activities | | | | | | |
| Grade 2. Not hospitalized but unable to resume normal daily life activities | | | | | | |
| Grade 3. Hospitalized, but do not require supplemental oxygen therapy | | | | | | |
| Grade 4. Hospitalized but require low flow supplemental oxygen therapy | | | | | | |
| Grade 5. Hospitalized, requiring high flow supplemental oxygen | | | | | | |
| Grade 6. Hospitalized, requiring mechanical ventilation | | | | | | |
| Grade 7. Death | | | | | | |
