## Supplementary Appendix for "Honey and *Nigella sativa* against COVID-19 in Pakistan (HNS-COVID-PK): A multi-center placebo-controlled randomized clinical trial"

**TABLE OF CONTENT:**


| **Contents** | **Page Number** |
|---|---|
| Team of investigators | 3 |
| Trial executive committee: | 7 |
| Trial steering committee | 9 |
| Additional details on participants | 10 |
| Figure S1: Enrolment criteria | 11 |
| Figure S2: SSC-2 (Shaikh Zayed SARS-CoV-2) SCORE | 12 |
| Additional details on randomization | 13 |
| Additional details on interventions | 13 |
| Additional details on outcomes | 14 |
| Figure S3: Participant timeline | 18 |
| Additional details on statistical analyses | 21 |
| Table S1. Baseline table | 26 |
| Table S2. Distribution of severity of cough over days | 31 |
| Table S3. Distribution of Severity of Symptoms over days | 32 |
| Table S4. Distribution of fever over days | 33 |
| Table S5. Distribution of Myalgia over days | 34 |
| Table S6. Distribution of Shortness of Breath over days | 35 |
| Table S7. Variation of oxygen saturation over days | 36 |
| Table S8. Distribution of How sick do you feel over days | 37 |
| Table S9 Variation of emotional status over days | 38 |
| Table S10 Variation of clinical status grading over days | 40 |

### TEAM OF INVESTIGATORS

#### Study Chair:

**Prof Muhammad Ashraf, PhD (University of Minnesota)**

Institutional Affiliation: Department of Pharmacology and Toxicology, University of Veterinary & Animal Sciences, Lahore, Pakistan

#### Study Director:

**Shoaib Ashraf, PhD (McGill University)**

Institutional Affiliation: Wellman center for photomedicine, Massachusetts General Hospital, Harvard University, Boston, USA

#### Study Principal Investigators:

1. **Moneeb Ashraf, MBBS, M.Phil. (Pharmacology).**
   Institutional Affiliation: Department of Pharmacology, King Edward Medical University, Mayo Hospital, Lahore, Pakistan

2. **Sohaib Ashraf, MBBS**
   Institutional Affiliation: Department of Cardiology, Shaikh Zayed Post-Graduate Medical Complex, Lahore, Pakistan

#### Study Co-Investigators:

1. **Ali Ahmad, PhD (Cornell University)**
   Institutional Affiliation: Department of Microbiology, Infectiology and Immunology, Centre Hospitalier Universitaire (CHU) Sainte Justin/University of Montreal, Montreal, Canada.

2. **Muhammad Ahmad Imran, MBBS**
   Institutional Affiliation: Department of Microbiology, Shaikh Zayed Post-Graduate Medical Complex, Lahore, Pakistan

#### Principal Investigator (SZH):

1. **Prof Mateen Izhar, MBBS, PhD, FRCPath**
   Institutional Affiliation: Department of Microbiology, Shaikh Zayed Medical Complex, Lahore, Pakistan

2. **Prof Ayesha Humayun, MBBS, PhD**
   Institutional Affiliation: Department of Community Medicine, Shaikh Zayed Medical Complex, Lahore, Pakistan

#### Co-Investigators (SZH):

1. **Prof Qazi Abdul Saboor, MBBS, FCPS (Cardiology)**
   Institutional Affiliation: Department of Cardiology, Shaikh Zayed Medical Complex, Lahore, Pakistan
2. **Prof Talha Mahmood, MBBS, MD, FCPS (Pulmonology)**
   Institutional Affiliation: Department of Pulmonology, Shaikh Zayed Medical Complex, Lahore, Pakistan
3. **Uzma Nasim Siddiqui, MBBS, FCPS (Medicine)**
   Institutional Affiliation: Department of Medicine, Shaikh Zayed Medical Complex, Lahore, Pakistan

#### Site Investigators (SZH):

1. Abubakar Hilal, MBBS, FCPS
2. Ammara Ahmad, MBBS
3. Usama Farooq, MBBS
4. Saif-ul-wahab, MBBS
5. Ansar Maqbool, MBBS
6. Zeeshan Shaukat, MD
7. Umair Hafeez, MBBS
8. Tayyab Mughal, MBBS
9. Ayesha Khaqan, MBBS
10. Qurrat-ul-ain Iqbal, MBBS

11. Hassan Riaz, MBBS
12. Kanwal Hayat, BSc (Nursing)
13. Ghazala BSc(Nursing)
14. Mehak Gul, BSc (Nursing)

#### Principal Investigator (SIMS):

1. **Prof Mahmood Ayyaz, MBBS, FCPS, FRCSEd, FICS, FACS**

   Institutional Affiliation: Chairman, Services Institute of Medical Sciences, Lahore, Pakistan.

#### Co-Investigators (SIMS):

1. **Nighat Majeed, MBBS, FCPS(Medicine)**

   Institutional Affiliation: Department of Medicine, Services Hospital, Lahore, Pakistan
2. **Larab Kalsoom, MBBS**

   Institutional Affiliation: Department of Medicine, Services Hospital, Lahore, Pakistan

#### Site Investigators (SIMS):

1. Iqra Farooq, MBBS
2. Abeer Bin Awais, MBBS
3. Hassan Mujtaba Cheema, MBBS
4. Abdur Rehman Virk, MBBS
5. Mehak Gul, MBBS
6. Ammara Ahmad, MBBS
7. Rashida Sharif, BSc (Nursing)

#### Investigators (Ali Clinic):

1. Zaighum Habib, MBBS, MS
2. Zain-ul-Abdin, MBBS
3. Shahroz Arshad, MBBS
4. Misbah Kousar, (BSc Nursing)

#### Investigators (Doctors Lounge):

1. Sundas Rafique, MBBS
2. Suneela Ahmad, MBBS
3. Sarmad Shahab, MBBS
4. Sumaira Hamid, BSc (Nursing)

**LEAD BIOSTATISTICIANS:**


**Prof Muhammad Azam, PhD**

Institutional Affiliation: University of Veterinary & Animal Sciences, Lahore, Pakistan

**Zheng Hui, PhD**

Institutional Affiliation: Massachusetts General Hospital, Boston, MA, USA

Harvard Medical School, Boston, MA, USA

**Sohail Ahmad, PhD**

Institutional Affiliation: University of Veterinary & Animal Sciences, Lahore, Pakistan

**Muhammad Kiwan Akram, M.Phil.**

Institutional Affiliation: University of Veterinary & Animal Sciences, Lahore, Pakistan


**RESEARCH PHARMACISTS & DRUG SAFETY PANELISTS:**


1. **Syed Muhammad Muneeb**, **PharmD, PhD**
   Institutional Affiliation: Institute of Pharmaceutical Sciences, University of Veterinary & Animal Sciences, Lahore, Pakistan
2. **Muhammad Faisal Nadeem, PharmD, PhD**
   Institutional Affiliation: Institute of Pharmaceutical Sciences, University of Veterinary & Animal Sciences, Lahore, Pakistan

**TRIAL EXECTIVE COMMITTEE:**

- **Prof Muhammad Ashraf, PhD (University of Minnesota)**
  Institutional Affiliation: University of Veterinary & Animal Sciences, Lahore, Pakistan
- **Shoaib Ashraf, PhD (McGill University)**
  Institutional Affiliation: Riphah College of Veterinary Sciences, Riphah University, Lahore, Pakistan.
- **Sidra Ashraf, M.Phil.**
  Institutional Affiliation: University of Veterinary & Animal Sciences, Lahore, Pakistan
- **Moneeb Ashraf, MBBS, M.Phil. (Pharmacology).**
  Institutional Affiliation: Kingeward Medical University, Mayo Hospital, Lahore Pakistan
- **Sohaib Ashraf, MBBS**
  Institutional Affiliation: Shaikh Zayed Medical Complex, Lahore, Pakistan
- **Prof Ali Ahmad, PhD**
  Institutional Affiliation: University of Montreal, Sainte Justin Hospital, Montreal, Canada.
- **Prof Mateen Izhar, MBBS, PhD, FRCPath**
  Institutional Affiliation: Shaikh Zayed Medical Complex, Lahore, Pakistan
- **Prof Mahmood Ayyaz MBBS, FCPS, FRCS, FACS**
  Institutional Affiliation: Services Institute of Medical Sciences, Lahore, Pakistan.
- **Prof Ayesha Humayun, MBBS, FCPS, PhD**
  Institutional Affiliation: Shaikh Khalifa bin Zayed Al-Nahyan Medical & Dental College, Lahore, Pakistan
- **Prof Talha Mahmood, MBBS, FCPS, MD (Pulmonology)**
  Institutional Affiliation: Shaikh Zayed Medical Complex, Lahore, Pakistan
- **Prof Qazi Abdul Saboor, MBBS, FCPS (Cardiology)**
  Institutional Affiliation: Shaikh Zayed Medical Complex, Lahore, Pakistan
- **Nighat Masood, MBBS, FCPS (Medicine)**
  Institutional Affiliation: Services Institute of Medical Sciences, Lahore, Pakistan.
- **Uzma Nasim Siddique, MBBS, FCPS (Medicine)**
  Institutional Affiliation: Shaikh Zayed Medical Complex, Lahore, Pakistan

- **Sundas Rafique, MBBS**

  Institutional Affiliation: Kingedward Medical Institute, Mayo Hospital, Lahore, Pakistan
- **Zain-ul-abdin, MBBS**

  Institutional Affiliation: Shaikh Zayed Hospital, Lahore, Pakistan
- **Muhammad Ahmad Imran, MBBS**

  Institutional Affiliation:  Federal Post-Graduate Medical Institute, Lahore, Pakistan.
- **Larab Kalsoom, MBBS**

  Institutional Affiliation: Services Hospital, Lahore, Pakistan
- **Iqra Farooq, MBBS**

  Institutional Affiliation: Children Hospital, Lahore, Pakistan
- **Muhammad Kiwan Akram, M.Phil. (Nutrition)**

  Institutional Affiliation: UVAS, Lahore, Pakistan
- **Kanwal Hayat, BSc (Nursing)**

  Institutional Affiliation: Shaikh Zayed Hospital, Lahore, Pakistan

**TRIAL STEERING COMMITTEE**

- **Muhammad Imran, PhD**
  Institutional Affiliation: Massachusetts Institute of Technology, Boston, USA
- **Yasir Zahoor, PhD**
  Institutional Affiliation: Boston Children Hospital, Harvard Medical School, USA
- **Umer Naveed, PhD**
  Institutional Affiliation: Royal (Dick) School of Veterinary Studies and Roslin Institute, Scotland, UK
- **Fazli Mabood, PhD**
  Institutional Affiliation: CHU Sainte-Justine Hospital, University of Montreal, Canada
- **Muhammad Bilal, M.Phil.**
  Institutional Affiliation: McGill University, Sainte-Anne-de-Bellevue, Quebec, Canada
- **Syed Kazim Ali, LLB**
  Institutional Affiliation: Lahore High Court Bar, Pakistan
  United Nations for Human Rights
- **Asad-ullah, Pharm.D, M.Phil.**
  Institutional Affiliation: Drug Regulation Authority Pakistan (DRAP)
- **Zaka-ur-Rehman, PhD**
  Institutional Affiliation: Ministry of Health Punjab, Pakistan.

### Supplemental Methods

#### Additional Details on Participants

All COVID-19 patients presented in study settings of Pakistan during the study period were enrolled.

<u>Inclusion Criteria:</u>

- All suspected patients with an SSC-2 score (Figure 2) fulfilling the enrolment criteria (Figure 1)
- Positive RT-PCR (Real Time Polymerase Chain Reaction)
- Patients who presented to seek medical care within 96 hours of ailment/randomization with positive RT-PCR and normal baseline laboratory parameters
- Varying degree of COVID-19 disease severity
- Both genders with age 18 years and above

<u>Exclusion Criteria:</u>

- Presence of co-morbidities like liver disease, immuno-compromised patient or any other chronic ailment (other than Diabetes and Hypertension)
- Females who are pregnant and breast feeding
- Uncontrolled Diabetes Mellitus and Hypertension
- If a patient is allergic to either honey or *Nigella sativa*.
- Refusal or withdrawal from informed consent.
- patient on ventilator support or PaO2/FIO2 of less than 100
- Hemodynamic instability
- Patients with positive SARS-CoV-2 screening during elective lists for any procedure were also excluded.

All suspected patients were assessed on SSC-2 score. If a score is less than 5, the patient was excluded from the study. If the score was ≥5, SARS-CoV-2 RT-PCR performed, if first RT-PCR negative, patient was excluded but if PCR was positive, patient included. (This decision is made on first RT-PCR).

**Enrolment Criteria:**

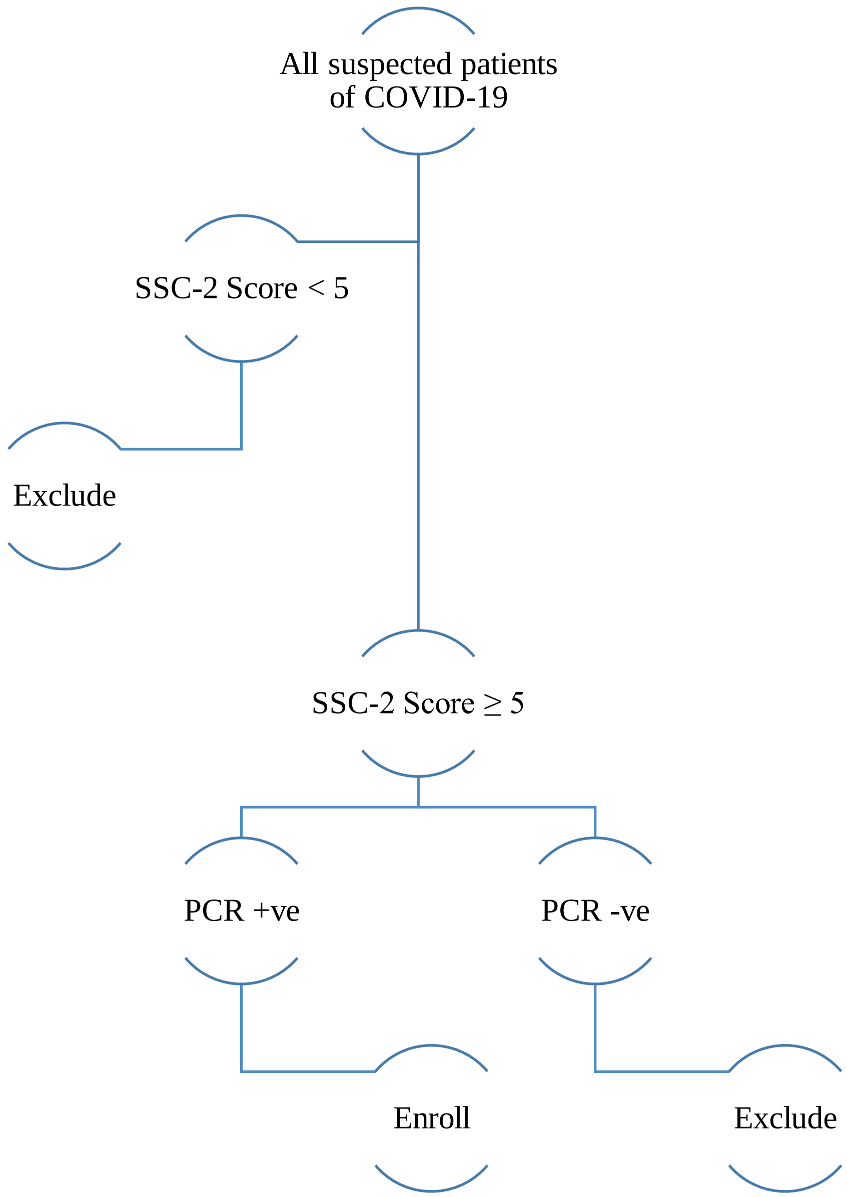


**Figure S1:** Enrollment criteria for suspected COVID-19 patients.

#### SSC-2 (Shaikh Zayed SARS-CoV-2) SCORE

| | |
|---|---|
| High grade Fever (without any other evident source of infection) | 5 |
| Low grade Fever (without any other evident source of infection) | 3 |
| Fever (with evident source of infection) | 2 |
| Fatigue/Myalgia | 2 |
| Dry Cough | 2 |
| Dyspnea | 3 |
| Any Atypical Symptom* | 1 |
| ARDS ` | 5 |
| Any other diagnosis unlikely | 2 |

**Score of less than 5:** Low probability

**Score of between 5 &10:** Moderate probability (need PCR to label as Corona infected Patient)

**Score of more than 10:** High probability

*(Headache, Anorexia, Sputum production, Pharyngitis, GI disturbances)

**Figure S2:** A score based on prevalence of symptoms reported to diagnose/ suspect patients with corona as per reported in multiple studies in COVID-19 patients.

**Additional Details on the Randomization**

As a pilot project 15 patients in each arm was initiated for this multi-centered study in Pakistan. Later, this study was expanded considering the feasibility of the trial, for the 3 months from start of study (30/4/2020 to 29/7/2020). Stratification was done via SAS version 9.4 for age groups, gender, additional drugs administered, base line severity of symptoms and co-morbidities was used to ensure that groups remain balanced in size for either arm after written informed consent to participate in our study. Randomization was done using a lottery method. To minimize allocation bias, we performed allocation concealment with an interactive voice/web-based response system until randomization was finished on the system through a computer or phone. The allocation sequence was computer generated that was concealed from all site investigators, allocated participants and treatment providers until the final interventional allocation.

#### Additional details on interventions

Both arms received standard care as per advised by treating physician and "Clinical Management Guidelines for COVID-19 by Ministry of National Health Services, Pakistan", standard care administered was comprised primarily of anti-pyretic, analgesics, antibiotics, multi-vitamins, intravenous fluids, supplemental oxygen, invasive/noninvasive ventilation, immune-modulatory agents, and inotropic support. The placebo comparator group received an empty capsule as a placebo in addition to standard care while the intervention arm received honey and Nigella sativa an add-on. Nigella sativa seeds (80 mg/kg/day) were grinded and packed in capsules which was be given to the interventional group along with 1 gm/kg/day of honey stirred in 250ml of warm water in 2 to 3 divided doses, for a maximum of 13 days or the patient completely recovered. To improve adherence to the intervention, participants were counselled about the advantages of this study. All the participants were monitored regarding compliance to their assigned treatment strategy. As this trial included admitted patients and patients enrolled via telemedicine and OPD direct observational method was followed for admitted patients and structured telephone calls for 13 days was used to make sure compliance and adherence to the interventions for telemedicine and OPD enrolled patients (in case, whenever a OPD patient missed a visit in his/her follow up 13th day period).

#### Additional details on outcomes:

**Primary Outcomes:**

- Time taken for alleviation of symptoms (Date of asymptomatic-Date of randomization)
- Time taken for viral load clearance (Date of 1st Negative PCR (done once patient was 48 hours asymptomatic after treatment) -Date of 1st Positive PCR)
- Clinical status grading at Day 6 (defined by seven-point ordinal scale as per World Health Organization R&D Blueprint Ordinal Clinical Scale).

**Secondary Outcomes:**

- Degree of Fever at day 4
- CRP level measured at day 6
- Severity of symptoms at day 8
- Clinical Status grading at day 10
- 30 Day Mortality

**Additional Outcomes:**

Median time to clinical improvement of severity of symptoms, degree of fever, severity of cough, degree of shortness of breath, degree of myalgia and emotional status.

**RT-PCR:** was done on enrolment day (i.e. day 0) which was repeated once patient became asymptomatic for more than 48 to 72 hours. If the PCR would still be positive, it was repeated on day 14th. RT-PCR is shown as positive or negative.

**OPERATIONAL DEFINITION**

**Cough**

1. None: No cough.
2. Mild: Occasional, transient cough
3. Moderate: Frequent cough, slightly influencing daytime activities
4. Severe: Frequent cough, significantly influencing daytime activities

**Fever**

1. None: <99 °F
2. Mild: 99-100 °F
3. Moderate: 100-101.9 °F
4. Severe: ≥ 102°F

**Myalgia**

It was classified as mild, moderate and severe on subjective basis. Patients have been provided with a scale of 1-10, with 1 being the lowest and 10 being the highest value.

1. None: 0
2. Mild: 1-3
3. Moderate: 4-6
4. Severe: >6-10

**Shortness of Breath**

It was classified as Grade1, Grade2, Grade3, Grade4 and Grade5.

1. Grade1= Not troubled by breathlessness except on strenuous exercise
2. Grade 2= Short of breath when hurrying on the level or walking up a slight hill
3. Grade 3= Walks slower than most people on the level, stops after a mile or so, or stop after 15 minutes walking at own pace
4. Grade 4= Stops for breath after walking about 100yds or a few minutes on level ground
5. Grade 5= Too breathless to leave the house, or breathless when undressing

**How Sick Do You Feel**

It was classified as mild, moderate and severe on subjective basis. Patients have been provided with a scale of 1-10, with 1 being the highest and 10 being the lowest

1. None: 10
2. Mild: >6-9
3. Moderate: 4-6
4. Severe: 1-3

**Time Taken for alleviation of symptoms:**

Number of days taken by the patients in experimental and control groups for resolution of symptoms.

**Grading of Clinical Status:**
Grading of clinical status was assessed at 0, 4, 6, 8, 10, $12^{th}$ day considering patient condition on that moment.

Grade 1. Not hospitalized with resumption of normal daily life activities
Grade 2. Not hospitalized but unable to resume normal daily life activities
Grade 3. Hospitalized, but do not require supplemental oxygen therapy
Grade 4. Hospitalized but require low flow supplemental oxygen therapy
Grade 5. Hospitalized, requiring high flow supplemental oxygen
Grade 6. Hospitalized, requiring mechanical ventilation
Grade 7. Death

**Severity of respiratory symptoms** is classified as mild, moderate, severe and Acute Respiratory Distress Syndrome (ARDS)

1. None: Baseline health of patient.
2. Mild: "symptoms of an upper respiratory tract viral infection i.e. low-grade fever, dry cough, sore throat, nasal congestion, malaise but no radiological evidence of pneumonia"
3. Moderate: "Moderate disease includes patients with Hypoxia (Oxygen saturation <94% but >90%) or chest X-ray with infiltrates involving <50% of the lung fields or fever, cough, sputum production, and other respiratory tract or non-specific symptoms along with manifestation of pneumonia on imaging but no signs of severe pneumonia"
4. Severe: "Fever is associated with severe dyspnoea, respiratory distress, tachypnea (> 30 breaths/min), and hypoxia ($SpO_2$ < 94% on room air)" or a $PaO_2$ to $FiO_2$ ratio of 300 or lower.

| | *STUDY PERIOD* | | | | | | | | | | | | | | | |
|---|---|---|---|---|---|---|---|---|---|---|---|---|---|---|---|---|
| | **Enrolment Allocation** | **Post-allocation** | | | | | | | | | | | | | | Follow-up |
| ***TIME POINT*** | Day 0 | Day 1 | Day 2 | Day 3 | Day 4 | Day 5 | Day 6 | Day 7 | Day 8 | Day 9 | Day 10 | Day 11 | Day 12 | Day 13 | Day 14 | Day 30 |
| ***ENROLMENT:*** | | | | | | | | | | | | | | | | |
| *Eligibility screen* <br> *Randomization* | x | | | | | | | | | | | | | | | |
| *Informed consent* | x | | | | | | | | | | | | | | | |
| *Demographics* [1] | x | | | | | | | | | | | | | | | |
| *Medical history and epidemiological information* [2] | x | | | | | | | | | | | | | | | |
| ***INTERVENTIONS:*** | | | | | | | | | | | | | | | | |
| *Honey and Nigella sativa* | | x | x | x | x | x | x | x | x | x | x | x | x | x | | |
| *Placebo* | | x | x | x | x | x | x | x | x | x | x | x | x | x | | |

| ***ASSESSMENTS:*** | | | | | | | | | | | | | | | | |
|---|---|---|---|---|---|---|---|---|---|---|---|---|---|---|---|---|
| *Physical exam* [3] | x | | | | | | | | | | | | | | | |
| *Vital signs* [4] | x | | | | | | | | | | | | | | | |
| *CBC* [5] | x | | | | | | | | | | | | | | | |
| *LFTs* [5] | x | | | | | | | | | | | | | | | |
| *RFTs* [5] | x | | | | | | | | | | | | | | | |
| *Clinical evaluation* | x | | | | | | | | | | | | | | | |
| *12 lead ECG* | x | | | | | | | | | | | | | | | |
| *Chest X-ray* | x | | | | | | | | | | | | | | | |
| *Alleviation of symptoms* | x | | ← | | | | | | | | | | | | | → |
| *RT-PCR* [6] | | ← | | | | | | | | | | | | | | → |
| *Clinical Grading Status on 7-pointer scale* | x | | | | x | | x | | x | | x | | x | | | |
| *Fever* | x | x | x | x | x | x | x | x | x | x | x | x | x | x | | |
| *Cough* | x | x | x | x | x | x | x | x | x | x | x | x | x | x | | |

| | | | | | | | | | | | | | | | | |
|---|---|---|---|---|---|---|---|---|---|---|---|---|---|---|---|---|
| *Shortness of breath* | x | x | x | x | x | x | x | x | x | x | x | x | x | x | | |
| *Myalgia* | x | x | x | x | x | x | x | x | x | x | x | x | x | x | | |
| *Rate your emotional status* | x | x | x | x | x | x | x | x | x | x | x | x | x | x | | |
| *How sick do you feel* | x | x | x | x | x | x | x | x | x | x | x | x | x | x | | |
| *Oxygen saturation on room air* | x | x | x | x | x | x | x | x | x | x | x | x | x | x | | |
| *Oxygen Therapy* | x | x | x | x | x | x | x | x | x | x | x | x | x | x | | |
| *Adverse effects of drug administration* | x | x | x | x | x | x | x | x | x | x | x | x | x | x | | |
| *Mortality* | | | ← | | | | | | | | | | | | | → |

**Figure S3:** Schedule of enrollment, interventions, and assessments according to the Standard Protocol Items. [1] Demographics include Name (optional), Age, Gender, Contact Number (mandatory), Profession, Blood group. [2] Epidemiological information includes recent travel history and exposure to epidemic area or infected patients; medical history includes present illness and past medical history including co-morbidity and medications. [3] Physical examination includes height, weight, general outlook, head, eye, ear, nose, throat, neck and thyroid, skin, cardiovascular system, Anterior chest and lung, abdomen, neuro system, skeletal muscle system and lymphatic system. [4] Vital signs: body temperature, respiratory rate, pulse, blood pressure; record maximum temperature of the visiting day and use same measurement criteria throughout the study period. [5] Laboratory parameters be exempted by the investigators if the participants can provide qualified examination results within 48 hours prior to initiation of the study [6] Second PCR will be performed 48 to 72 hours after the patient becomes asymptomatic if it’s still positive another PCR will be conducted on day 14th

#### Additional Details on Statistical Analyses

Biostatistician using SAS software version 9.4 analysed all trial data. Mean ± S.D was used for quantitative data and f (%) was be used for categorical data. Frequency and percentages were measured for categorical data. Data supporting the necessary assumptions of time to event data, survival analysis/ Kaplan Meier test was applied. P-value < 0.05 was considered as significant.

| Outcome | Hypothesis | Outcome measure | Method of analysis |
|---|---|---|---|
| **PRIMARY OUTCOMES** | | | |
| 1. Time Taken for Alleviation of Symptoms | Earlier resolution of symptoms | Yes/No | Multivariant analysis, cox proportional hazards models |
| 2. Viral Load Clearance | Earlier RT-PCR negative | Positive/ negative | |
| 3. Clinical Grading Score (CGS) at Day 6 | Lower CGS score on seven pointer ordinal scale at day 6 | Grade from 1-7<br>Grade 1. Not hospitalized with resumption of normal daily life activities<br>Grade 2. Not hospitalized but unable to resume normal daily life activities<br>Grade 3. Hospitalized, but do not require supplemental oxygen therapy<br>Grade 4. Hospitalized but require low flow supplemental oxygen therapy<br>Grade 5. Hospitalized, requiring high flow supplemental oxygen<br>Grade 6. Hospitalized, requiring mechanical ventilation<br>Grade 7. Death | Mixed Ordinal Logistic Regression Model |

| SECONDARY OUTCOMES | | | |
|---|---|---|---|
| 1. Degree of fever at day 4 | Lower degree of fever at day 4 | Yes/No | Mixed Ordinal Logistic Regression Model |
| 2. CRP at Day 6 | Lower CRP levels at day 6 | Yes/No | Linear regression model |
| 3. Severity of symptoms at day 8 | Lower degree of severity at day 8 | Severity of symptoms is classified as mild, moderate or severe. | Mixed Ordinal Logistic Regression Model |
| 4. Clinical Grading Score (CGS) at Day 10 | Lower CGS score on seven pointer ordinal scale at day 10 | Grade from 1-7<br>Grade 1. Not hospitalized with resumption of normal daily life activities<br>Grade 2. Not hospitalized but unable to resume normal daily life activities<br>Grade 3. Hospitalized, but do not require supplemental oxygen therapy<br>Grade 4. Hospitalized but require low flow supplemental oxygen therapy<br>Grade 5. Hospitalized, requiring high flow supplemental oxygen<br>Grade 6. Hospitalized, requiring mechanical ventilation<br>Grade 7. Death | Mixed Ordinal Logistic Regression Mode |
| 5. 30 Day Mortality | Lower mortality | Yes/No | Fisher’s Exact test |

| ADDITIONAL OUTCOMES | | | |
|---|---|---|---|
| **Median time to clinical improvement of severity of symptoms** | | | |
| Improvement of one category on ordinal scale | Earlier improvement in severity of symptoms | 1. None<br>2. Mild<br>3. Moderate<br>4. Severe | Univariate Log-rank test |
| Improvement of two category on ordinal scale | | | |
| Achievement of normal status on ordinal scale | | | |
| **Median time to clinical improvement of degree of fever** | | | |
| Improvement of one category on ordinal scale | Earlier improvement in degree of fever | 1. None: <99 °F<br>2. Mild: 99-100 °F<br>3. Moderate: 100-101.9 °F<br>4. Severe: ≥ 102°F | Univariate Log-rank test |
| Improvement of two category on ordinal scale | | | |
| Achievement of normal status on ordinal scale | | | |
| **Median time to clinical improvement of cough** | | | |
| Improvement of one category on ordinal scale | Earlier improvement in degree of cough | 1. None: No cough.<br>2. Mild: Occasional, transient cough<br>3. Moderate: Frequent cough, slightly influencing daytime activities<br>5. Severe: Frequent cough, significantly influencing daytime activities | Univariate Log-rank test |
| Improvement of two category on ordinal scale | | | |
| Achievement of normal status on ordinal scale | | | |

| | | | |
|---|---|---|---|
| **Median time to clinical improvement of shortness of breath** | | | |
| Improvement of one category on ordinal scale | Earlier improvement in degree of shortness of breath | Grade1= Not troubled by breathlessness except on strenuous exercise<br>Grade 2= Short of breath when hurrying on the level or walking up a slight hill<br>Grade 3= Walks slower than most people on the level, stops after a mile or so, or stop after 15 minutes walking at own pace<br>Grade 4= Stops for breath after walking about 100yds or a few minutes on level ground.<br>Grade 5= Too breathless to leave the house, or breathless when undressing | Univariate Log-rank test |
| Improvement of two category on ordinal scale | | | |
| Achievement of normal status on ordinal scale | | | |
| **Median time to clinical improvement of myalgia** | | | |
| Improvement of one category on ordinal scale | Earlier improvement in degree of myalgia | 1. None<br>2. Mild<br>3. Moderate<br>4. Severe | Univariate Log-rank test |
| Improvement of two category on ordinal scale | | | |
| Achievement of normal status on ordinal scale | | | |
| **Median time to clinical improvement of "how sick do you feel"** | | | |
| Improvement of one category on ordinal scale | Earlier improvement in status "How sick do you feel" | 1. None<br>2. Mild<br>3. Moderate<br>4. Severe | Univariate Log-rank test |
| Improvement of two category on ordinal scale | | | |
| Achievement of normal status on ordinal scale | | | |

**Figure S4: Statistical Analysis.** Two tailed P-value < 0.05 with 95% confidence interval was considered as significant.

### SUPPLEMENTARY TABLES

| Table S1. **Characteristics of the Patients at Baseline (Intention-to-Treat Population)** * | | | |
|---|---|---|---|
| **Baseline Demographics** | **Total (n=313)** | **Standard Care Alone (n=156)** | **Honey-*Nigella Sativa* (n=157)** |
| **Age (Year) — No. (%)** | | | |
| ≤40 | 156 (49.84) | 80 (25.56) | 76 (24.28) |
| 40-59 | 93 (29.71) | 45 (14.38) | 48 (15.34) |
| 60-79 | 52 (16.61) | 26 (8.31) | 26 (8.31) |
| ≥80 | 12 (3.83) | 5 (1.6) | 7 (2.24) |
| **Sex — No. (%)** | | | |
| Male | 178 (56.87) | 88 (56.41) | 90 (57.32) |
| Female | 135 (43.13) | 68 (43.59) | 67 (42.68) |
| **Profession — No. (%)** | | | |
| Health care¶ | 71 (22.68) | 38 (24.36) | 33 (21.02) |
| Non-Health care | 242 (77.32) | 118 (75.64) | 124 (78.98) |
| **Co-Morbidities — No. (%)** | | | |
| Hypertension | 99 (31.63) | 51 (16.29) | 48 (15.34) |
| Diabetes Mellitus | 115 (36.74) | 60 (19.17) | 55 (17.57) |
| **Onset of symptoms before admission — No. (%)** | | | |
| 48 hours | 88 (38.1) | 49 (41.53) | 39 (34.51) |
| 72 hours | 143 (61.9) | 69 (58.47) | 74 (65.49) |
| 96 hours | 82 (36.44) | 38 (35.51) | 44 (37.29) |
| **Severity of Symptoms — No. (%)** | | | |
| Moderate | 210 (67.09) | 103 (66.03) | 107 (68.15) |
| Severe | 103 (32.91) | 53 (33.97) | 50 (31.85) |

| | | | |
|---|---|---|---|
| **ARDS —No. (%)** | 57 (17.38) | 28 (17.95) | 29 (16.86) |
| **Chest X-Ray — No. (%)** | | | |
| Normal | 217 (66.16) | 101 (30.79) | 116 (35.37) |
| Pneumonic Patch | 12 (3.66) | 8 (2.44) | 4 (1.22) |
| Unilateral Infiltrates | 40 (12.2) | 19 (5.79) | 21 (6.4) |
| Bilateral Infiltrates | 59 (17.99) | 28 (8.54) | 31 (9.45) |
| **ECG — No. (%)** | | | |
| Normal | 230(73.48) | 119(38.02) | 111(35.46) |
| LBBB | 62(19.81) | 28(8.95) | 34(10.86) |
| Other | 21(6.71) | 9(2.88) | 12(3.83) |
| **Clinical Status Grading at Day 0 — No. (%)** | | | |
| 2- Not hospitalized with unable to resume normal activities — No. (%) | 139 (44.41) | 68 (43.59) | 71 (45.22) |
| 3- Hospitalized, not requiring supplemental oxygen — No. (%) | 71 (22.68) | 35 (22.44) | 36 (22.93) |
| 4- Hospitalized, requiring low flow supplemental oxygen — No. (%) | 44 (14.06) | 23 (14.74) | 21 (13.38) |
| 5- Hospitalized, requiring high flow supplemental oxygen — No. (%) | 59 (18.85) | 30 (19.23) | 29 (18.47) |
| **Hospital — No. (%)** | | | |
| Shaikh Zayed Hospital | 78 (25.66) | 39 (25.83) | 39 (25.49) |
| Services Institute of Medical Sciences | 91 (29.93) | 48 (31.79) | 43 (28.1) |
| Doctors Lounge | 52 (17.11) | 27 (17.88) | 25 (16.34) |
| Ali Clinic [III] | 83 (27.3) | 37 (24.5) | 46 (30.07) |

| | | | |
|---|---|---|---|
| **Frequency of Symptoms — No. (%)** | | | |
| Fever | 303 (96.81) | 152 (48.56) | 151 (48.24) |
| SOB | 106 (33.87) | 56 (17.89) | 50 (15.97) |
| Cough | 192 (61.34) | 90 (28.75) | 102 (32.59) |
| Myalgia | 169 (53.99) | 89 (28.43) | 80 (25.56) |
| Sputum | 23(7.35) | 13(4.15) | 10(3.19) |
| Anorexia | 63(20.13) | 31(9.9) | 32(10.22) |
| Fatigue | 47(15.02) | 27(8.63) | 20(6.39) |
| Loss of smell | 76(24.28) | 36(11.5) | 40(12.78) |
| Change in taste | 77(24.6) | 39(12.46) | 38(12.14) |
| **Treatment Options — No. (%) ¥** | | | |
| Panadol | 297 (94.89) | 147 (46.96) | 150 (47.92) |
| Azithromycin | 231 (73.8) | 120 (38.34) | 111 (35.46) |
| Montelukast | 106 (33.87) | 56 (17.89) | 50 (15.97) |
| Supplemental Oxygen | 105 (33.55) | 55 (17.57) | 50 (15.97) |
| Low Molecular Weight Heparin | 72 (23) | 38 (12.14) | 34 (10.86) |
| Hydrocortisone | 83 (26.52) | 45 (14.38) | 38 (12.14) |
| Multivitamins | 147 (46.96) | 73 (23.32) | 74 (23.64) |
| Tazobactam + Piperacillin | 73 (23.32) | 42 (13.42) | 31 (9.9) |
| Ivermectin | 114 (36.42) | 60 (19.17) | 54 (17.25) |
| Meropenem | 62 (19.81) | 35 (11.18) | 27 (8.63) |
| Ibuprofen | 101(32.27) | 57(18.21) | 44(14.06) |

| | | | |
|---|---|---|---|
| **Blood Group— No. (%)** | | | |
| A+ | 92(29.39) | 45(14.38) | 47(15.02) |
| A- | 7(2.24) | 2(0.64) | 5(1.6) |
| B+ | 96(30.67) | 54(17.25) | 42(13.42) |
| B- | 19(6.07) | 12(3.83) | 7(2.24) |
| AB+ | 50(15.97) | 24(7.67) | 26(8.31) |
| AB- | 11(3.51) | 2(0.64) | 9(2.88) |
| O+ | 28(8.95) | 11(3.51) | 17(5.43) |
| O- | 10(3.19) | 6(1.92) | 4(1.28) |
| **Vitals— No. (%)** | | | |
| Systolic Blood Pressure, median (IQR) — mm of Hg | 120(110-140) | 120(110-140) | 120(110-140) |
| Diastolic Blood Pressure, median (IQR) — mm of Hg | 80(70-85) | 80(70-85) | 80(70-90) |
| Temperature, median (IQR) — °F | 101(100-102) | 101(100-102) | 101(100-102) |
| ≤100°F — No. (%) | 89(28.43) | 46(14.7) | 43(13.74) |
| 100°F-102°F — No. (%) | 100(31.95) | 46(14.7) | 54(17.25) |
| ≥102°F —No. (%) | 124(39.62) | 64(20.45) | 60(19.17) |
| Respiratory Rate, median (IQR) — breaths per minute | 19(18-25) | 20.50(18-26) | 18(18-24) |
| Heart Rate, median (IQR) — beats per minute | 87(78-105) | 88(78-106) | 86(78-100) |

| **Laboratory Parameters** | | | |
|---|---|---|---|
| Hemoglobin, median (IQR) — mg/dL | 13.30(12.60-14.20) | 13.30(12.60-14.20) | 13.30(12.40-14.20) |
| Total Leukocyte Count, median (IQR) — mg/dL | 6.40(5.60-7.70) | 6.40(5.60-8.40) | 6.30(5.50-7.40) |
| Neutrophils, median (IQR) — mg/dL | 6.0(4.70-6.90) | 6.05(4.70-7.10) | 5.90(4.70-6.90) |
| Lymphocyte, median (IQR) — mg/dL | 1.90(1.60-2.30) | 1.90(1.60-2.40) | 1.90(1.60-2.30) |
| Platelets, median (IQR) — mg/dL | 229(189-255) | 229(189-264) | 223(188.0-253.0) |
| ALT, median (IQR) — U/L | 43(39-47) | 43(38-47) | 44(40.0-48.0) |
| AST, median (IQR) — U/L | 33(29.0-38.0) | 32(29-28) | 33(29.0-38.0) |
| Creatinine, median (IQR) — mg/dL | 1(1.0-1.1) | 1(0.9-1.20) | 1(1.0-1.1) |
| ≤ 1.3mg/dL — No. (%) | 295(147) | 148(94.87) | 147(93.63) |
| >1.3mg/dL — No. (%) | 18(10) | 8(5.13) | 10(6.37) |
| CRP, median (IQR) — mg/L | 16(11-21) | 17(12-21) | 14(10.0-21.0) |
| O2 Saturation at room air, median (IQR) — (%) | 96(87-96) | 96(86-96) | 96(87-97) |

*Data are presented as no. (%) unless indicated otherwise. The Intention-to-Treat Population included all the patients who had undergone randomization. Information on co-morbidities, profession, age and gender was obtained from medical records. ECMO denotes extracorporeal membrane oxygenation, CRP C-Reactive protein, AST Aspartate transaminase ALT Alanine transaminase, ECG electrocardiography, ARDS acute respiratory distress syndrome and IQR interquartile range.

¥ These medications were part of standard care therapy as per Clinical Management Guidelines for COVID-19 by Ministry of National Health Services, Pakistan.

¶ Medical doctors, nurses and paramedics

**Table S2**. Distribution of severity of cough over time*

| | | | Number of Days | | | | | | | | | | | | |
|---|---|---|---|---|---|---|---|---|---|---|---|---|---|---|---|
| | | | **1** | **2** | **3** | **4** | **5** | **6** | **7** | **8** | **9** | **10** | **11** | **12** | **13** |
| *Standard Care Alone* | **Moderate** | **None**[€] | 45 (43.69) | 44 (42.72) | 48 (46.6) | 43 (41.75) | 47 (45.63) | 58 (56.31) | 68 (66.02) | 83 (80.58) | 91 (88.35) | 92 (89.32) | 97 (94.17) | 99 (96.12) | 99 (97.06) |
| | | **Mild** [db] | 9 (8.74) | 11 (10.68) | 18 (17.48) | 31 (30.1) | 31 (30.1) | 30 (29.13) | 25 (24.27) | 13 (12.62) | 8 (7.77) | 7 (6.8) | 3 (2.91) | 2 (1.94) | 1 (0.98) |
| | | **Moderate**[φ] | 43 (41.75) | 42 (40.78) | 33 (32.04) | 29 (28.16) | 24 (23.3) | 15 (14.56) | 10 (9.71) | 7 (6.8) | 3 (2.91) | 4 (3.88) | 3 (2.91) | 2 (1.94) | 1 (0.98) |
| | | **Severe** [Σ] | 6 (5.83) | 6 (5.83) | 4 (3.88) | 0 (0) | 1 (0.97) | 0 (0) | 0 (0) | 0 (0) | 1 (0.97) | 0 (0) | 0 (0) | 0 (0) | 1 (0.98) |
| | **Severe** | **None**[€] | 10 (18.87) | 8 (15.09) | 7 (13.21) | 6 (11.32) | 14 (26.42) | 16 (30.19) | 18 (35.29) | 26 (50.98) | 26 (53.06) | 29 (59.18) | 29 (61.7) | 34 (73.91) | 36 (78.26) |
| | | **Mild** [db] | 0 (0) | 1 (1.89) | 2 (3.77) | 9 (16.98) | 8 (15.09) | 17 (32.08) | 14 (27.45) | 8 (15.69) | 14 (28.57) | 7 (14.29) | 4 (8.51) | 4 (8.7) | 5 (10.87) |
| | | **Moderate**[φ] | 15 (28.3) | 24 (45.28) | 26 (49.06) | 22 (41.51) | 23 (43.4) | 13 (24.53) | 16 (31.37) | 15 (29.41) | 9 (18.37) | 11 (22.45) | 10 (21.28) | 5 (10.87) | 5 (10.87) |
| | | **Severe** [Σ] | 28 (52.83) | 20 (37.74) | 18 (33.96) | 16 (30.19) | 8 (15.09) | 7 (13.21) | 3 (5.88) | 2 (3.92) | 0 (0) | 2 (4.08) | 4 (8.51) | 3 (6.52) | 0 (0) |
| *Honey-Nigella Sativa* | **Moderate** | **None**[€] | 42 (39.25) | 41 (38.32) | 51 (47.66) | 66 (61.68) | 75 (70.09) | 105 (98.13) | 105 (98.13) | 105 (98.13) | 107 (100) | 106 (99.07) | 104 (97.2) | 107 (100) | 107 (100) |
| | | **Mild** [db] | 5 (4.67) | 11 (10.28) | 30 (28.04) | 34 (31.78) | 31 (28.97) | 2 (1.87) | 2 (1.87) | 2 (1.87) | 0 (0) | 1 (0.93) | 3 (2.8) | 0 (0) | 0 (0) |
| | | **Moderate**[φ] | 53 (49.53) | 50 (46.73) | 25 (23.36) | 7 (6.54) | 1 (0.93) | 0 (0) | 0 (0) | 0 (0) | 0 (0) | 0 (0) | 0 (0) | 0 (0) | 0 (0) |
| | | **Severe** [Σ] | 7 (6.54) | 5 (4.67) | 1 (0.93) | 0 (0) | 0 (0) | 0 (0) | 0 (0) | 0 (0) | 0 (0) | 0 (0) | 0 (0) | 0 (0) | 0 (0) |
| | **Severe** | **None**[€] | 10 (20) | 9 (18) | 14 (28) | 13 (26) | 22 (44) | 30 (60) | 36 (72) | 38 (76) | 39 (78) | 44 (88) | 40 (80) | 46 (92) | 45 (90) |
| | | **Mild** [db] | 7 (14) | 4 (8) | 3 (6) | 15 (30) | 14 (28) | 8 (16) | 7 (14) | 5 (10) | 6 (12) | 2 (4) | 4 (8) | 0 (0) | 0 (0) |
| | | **Moderate**[φ] | 10 (20) | 20 (40) | 26 (52) | 19 (38) | 10 (20) | 10 (20) | 4 (8) | 7 (14) | 4 (8) | 4 (8) | 5 (10) | 3 (6) | 5 (10) |
| | | **Severe** [Σ] | 23 (46) | 17 (34) | 7 (14) | 3 (6) | 4 (8) | 2 (4) | 3 (6) | 0 (0) | 1 (2) | 0 (0) | 1 (2) | 1 (2) | 0 (0) |

* Data are presented as no. (%) unless indicated otherwise. Cough is classified as mild, moderate and severe, **€**No cough, **db** occasional/transient cough, **φ** frequent cough, slightly influencing daytime activities, **Σ** frequent cough, significantly influencing daytime activities

**Table S3**. Distribution of Severity of Symptoms over time*

| | | | Number of Days | | | | | | | | | | | | |
|---|---|---|---|---|---|---|---|---|---|---|---|---|---|---|---|
| | | | **1** | **2** | **3** | **4** | **5** | **6** | **7** | **8** | **9** | **10** | **11** | **12** | **13** |
| ***Standard Care Alone*** | **Moderate** | **None**[€] | 0 (0) | 0 (0) | 3 (2.91) | 5 (4.85) | 9 (8.74) | 29 (28.16) | 39 (37.86) | 58 (56.31) | 67 (65.05) | 77 (74.76) | 97 (94.17) | 97 (94.17) | 99 (96.12) |
| | | **Mild** [ȸ] | 2 (1.94) | 3 (2.91) | 9 (8.74) | 23 (22.33) | 40 (38.83) | 36 (34.95) | 32 (31.07) | 18 (17.48) | 24 (23.3) | 22 (21.36) | 2 (1.94) | 2 (1.94) | 0 (0) |
| | | **Moderate**[φ] | 91 (88.35) | 91 (88.35) | 81 (78.64) | 67 (65.05) | 45 (43.69) | 29 (28.16) | 24 (23.3) | 21 (20.39) | 8 (7.77) | 0 (0) | 0 (0) | 0 (0) | 0 (0) |
| | | **Severe** [Σ] | 10 (9.71) | 9 (8.74) | 10 (9.71) | 8 (7.77) | 9 (8.74) | 9 (8.74) | 8 (7.77) | 6 (5.83) | 4 (3.88) | 4 (3.88) | 4 (3.88) | 4 (3.88) | 4 (3.88) |
| | **Severe** | **None**[€] | 0 (0) | 0 (0) | 0 (0) | 1 (1.89) | 0 (0) | 5 (9.62) | 6 (11.76) | 10 (19.61) | 14 (28) | 19 (38) | 24 (50) | 24 (50) | 27 (56.25) |
| | | **Mild** [ȸ] | 0 (0) | 0 (0) | 1 (1.89) | 2 (3.77) | 4 (7.69) | 4 (7.69) | 13 (25.49) | 15 (29.41) | 9 (18) | 5 (10) | 0 (0) | 4 (8.33) | 3 (6.25) |
| | | **Moderate**[φ] | 0 (0) | 2 (3.77) | 7 (13.21) | 15 (28.3) | 24 (46.15) | 16 (30.77) | 8 (15.69) | 4 (7.84) | 12 (24) | 9 (18) | 13 (27.08) | 7 (14.58) | 9 (18.75) |
| | | **Severe** [Σ] | 53 (100) | 51 (96.23) | 45 (84.91) | 35 (66.04) | 24 (46.15) | 27 (51.92) | 24 (47.06) | 22 (43.14) | 15 (30) | 17 (34) | 11 (22.92) | 13 (27.08) | 9 (18.75) |
| ***Honey-Nigella Sativa*** | **Moderate** | **None**[€] | 0 (0) | 0 (0) | 4 (3.74) | 43 (40.19) | 60 (56.07) | 101 (94.39) | 104 (97.2) | 105 (98.13) | 107 (100) | 106 (99.07) | 106 (99.07) | 107 (100) | 106 (99.07) |
| | | **Mild** [ȸ] | 2 (1.87) | 9 (8.41) | 52 (48.6) | 45 (42.06) | 41 (38.32) | 5 (4.67) | 2 (1.87) | 2 (1.87) | 0 (0) | 1 (0.93) | 1 (0.93) | 0 (0) | 1 (0.93) |
| | | **Moderate**[φ] | 98 (91.59) | 93 (86.92) | 50 (46.73) | 18 (16.82) | 5 (4.67) | 0 (0) | 0 (0) | 0 (0) | 0 (0) | 0 (0) | 0 (0) | 0 (0) | 0 (0) |
| | | **Severe** [Σ] | 7 (6.54) | 5 (4.67) | 1 (0.93) | 1 (0.93) | 1 (0.93) | 1 (0.93) | 1 (0.93) | 0 (0) | 0 (0) | 0 (0) | 0 (0) | 0 (0) | 0 (0) |
| | **Severe** | **None**[€] | 0 (0) | 0 (0) | 0 (0) | 1 (2) | 6 (12) | 24 (48) | 28 (56) | 35 (70) | 39 (78) | 41 (82) | 41 (82) | 41 (82) | 41 (82) |
| | | **Mild** [ȸ] | 0 (0) | 0 (0) | 1 (2) | 13 (26) | 21 (42) | 12 (24) | 13 (26) | 7 (14) | 2 (4) | 0 (0) | 0 (0) | 0 (0) | 0 (0) |
| | | **Moderate**[φ] | 0 (0) | 6 (12) | 24 (48) | 24 (48) | 16 (32) | 9 (18) | 2 (4) | 2 (4) | 6 (12) | 3 (6) | 5 (10) | 5 (10) | 5 (10) |
| | | **Severe** [Σ] | 50 (100) | 44 (88) | 25 (50) | 12 (24) | 7 (14) | 5 (10) | 7 (14) | 6 (12) | 3 (6) | 6 (12) | 4 (8) | 4 (8) | 4 (8) |

*Data are presented as no. (%) unless indicated otherwise. Severity of symptoms is classified as mild, moderate and severe. **€** No symptoms, **ȸ** symptoms of an upper respiratory tract viral infection i.e. low grade fever, dry cough, sore throat, nasal congestion, malaise but no radiological evidence of pneumonia **φ** Moderate disease includes patients with Hypoxia (Oxygen saturation <94% but >90%) or chest X-ray with infiltrates involving <50% of the lung fields or fever, cough, sputum production, and other respiratory tract or non-specific symptoms along with manifestation of pneumonia on imaging but no signs of severe pneumonia, **Σ** Fever is associated with severe dyspnoea, respiratory distress, tachypnea (> 30 breaths/min), and hypoxia ($SpO_2$ < 94% on room air)" or a $PaO_2$ to $FiO_2$ ratio of 300 or lower.

**Table S4**. Distribution of fever over time*

| | | | Number of Days | | | | | | | | | | | | |
|---|---|---|---|---|---|---|---|---|---|---|---|---|---|---|---|
| | | | **1** | **2** | **3** | **4** | **5** | **6** | **7** | **8** | **9** | **10** | **11** | **12** | **13** |
| ***Standard Care Alone*** | **Moderate** | **None€** | 1 (0.97) | 1 (0.97) | 1 (0.97) | 4 (3.88) | 14 (13.59) | 37 (35.92) | 46 (44.66) | 63 (61.17) | 72 (69.9) | 87 (84.47) | 96 (93.2) | 101 (98.06) | 102 (99.03) |
| | | **Mild ȸ** | 0 (0) | 7 (6.8) | 14 (13.59) | 30 (29.13) | 47 (45.63) | 29 (28.16) | 36 (34.95) | 24 (23.3) | 23 (22.33) | 10 (9.71) | 5 (4.85) | 0 (0) | 0 (0) |
| | | **Moderateȹ** | 90 (87.38) | 86 (83.5) | 81 (78.64) | 60 (58.25) | 36 (34.95) | 33 (32.04) | 19 (18.45) | 15 (14.56) | 8 (7.77) | 6 (5.83) | 2 (1.94) | 2 (1.94) | 1 (0.97) |
| | | **Severe Σ** | 12 (11.65) | 9 (8.74) | 7 (6.8) | 9 (8.74) | 6 (5.83) | 4 (3.88) | 2 (1.94) | 1 (0.97) | 0 (0) | 0 (0) | 0 (0) | 0 (0) | 0 (0) |
| | **Severe** | **None€** | 3 (5.66) | 3 (5.66) | 3 (5.66) | 2 (3.77) | 6 (11.54) | 11 (21.15) | 16 (31.37) | 19 (37.25) | 21 (42.86) | 26 (53.06) | 30 (63.83) | 32 (69.57) | 34 (73.91) |
| | | **Mild ȸ** | 0 (0) | 0 (0) | 1 (1.89) | 12 (22.64) | 9 (17.31) | 7 (13.46) | 10 (19.61) | 11 (21.57) | 12 (24.49) | 6 (12.24) | 2 (4.26) | 5 (10.87) | 6 (13.04) |
| | | **Moderateȹ** | 6 (11.32) | 11 (20.75) | 28 (52.83) | 23 (43.4) | 23 (44.23) | 25 (48.08) | 19 (37.25) | 14 (27.45) | 13 (26.53) | 12 (24.49) | 12 (25.53) | 8 (17.39) | 6 (13.04) |
| | | **Severe Σ** | 44 (83.02) | 39 (73.58) | 21 (39.62) | 16 (30.19) | 14 (26.92) | 9 (17.31) | 6 (11.76) | 7 (13.73) | 3 (6.12) | 5 (10.2) | 3 (6.38) | 1 (2.17) | 0 (0) |
| ***Honey-Nigella Sativa*** | **Moderate** | **None€** | 4 (3.74) | 4 (3.74) | 9 (8.41) | 63 (58.88) | 80 (74.77) | 100 (93.46) | 101 (94.39) | 107 (100) | 107 (100) | 107 (100) | 107 (100) | 107 (100) | 107 (100) |
| | | **Mild ȸ** | 0 (0) | 19 (17.76) | 67 (62.62) | 31 (28.97) | 21 (19.63) | 6 (5.61) | 5 (4.67) | 0 (0) | 0 (0) | 0 (0) | 0 (0) | 0 (0) | 0 (0) |
| | | **Moderateȹ** | 96 (89.72) | 80 (74.77) | 30 (28.04) | 12 (11.21) | 6 (5.61) | 1 (0.93) | 1 (0.93) | 0 (0) | 0 (0) | 0 (0) | 0 (0) | 0 (0) | 0 (0) |
| | | **Severe Σ** | 7 (6.54) | 4 (3.74) | 1 (0.93) | 1 (0.93) | 0 (0) | 0 (0) | 0 (0) | 0 (0) | 0 (0) | 0 (0) | 0 (0) | 0 (0) | 0 (0) |
| | **Severe** | **None€** | 2 (4) | 2 (4) | 2 (4) | 11 (22) | 17 (34) | 31 (62) | 32 (64) | 35 (70) | 40 (80) | 44 (88) | 47 (94) | 47 (94) | 49 (98) |
| | | **Mild ȸ** | 0 (0) | 0 (0) | 7 (14) | 13 (26) | 20 (40) | 8 (16) | 11 (22) | 7 (14) | 2 (4) | 0 (0) | 0 (0) | 0 (0) | 0 (0) |
| | | **Moderateȹ** | 3 (6) | 24 (48) | 28 (56) | 24 (48) | 10 (20) | 8 (16) | 6 (12) | 6 (12) | 6 (12) | 4 (8) | 2 (4) | 2 (4) | 1 (2) |
| | | **Severe Σ** | 45 (90) | 24 (48) | 13 (26) | 2 (4) | 3 (6) | 3 (6) | 1 (2) | 2 (4) | 2 (4) | 2 (4) | 1 (2) | 1 (2) | 0 (0) |

* Data are presented as no. (%) unless indicated otherwise and fever is classified as mild, moderate and severe**,** **€**98-99 °F, **ȸ** <100 °F, **ȹ**100-101.9 °F, **Σ** ≥ 102°F

**Table S5**. Distribution of Myalgia over time*

| | | | Number of Days | | | | | | | | | | | | |
|---|---|---|---|---|---|---|---|---|---|---|---|---|---|---|---|
| | | | **1** | **2** | **3** | **4** | **5** | **6** | **7** | **8** | **9** | **10** | **11** | **12** | **13** |
| ***Standard Care Alone*** | **Moderate** | **None€** | 40 (38.83) | 42 (40.78) | 38 (36.89) | 47 (45.63) | 47 (45.63) | 65 (63.11) | 73 (70.87) | 76 (73.79) | 84 (81.55) | 99 (96.12) | 96 (93.2) | 101 (98.06) | 101 (98.06) |
| | | **Mild ȸ** | 9 (8.74) | 9 (8.74) | 25 (24.27) | 27 (26.21) | 29 (28.16) | 19 (18.45) | 23 (22.33) | 20 (19.42) | 14 (13.59) | 0 (0) | 5 (4.85) | 0 (0) | 0 (0) |
| | | **Moderateφ** | 49 (47.57) | 49 (47.57) | 36 (34.95) | 26 (25.24) | 27 (26.21) | 19 (18.45) | 7 (6.8) | 6 (5.83) | 4 (3.88) | 3 (2.91) | 1 (0.97) | 1 (0.97) | 2 (1.94) |
| | | **Severe Σ** | 5 (4.85) | 3 (2.91) | 4 (3.88) | 3 (2.91) | 0 (0) | 0 (0) | 0 (0) | 1 (0.97) | 1 (0.97) | 1 (0.97) | 1 (0.97) | 1 (0.97) | 0 (0) |
| | **Severe** | **None€** | 19 (35.85) | 19 (35.85) | 17 (32.08) | 18 (33.96) | 21 (40.38) | 19 (36.54) | 24 (47.06) | 25 (49.02) | 31 (63.27) | 32 (65.31) | 37 (78.72) | 42 (91.3) | 42 (91.3) |
| | | **Mild ȸ** | 0 (0) | 0 (0) | 8 (15.09) | 9 (16.98) | 6 (11.54) | 9 (17.31) | 11 (21.57) | 7 (13.73) | 8 (16.33) | 7 (14.29) | 6 (12.77) | 3 (6.52) | 3 (6.52) |
| | | **Moderateφ** | 24 (45.28) | 20 (37.74) | 19 (35.85) | 23 (43.4) | 19 (36.54) | 17 (32.69) | 10 (19.61) | 18 (35.29) | 9 (18.37) | 10 (20.41) | 4 (8.51) | 1 (2.17) | 1 (2.17) |
| | | **Severe Σ** | 10 (18.87) | 14 (26.42) | 9 (16.98) | 3 (5.66) | 6 (11.54) | 7 (13.46) | 6 (11.76) | 1 (1.96) | 1 (2.04) | 0 (0) | 0 (0) | 0 (0) | 0 (0) |
| ***Honey-Nigella Sativa*** | **Moderate** | **None€** | 58 (54.21) | 54 (50.47) | 67 (62.62) | 78 (72.9) | 95 (88.79) | 104 (97.2) | 107 (100) | 107 (100) | 107 (100) | 104 (97.2) | 107 (100) | 107 (100) | 105 (98.13) |
| | | **Mild ȸ** | 9 (8.41) | 28 (26.17) | 31 (28.97) | 25 (23.36) | 11 (10.28) | 3 (2.8) | 0 (0) | 0 (0) | 0 (0) | 3 (2.8) | 0 (0) | 0 (0) | 2 (1.87) |
| | | **Moderateφ** | 38 (35.51) | 23 (21.5) | 8 (7.48) | 3 (2.8) | 1 (0.93) | 0 (0) | 0 (0) | 0 (0) | 0 (0) | 0 (0) | 0 (0) | 0 (0) | 0 (0) |
| | | **Severe Σ** | 2 (1.87) | 2 (1.87) | 1 (0.93) | 1 (0.93) | 0 (0) | 0 (0) | 0 (0) | 0 (0) | 0 (0) | 0 (0) | 0 (0) | 0 (0) | 0 (0) |
| | **Severe** | **None€** | 19 (38) | 18 (36) | 19 (38) | 27 (54) | 36 (72) | 40 (80) | 44 (88) | 45 (90) | 46 (92) | 45 (90) | 47 (94) | 48 (96) | 47 (94) |
| | | **Mild ȸ** | 1 (2) | 3 (6) | 9 (18) | 11 (22) | 6 (12) | 3 (6) | 0 (0) | 1 (2) | 1 (2) | 2 (4) | 1 (2) | 1 (2) | 0 (0) |
| | | **Moderateφ** | 18 (36) | 18 (36) | 19 (38) | 11 (22) | 5 (10) | 7 (14) | 4 (8) | 3 (6) | 2 (4) | 3 (6) | 2 (4) | 1 (2) | 3 (6) |
| | | **Severe Σ** | 12 (24) | 11 (22) | 3 (6) | 1 (2) | 3 (6) | 0 (0) | 2 (4) | 1 (2) | 1 (2) | 0 (0) | 0 (0) | 0 (0) | 0 (0) |

* Data are presented as no. (%) unless indicated otherwise. Myalgia is classified as mild, moderate and severe on subjective basis. Patients have been provided with a scale of 1-10, with 1 being the lowest and 10 being the highest value.

**€**No Myalgia **ȸ** 1-3 **φ** 4-6 **Σ** >6-10

**Table S6**. Distribution of Shortness of Breath over time*

| | | | Number of Days | | | | | | | | | | | | |
|---|---|---|---|---|---|---|---|---|---|---|---|---|---|---|---|
| | | | **1** | **2** | **3** | **4** | **5** | **6** | **7** | **8** | **9** | **10** | **11** | **12** | **13** |
| *Standard Care Alone* | **Moderate** | **None€** | 99 (96.12) | 102 (99.03) | 99 (96.12) | 95 (92.23) | 93 (90.29) | 91 (88.35) | 94 (91.26) | 94 (91.26) | 96 (93.2) | 94 (91.26) | 96 (93.2) | 99 (96.12) | 99 (96.12) |
| | | **Mild ȸ** | 2 (1.94) | 1 (0.97) | 1 (0.97) | 3 (2.91) | 4 (3.88) | 6 (5.83) | 3 (2.91) | 3 (2.91) | 3 (2.91) | 3 (2.91) | 4 (3.88) | 0 (0) | 0 (0) |
| | | **Moderateφ** | 2 (1.94) | 0 (0) | 3 (2.91) | 1 (0.97) | 2 (1.94) | 1 (0.97) | 1 (0.97) | 0 (0) | 0 (0) | 3 (2.91) | 0 (0) | 0 (0) | 0 (0) |
| | | **Severe Σ** | 0 (0) | 0 (0) | 0 (0) | 4 (3.88) | 4 (3.88) | 5 (4.85) | 5 (4.85) | 5 (4.85) | 3 (2.91) | 2 (1.94) | 2 (1.94) | 3 (2.91) | 3 (2.91) |
| | | **Very Severe¥** | 0 (0) | 0 (0) | 0 (0) | 0 (0) | 0 (0) | 0 (0) | 0 (0) | 1 (0.97) | 1 (0.97) | 1 (0.97) | 1 (0.97) | 1 (0.97) | 1 (0.97) |
| | **Severe** | **None€** | 0 (0) | 1 (1.89) | 1 (1.89) | 7 (14) | 7 (13.46) | 12 (26.09) | 13 (25.49) | 22 (44.9) | 24 (50) | 23 (46.94) | 24 (53.33) | 25 (56.82) | 28 (63.64) |
| | | **Mild ȸ** | 3 (5.66) | 3 (5.66) | 8 (15.09) | 7 (14) | 6 (11.54) | 4 (8.7) | 9 (17.65) | 3 (6.12) | 0 (0) | 1 (2.04) | 0 (0) | 2 (4.55) | 3 (6.82) |
| | | **Moderateφ** | 18 (33.96) | 16 (30.19) | 12 (22.64) | 10 (20) | 17 (32.69) | 16 (34.78) | 8 (15.69) | 4 (8.16) | 5 (10.42) | 5 (10.2) | 6 (13.33) | 7 (15.91) | 4 (9.09) |
| | | **Severe Σ** | 17 (32.08) | 18 (33.96) | 17 (32.08) | 12 (24) | 6 (11.54) | 3 (6.52) | 6 (11.76) | 8 (16.33) | 7 (14.58) | 9 (18.37) | 10 (22.22) | 6 (13.64) | 5 (11.36) |
| | | **Very Severe¥** | 15 (28.3) | 15 (28.3) | 15 (28.3) | 14 (28) | 16 (30.77) | 11 (23.91) | 15 (29.41) | 12 (24.49) | 12 (25) | 11 (22.45) | 5 (11.11) | 4 (9.09) | 4 (9.09) |
| *Honey-Nigella Sativa* | **Moderate** | **None€** | 106 (99.07) | 106 (99.07) | 105 (98.13) | 106 (99.07) | 105 (98.13) | 106 (99.07) | 106 (99.07) | 107 (100) | 107 (100) | 106 (99.07) | 106 (99.07) | 106 (99.07) | 106 (99.07) |
| | | **Mild ȸ** | 0 (0) | 1 (0.93) | 2 (1.87) | 1 (0.93) | 2 (1.87) | 1 (0.93) | 1 (0.93) | 0 (0) | 0 (0) | 1 (0.93) | 1 (0.93) | 1 (0.93) | 1 (0.93) |
| | | **Moderateφ** | 1 (0.93) | 0 (0) | 0 (0) | 0 (0) | 0 (0) | 0 (0) | 0 (0) | 0 (0) | 0 (0) | 0 (0) | 0 (0) | 0 (0) | 0 (0) |
| | | **Severe Σ** | 0 (0) | 0 (0) | 0 (0) | 0 (0) | 0 (0) | 0 (0) | 0 (0) | 0 (0) | 0 (0) | 0 (0) | 0 (0) | 0 (0) | 0 (0) |
| | | **Very Severe¥** | 0 (0) | 0 (0) | 0 (0) | 0 (0) | 0 (0) | 0 (0) | 0 (0) | 0 (0) | 0 (0) | 0 (0) | 0 (0) | 0 (0) | 0 (0) |
| | **Severe** | **None€** | 0 (0) | 1 (2) | 1 (2) | 17 (34) | 23 (46) | 35 (70) | 35 (70) | 37 (74) | 40 (80) | 40 (80) | 41 (82) | 41 (82) | 41 (82) |
| | | **Mild ȸ** | 2 (4) | 3 (6) | 18 (36) | 13 (26) | 11 (22) | 4 (8) | 5 (10) | 5 (10) | 1 (2) | 1 (2) | 1 (2) | 1 (2) | 2 (4) |
| | | **Moderateφ** | 21 (42) | 24 (48) | 13 (26) | 11 (22) | 8 (16) | 3 (6) | 2 (4) | 1 (2) | 3 (6) | 3 (6) | 4 (8) | 5 (10) | 4 (8) |
| | | **Severe Σ** | 19 (38) | 14 (28) | 10 (20) | 2 (4) | 2 (4) | 1 (2) | 2 (4) | 2 (4) | 1 (2) | 4 (8) | 2 (4) | 1 (2) | 0 (0) |
| | | **Very Severe¥** | 8 (16) | 8 (16) | 8 (16) | 7 (14) | 6 (12) | 7 (14) | 6 (12) | 5 (10) | 5 (10) | 2 (4) | 2 (4) | 2 (4) | 3 (6) |

* Data are presented as no. (%) unless indicated otherwise. Shortness of breath is classified as Grade1, Grade2, Grade3, Grade4 and Grade5.

**€** Grade1 (none)=Not troubled by breathlessness except on strenuous exercise, **ȸ** Grade 2 (mild)=Short of breath when hurrying on the level or walking up a slight hill, **φ** Grade 3 (moderate)=Walks slower than most people on the level, stops after a mile or so, or stop after 15 minutes walking at own pace, **Σ** Grade 4 (severe)=Stops for breath after walking about 100yds or a few minutes on level ground **¥** Grade 5 (very severe)=Too breathless to leave the house, or breathless when undressing

**Table S7** Variation of oxygen saturation over days*

| No. of days | Honey and *Nigella Sativa* with Standard of care | Standard of care alone |
|---|---|---|
| 1 | 80.92 | 80.91 |
| 2 | 83.32 | 80.68 |
| 3 | 84.70 | 81.45 |
| 4 | 88.52 | 82.38 |
| 5 | 90.92 | 83.81 |
| 6 | 92.64 | 84.18 |
| 7 | 93.52 | 85.52 |
| 8 | 94.14 | 86.96 |
| 9 | 94.62 | 88.33 |
| 10 | 95.14 | 88.22 |
| 11 | 95.72 | 90.72 |
| 12 | 95.70 | 89.39 |
| 13 | 95.46 | 91.50 |

*oxygen saturation is given in percentage unless indicated otherwise.

**Table S8**. Distribution of How sick do you feel over time*

| | | | Number of Days | | | | | | | | | | | | |
|---|---|---|---|---|---|---|---|---|---|---|---|---|---|---|---|
| | | | 1 | 2 | 3 | 4 | 5 | 6 | 7 | 8 | 9 | 10 | 11 | 12 | 13 |
| *Standard Care Alone* | **Moderate** | **None€** | 0 (0) | 4 (3.88) | 1 (0.97) | 8 (7.77) | 11 (10.68) | 28 (27.18) | 44 (42.72) | 61 (59.22) | 77 (74.76) | 81 (78.64) | 90 (87.38) | 97 (94.17) | 98 (95.15) |
| | | **Mild ȸ** | 0 (0) | 0 (0) | 7 (6.8) | 23 (22.33) | 42 (40.78) | 39 (37.86) | 29 (28.16) | 24 (23.3) | 11 (10.68) | 11 (10.68) | 7 (6.8) | 2 (1.94) | 1 (0.97) |
| | | **Moderateφ** | 91 (88.35) | 91 (88.35) | 88 (85.44) | 67 (65.05) | 44 (42.72) | 28 (27.18) | 24 (23.3) | 12 (11.65) | 11 (10.68) | 8 (7.77) | 3 (2.91) | 0 (0) | 1 (0.97) |
| | | **Severe Σ** | 12 (11.65) | 8 (7.77) | 7 (6.8) | 5 (4.85) | 6 (5.83) | 8 (7.77) | 6 (5.83) | 6 (5.83) | 4 (3.88) | 3 (2.91) | 3 (2.91) | 4 (3.88) | 3 (2.91) |
| | **Severe** | **None€** | 0 (0) | 0 (0) | 0 (0) | 0 (0) | 0 (0) | 5 (9.8) | 6 (12) | 11 (21.57) | 14 (28.57) | 19 (38.78) | 22 (46.81) | 26 (56.52) | 28 (60.87) |
| | | **Mild ȸ** | 0 (0) | 0 (0) | 1 (1.92) | 7 (13.46) | 8 (15.69) | 8 (15.69) | 14 (28) | 16 (31.37) | 12 (24.49) | 5 (10.2) | 4 (8.51) | 2 (4.35) | 2 (4.35) |
| | | **Moderateφ** | 1 (1.92) | 6 (11.54) | 8 (15.38) | 13 (25) | 21 (41.18) | 15 (29.41) | 8 (16) | 1 (1.96) | 5 (10.2) | 7 (14.29) | 5 (10.64) | 9 (19.57) | 6 (13.04) |
| | | **Severe Σ** | 51 (98.08) | 46 (88.46) | 43 (82.69) | 32 (61.54) | 22 (43.14) | 23 (45.1) | 22 (44) | 23 (45.1) | 18 (36.73) | 18 (36.73) | 16 (34.04) | 9 (19.57) | 10 (21.74) |
| *Honey-Nigella Sativa* | **Moderate** | **None€** | 2 (1.87) | 2 (1.87) | 6 (5.61) | 48 (44.86) | 58 (54.21) | 98 (91.59) | 102 (95.33) | 104 (97.2) | 106 (99.07) | 106 (99.07) | 106 (99.07) | 106 (99.07) | 106 (99.07) |
| | | **Mild ȸ** | 0 (0) | 10 (9.35) | 54 (50.47) | 40 (37.38) | 44 (41.12) | 7 (6.54) | 3 (2.8) | 3 (2.8) | 1 (0.93) | 1 (0.93) | 1 (0.93) | 1 (0.93) | 1 (0.93) |
| | | **Moderateφ** | 98 (91.59) | 90 (84.11) | 46 (42.99) | 19 (17.76) | 5 (4.67) | 2 (1.87) | 2 (1.87) | 0 (0) | 0 (0) | 0 (0) | 0 (0) | 0 (0) | 0 (0) |
| | | **Severe Σ** | 7 (6.54) | 5 (4.67) | 1 (0.93) | 0 (0) | 0 (0) | 0 (0) | 0 (0) | 0 (0) | 0 (0) | 0 (0) | 0 (0) | 0 (0) | 0 (0) |
| | **Severe** | **None€** | 0 (0) | 0 (0) | 0 (0) | 3 (6) | 7 (14) | 24 (48) | 30 (60) | 35 (70) | 39 (78) | 41 (82) | 40 (80) | 41 (82) | 41 (82) |
| | | **Mild ȸ** | 0 (0) | 0 (0) | 2 (4) | 11 (22) | 23 (46) | 13 (26) | 7 (14) | 7 (14) | 2 (4) | 0 (0) | 1 (2) | 0 (0) | 0 (0) |
| | | **Moderateφ** | 3 (6) | 12 (24) | 21 (42) | 23 (46) | 13 (26) | 5 (10) | 6 (12) | 3 (6) | 3 (6) | 4 (8) | 2 (4) | 4 (8) | 4 (8) |
| | | **Severe Σ** | 47 (94) | 38 (76) | 27 (54) | 13 (26) | 7 (14) | 8 (16) | 7 (14) | 5 (10) | 6 (12) | 5 (10) | 7 (14) | 5 (10) | 5 (10) |

* Data are presented as no. (%) unless indicated otherwise. Responses to “How sick do you feel” are classified as mild, moderate and severe on subjective basis. Patients have been provided with a scale of 1-10, with 1 being the highest and 10 being the lowest

**€** patient is healthy/usual state of health **ȸ** >6 **φ** 4-6 **Σ** 1-3

**Table S9** Variation in emotional status over time*

| | | | Number of Days | | | | | | | | | | | | |
|---|---|---|---|---|---|---|---|---|---|---|---|---|---|---|---|
| | | RYES | **1** | **2** | **3** | **4** | **5** | **6** | **7** | **8** | **9** | **10** | **11** | **12** | **13** |
| *Standard Care Alone* | **Moderate** | **1** | 0 (0) | 0 (0) | 0 (0) | 0 (0) | 0 (0) | 0 (0) | 0 (0) | 0 (0) | 0 (0) | 0 (0) | 0 (0) | 0 (0) | 0 (0) |
| | | **2** | 0 (0) | 0 (0) | 2 (1.94) | 2 (1.94) | 2 (1.94) | 2 (1.94) | 2 (1.94) | 2 (1.94) | 1 (0.97) | 1 (0.97) | 1 (0.97) | 1 (0.97) | 0 (0) |
| | | **3** | 0 (0) | 0 (0) | 0 (0) | 4 (3.88) | 5 (4.85) | 8 (7.77) | 0 (0) | 1 (0.97) | 3 (2.91) | 1 (0.97) | 0 (0) | 0 (0) | 3 (2.91) |
| | | **4** | 31 (30.1) | 29 (28.16) | 24 (23.3) | 20 (19.42) | 9 (8.74) | 3 (2.91) | 2 (1.94) | 1 (0.97) | 0 (0) | 1 (0.97) | 2 (1.94) | 3 (2.91) | 1 (0.97) |
| | | **5** | 62 (60.19) | 59 (57.28) | 48 (46.6) | 20 (19.42) | 12 (11.65) | 3 (2.91) | 8 (7.77) | 2 (1.94) | 0 (0) | 1 (0.97) | 1 (0.97) | 0 (0) | 0 (0) |
| | | **6** | 10 (9.71) | 11 (10.68) | 15 (14.56) | 26 (25.24) | 28 (27.18) | 19 (18.45) | 11 (10.68) | 3 (2.91) | 4 (3.88) | 2 (1.94) | 1 (0.97) | 0 (0) | 0 (0) |
| | | **7** | 0 (0) | 4 (3.88) | 9 (8.74) | 24 (23.3) | 26 (25.24) | 29 (28.16) | 20 (19.42) | 16 (15.53) | 6 (5.83) | 2 (1.94) | 1 (0.97) | 0 (0) | 0 (0) |
| | | **8** | 0 (0) | 0 (0) | 5 (4.85) | 2 (1.94) | 12 (11.65) | 9 (8.74) | 17 (16.5) | 15 (14.56) | 14 (13.59) | 8 (7.77) | 2 (1.94) | 2 (1.94) | 0 (0) |
| | | **9** | 0 (0) | 0 (0) | 0 (0) | 5 (4.85) | 5 (4.85) | 21 (20.39) | 16 (15.53) | 16 (15.53) | 10 (9.71) | 11 (10.68) | 5 (4.85) | 5 (4.85) | 6 (5.83) |
| | | **10** | 0 (0) | 0 (0) | 0 (0) | 0 (0) | 4 (3.88) | 9 (8.74) | 27 (26.21) | 47 (45.63) | 65 (63.11) | 76 (73.79) | 90 (87.38) | 92 (89.32) | 93 (90.29) |
| | **Severe** | **1** | 0 (0) | 0 (0) | 0 (0) | 0 (0) | 0 (0) | 0 (0) | 0 (0) | 0 (0) | 2 (4.08) | 2 (4.08) | 0 (0) | 0 (0) | 0 (0) |
| | | **2** | 3 (5.66) | 2 (3.77) | 4 (7.55) | 5 (9.43) | 4 (7.69) | 3 (5.77) | 5 (9.8) | 4 (7.84) | 2 (4.08) | 0 (0) | 0 (0) | 0 (0) | 0 (0) |
| | | **3** | 7 (13.21) | 13 (24.53) | 16 (30.19) | 18 (33.96) | 11 (21.15) | 14 (26.92) | 10 (19.61) | 8 (15.69) | 5 (10.2) | 5 (10.2) | 5 (10.64) | 4 (8.7) | 2 (4.35) |
| | | **4** | 26 (49.06) | 28 (52.83) | 22 (41.51) | 5 (9.43) | 8 (15.38) | 5 (9.62) | 8 (15.69) | 8 (15.69) | 6 (12.24) | 9 (18.37) | 11 (23.4) | 8 (17.39) | 3 (6.52) |
| | | **5** | 12 (22.64) | 9 (16.98) | 8 (15.09) | 14 (26.42) | 10 (19.23) | 4 (7.69) | 3 (5.88) | 3 (5.88) | 9 (18.37) | 1 (2.04) | 1 (2.13) | 1 (2.17) | 6 (13.04) |
| | | **6** | 5 (9.43) | 0 (0) | 3 (5.66) | 8 (15.09) | 14 (26.92) | 12 (23.08) | 4 (7.84) | 3 (5.88) | 1 (2.04) | 8 (16.33) | 4 (8.51) | 1 (2.17) | 3 (6.52) |
| | | **7** | 0 (0) | 1 (1.89) | 0 (0) | 3 (5.66) | 1 (1.92) | 9 (17.31) | 10 (19.61) | 5 (9.8) | 3 (6.12) | 1 (2.04) | 1 (2.13) | 3 (6.52) | 1 (2.17) |
| | | **8** | 0 (0) | 0 (0) | 0 (0) | 0 (0) | 4 (7.69) | 1 (1.92) | 6 (11.76) | 9 (17.65) | 7 (14.29) | 4 (8.16) | 1 (2.13) | 2 (4.35) | 2 (4.35) |
| | | **9** | 0 (0) | 0 (0) | 0 (0) | 0 (0) | 0 (0) | 4 (7.69) | 1 (1.96) | 4 (7.84) | 4 (8.16) | 5 (10.2) | 2 (4.26) | 3 (6.52) | 2 (4.35) |
| | | **10** | 0 (0) | 0 (0) | 0 (0) | 0 (0) | 0 (0) | 0 (0) | 4 (7.84) | 7 (13.73) | 10 (20.41) | 14 (28.57) | 22 (46.81) | 24 (52.17) | 27 (58.7) |

| | | | | | | | | | | | | | | | |
|---|---|---|---|---|---|---|---|---|---|---|---|---|---|---|---|
| ***Honey-Nigella Sativa*** | **Moderate** | **1** | 0 (0) | 0 (0) | 0 (0) | 0 (0) | 0 (0) | 0 (0) | 0 (0) | 0 (0) | 0 (0) | 0 (0) | 0 (0) | 0 (0) | 0 (0) |
| | | **2** | 0 (0) | 2 (1.87) | 0 (0) | 0 (0) | 0 (0) | 0 (0) | 0 (0) | 0 (0) | 0 (0) | 0 (0) | 0 (0) | 0 (0) | 0 (0) |
| | | **3** | 0 (0) | 0 (0) | 0 (0) | 0 (0) | 0 (0) | 0 (0) | 0 (0) | 0 (0) | 0 (0) | 0 (0) | 0 (0) | 0 (0) | 0 (0) |
| | | **4** | 17 (15.89) | 14 (13.08) | 8 (7.48) | 5 (4.67) | 2 (1.87) | 0 (0) | 0 (0) | 0 (0) | 0 (0) | 0 (0) | 0 (0) | 0 (0) | 0 (0) |
| | | **5** | 70 (65.42) | 57 (53.27) | 22 (20.56) | 7 (6.54) | 4 (3.74) | 2 (1.87) | 1 (0.93) | 0 (0) | 0 (0) | 0 (0) | 1 (0.93) | 1 (0.93) | 1 (0.93) |
| | | **6** | 19 (17.76) | 26 (24.3) | 29 (27.1) | 13 (12.15) | 6 (5.61) | 1 (0.93) | 0 (0) | 0 (0) | 0 (0) | 1 (0.93) | 0 (0) | 0 (0) | 0 (0) |
| | | **7** | 1 (0.93) | 7 (6.54) | 33 (30.84) | 25 (23.36) | 11 (10.28) | 5 (4.67) | 3 (2.8) | 2 (1.89) | 0 (0) | 0 (0) | 0 (0) | 0 (0) | 0 (0) |
| | | **8** | 0 (0) | 1 (0.93) | 12 (11.21) | 23 (21.5) | 22 (20.56) | 2 (1.87) | 3 (2.8) | 3 (2.83) | 2 (1.87) | 0 (0) | 0 (0) | 0 (0) | 0 (0) |
| | | **9** | 0 (0) | 0 (0) | 3 (2.8) | 27 (25.23) | 20 (18.69) | 28 (26.17) | 7 (6.54) | 7 (6.6) | 0 (0) | 2 (1.87) | 0 (0) | 0 (0) | 1 (0.93) |
| | | **10** | 0 (0) | 0 (0) | 0 (0) | 7 (6.54) | 42 (39.25) | 69 (64.49) | 93 (86.92) | 94 (88.68) | 105 (98.13) | 104 (97.2) | 106 (99.07) | 106 (99.07) | 105 (98.13) |
| | **Severe** | **1** | 0 (0) | 0 (0) | 0 (0) | 0 (0) | 0 (0) | 0 (0) | 0 (0) | 0 (0) | 0 (0) | 0 (0) | 0 (0) | 0 (0) | 0 (0) |
| | | **2** | 1 (2) | 1 (2) | 1 (2) | 2 (4) | 2 (4) | 4 (8) | 0 (0) | 1 (2) | 1 (2) | 2 (4) | 0 (0) | 2 (4) | 2 (4) |
| | | **3** | 13 (26) | 8 (16) | 7 (14) | 6 (12) | 5 (10) | 3 (6) | 6 (12) | 3 (6) | 1 (2) | 0 (0) | 2 (4) | 0 (0) | 1 (2) |
| | | **4** | 26 (52) | 30 (60) | 4 (8) | 3 (6) | 1 (2) | 1 (2) | 2 (4) | 4 (8) | 6 (12) | 6 (12) | 6 (12) | 6 (12) | 0 (0) |
| | | **5** | 9 (18) | 5 (10) | 20 (40) | 2 (4) | 1 (2) | 1 (2) | 0 (0) | 0 (0) | 0 (0) | 0 (0) | 0 (0) | 1 (2) | 6 (12) |
| | | **6** | 1 (2) | 3 (6) | 13 (26) | 19 (38) | 7 (14) | 2 (4) | 2 (4) | 0 (0) | 1 (2) | 1 (2) | 1 (2) | 0 (0) | 0 (0) |
| | | **7** | 0 (0) | 3 (6) | 3 (6) | 12 (24) | 15 (30) | 4 (8) | 2 (4) | 1 (2) | 0 (0) | 0 (0) | 0 (0) | 0 (0) | 0 (0) |
| | | **8** | 0 (0) | 0 (0) | 2 (4) | 3 (6) | 12 (24) | 15 (30) | 11 (22) | 3 (6) | 1 (2) | 0 (0) | 0 (0) | 0 (0) | 0 (0) |
| | | **9** | 0 (0) | 0 (0) | 0 (0) | 3 (6) | 5 (10) | 14 (28) | 9 (18) | 8 (16) | 3 (6) | 1 (2) | 0 (0) | 0 (0) | 0 (0) |
| | | **10** | 0 (0) | 0 (0) | 0 (0) | 0 (0) | 2 (4) | 6 (12) | 18 (36) | 30 (60) | 37 (74) | 40 (80) | 41 (82) | 41 (82) | 41 (82) |

*Data are presented as no. (%) unless indicated otherwise. Self-ratings on “Rate your emotional status (RYES)” by patients are classified on subjective basis. Patients have been provided with a scale of 1-10, with 1 being the lowest and 10 being the highest

**Table S10** Variation of clinical status grading over days*

| | | | **Number of Days** | | | | | |
|---|---|---|---|---|---|---|---|---|
| | | CGS | **0** | **4** | **6** | **8** | **10** | 12 |
| ***Standard Care Alone*** | **Moderate** | **1** | 0 (0) | 2 (1.94) | 11 (10.68) | 36 (34.95) | 71 (68.93) | 98 (95.15) |
| | | **2** | 68 (66.02) | 54 (52.43) | 51 (49.51) | 50 (48.54) | 26 (25.24) | 1 (0.97) |
| | | **3** | 35 (33.98) | 42 (40.78) | 35 (33.98) | 12 (11.65) | 2 (1.94) | 0 (0) |
| | | **4** | 0 (0) | 4 (3.88) | 4 (3.88) | 1 (0.97) | 2 (1.94) | 1 (0.97) |
| | | **5** | 0 (0) | 0 (0) | 2 (1.94) | 3 (2.91) | 1 (0.97) | 2 (1.94) |
| | | **6** | 0 (0) | 1 (0.97) | 0 (0) | 1 (0.97) | 1 (0.97) | 1 (0.97) |
| | | **7** | 0 (0) | 0 (0) | 0 (0) | 0 (0) | 0 (0) | 0 (0) |
| | **Severe** | **1** | 0 (0) | 0 (0) | 1 (1.89) | 8 (15.09) | 10 (18.87) | 24 (45.28) |
| | | **2** | 0 (0) | 0 (0) | 1 (1.89) | 1 (1.89) | 13 (24.53) | 0 (0) |
| | | **3** | 0 (0) | 3 (5.66) | 10 (18.87) | 17 (32.08) | 2 (3.77) | 2 (3.77) |
| | | **4** | 23 (43.4) | 24 (45.28) | 23 (43.4) | 12 (22.64) | 16 (30.19) | 13 (24.53) |
| | | **5** | 30 (56.6) | 22 (41.51) | 13 (24.53) | 9 (16.98) | 4 (7.55) | 4 (7.55) |
| | | **6** | 0 (0) | 3 (5.66) | 3 (5.66) | 3 (5.66) | 4 (7.55) | 3 (5.66) |
| | | **7** | 0 (0) | 1 (1.89) | 2 (3.77) | 3 (5.66) | 4 (7.55) | 7 (13.21) |
| ***Honey-Nigella Sativa*** | **Moderate** | **1** | 0 (0) | 25 (23.36) | 68 (63.55) | 97 (90.65) | 103 (96.26) | 106 (99.07) |
| | | **2** | 71 (66.36) | 67 (62.62) | 35 (32.71) | 9 (8.41) | 3 (2.8) | 0 (0) |
| | | **3** | 36 (33.64) | 14 (13.08) | 3 (2.8) | 1 (0.93) | 0 (0) | 0 (0) |
| | | **4** | 0 (0) | 1 (0.93) | 1 (0.93) | 0 (0) | 1 (0.93) | 1 (0.93) |
| | | **5** | 0 (0) | 0 (0) | 0 (0) | 0 (0) | 0 (0) | 0 (0) |
| | | **6** | 0 (0) | 0 (0) | 0 (0) | 0 (0) | 0 (0) | 0 (0) |
| | | **7** | 0 (0) | 0 (0) | 0 (0) | 0 (0) | 0 (0) | 0 (0) |
| | **Severe** | **1** | 0 (0) | 2 (4) | 14 (28) | 29 (58) | 39 (78) | 41 (82) |
| | | **2** | 0 (0) | 1 (2) | 11 (22) | 10 (20) | 2 (4) | 1 (2) |
| | | **3** | 0 (0) | 20 (40) | 13 (26) | 4 (8) | 3 (6) | 5 (10) |
| | | **4** | 21 (42) | 21 (42) | 10 (20) | 5 (10) | 4 (8) | 0 (0) |
| | | **5** | 29 (58) | 6 (12) | 2 (4) | 2 (4) | 1 (2) | 1 (2) |
| | | **6** | 0 (0) | 0 (0) | 0 (0) | 0 (0) | 1 (2) | 2 (4) |
| | | **7** | 0 (0) | 0 (0) | 0 (0) | 0 (0) | 0 (0) | 0 (0) |

*Data are presented as no. (%) unless indicated otherwise. Clinical Grade Score (CGS) was done on seven level ordinal scale as 1: indicated not hospitalized with resumption of normal daily activities; 2, Not hospitalized with limitations on normal daily activities; 3, Hospitalized and not receiving supplemental oxygen; 4, Hospitalized and receiving supplemental oxygen; 5, Hospitalized and receiving oxygen supplementation administered by a high-flow nasal cannula or noninvasive ventilation; 6, Hospitalized and receiving mechanical ventilation; and 7, Death.
